# Glucagon-like peptide-1 receptor activation and mental health: a drug-target Mendelian randomization study

**DOI:** 10.1101/2025.02.12.25322150

**Authors:** Guoyi Yang, Stephen Burgess, C Mary Schooling

## Abstract

Concerns have been raised about the psychiatric safety of glucagon-like peptide-1 receptor (GLP-1R) agonists, but trial evidence suggests that they ameliorate depressive symptoms. We aimed to assess the associations of GLP-1R activation with mental health well-being and the risk of mental health disorders and substance use disorders. We performed drug-target Mendelian randomization and colocalization analyses using the largest relevant genome-wide association studies and replicated in FinnGen. After correcting for multiple comparisons, genetically predicted lower body mass index (BMI) via GLP-1R activation was associated with a better well-being spectrum (0.06 standard deviation [95% confidence interval 0.03-0.08]), lower risk of depression (odds ratio 0.83 [0.74-0.94]), and lower risk of bipolar disorder (odds ratio 0.61 [0.47-0.79]) per 1-kg/m^2^ decrease in BMI. There was also suggestive evidence that genetically predicted lower BMI via GLP-1R activation was associated with lower risk of substance use disorders. These associations were stronger than the associations for genetically predicted lower BMI and lower glycated hemoglobin (HbA1c) based on genome-wide variants. The posterior probabilities of colocalization of BMI and each outcome at the *GLP1R* gene were 59.3% for the well-being spectrum, 3.8% for depression, and 45.9% for bipolar disorder. However, the posterior probabilities of colocalization were > 80% for the well-being spectrum and bipolar disorder when conditioning on the presence of a variant associated with the outcome. This study provides genetic evidence that GLP-1R activation is associated with better mental health well-being and lower risk of bipolar disorder, possibly beyond its effect on BMI and HbA1c.

## Introduction

Glucagon-like peptide-1 receptor (GLP-1R) agonists reduce body weight and blood glucose.^1^ Randomized controlled trials (RCTs) have shown cardiovascular and mortality benefits of GLP-1R agonists.^2,3^ GLP-1R agonists act on the hindbrain, the hypothalamus, and vagal afferents to suppress food intake and reduce body weight.^4–6^ Anti-obesity drugs affecting central nervous systems could have psychiatric effects. For example, the combination of phentermine and topiramate increases depression and anxiety.^7^ Rimonabant was withdrawn from the market due to serious psychiatric adverse events.^8^ In contrast, the combination of naltrexone and bupropion improves depression^9^ and substance use disorder.^10^

Concerns have been raised about the risk of suicidal ideation and self-injury in people using GLP-1R agonists.^11^ However, RCTs showed that GLP-1R agonists moderately reduced depressive symptoms in people who are overweight or obese^12^ and in people with type 2 diabetes.^13^ Animal,^14,15^ observational,^16,17^ and genetic studies^18^ suggested potential benefits of GLP-1R agonists in alcohol use disorder, but RCTs yielded contradictory evidence showing benefits or null effects.^19,20^ Early-stage trials are on-going to investigate the efficacy of GLP-1R agonists in individuals with mental illnesses^21^ and substance use disorders.^22^ A better understanding of the association of GLP-1R activation, the intended target of GLP-1R agonists, with mental health has implications for informing their psychiatric safety and potential repurposing opportunities.

To address this gap, we used Mendelian randomization (MR), an instrumental variable analysis with genetic instruments, to obtain less confounded estimates than conventional observational studies.^23^ We used MR to assess the associations of GLP-1R activation with mental health outcomes including the well-being spectrum, mental health disorders, and substance use disorders. We further investigated whether any associations were explained by lowering body mass index (BMI) or lowering glycated hemoglobin (HbA1c).

## Subjects and Methods

### Ethics approval and consent to participate

All the analyses were conducted using publicly available summary statistics, which does not require ethical approval. Participants of the original studies of publicly available summary statistics provided informed consent.

### Study design

MR relies on the instrumental variable assumptions of relevance (the genetic instruments should be related to the exposure), independence (no common cause of the genetic instruments and the outcome exists), and exclusion restriction (the genetic instruments should be independent of the outcome given the exposure).^23^ This MR study took advantage of the largest relevant publicly available genome-wide association studies (GWASs) (eTable 1). First, we assessed the associations of GLP-1R activation predicted by *GLP1R* variants with mental health outcomes including the well-being spectrum, mental health disorders, and substance use disorders. We compared these associations with the associations for lower BMI and lower HbA1c predicted by genome-wide variants. Second, we performed colocalization analyses to examine whether any associations found for GLP-1R activation were driven by a shared causal variant affecting both BMI or HbA1c and the outcome at the *GLP1R* gene.^24^ Where possible, we conducted sex-specific analyses, because women are more likely to have depression than men,^25^ but boys are more likely to develop neurodevelopmental disorders than girls.^26^

### Genetic instruments

We selected genetic instruments for GLP-1R activation based on their associations with downstream traits related to drug target effects (BMI and HbA1c), because this approach reduces the risk of the variant-outcome association being lost or confounded by pleiotropic effects.^27^ We obtained genetic associations with BMI (*N* = 806,834) from a meta-analysis of the UK Biobank and GIANT^28^ and with HbA1c (*N* = 344,182) from a GWAS of the UK Biobank.^29^ We considered these two sets of instruments separately, because they might represent distinct mechanisms.^30^ For primary analyses, we extracted variants in or near (+-1Mb) the *GLP1R* gene that were uncorrelated (r^2^<0.001) and genome-wide significantly (*p* value <5×10^-8^) associated with BMI to mimic BMI-related mechanisms. We used the same approach based on their associations with HbA1c to select genetic variants mimicking HbA1c-related mechanisms. For sensitivity analyses, we used a less stringent significance threshold for instrument selection (*p* value < 1×10^-5^). For comparison, we also extracted genetic instruments for BMI and HbA1c from across the genome (genome-wide variants) that were uncorrelated (r^2^ <0.001) and genome-wide significantly (*p* value <5×10^-8^) associated with BMI and HbA1c, respectively.

We used coronary artery disease (CAD) and all-cause mortality as positive control outcomes, because RCTs have shown that GLP-1R agonists reduce cardiovascular events and death.^2,3^ We obtained genetic associations with CAD (181,522 cases/984,168 controls) from a meta-analysis of the CARDIoGRAMplusC4D consortium and the UK Biobank.^31^ We obtained genetic associations with parental mortality (609,139 cases/403,101 controls), determined by parental attained age and their alive/dead status, from a meta-analysis of the UK Biobank and LifeGen as a measure of all-cause mortality,^32^ because it has greater power than participant’s mortality.

### Genetic associations with mental health outcomes

We obtained sex-combined and sex-specific genetic associations with mental health outcomes from the largest relevant publicly available GWASs (eTable 1).^29,33–47^ Primary outcomes included the well-being spectrum, mental health disorders (depression, bipolar disorder, post-traumatic stress disorder (PTSD), schizophrenia, anorexia nervosa, attention deficit hyperactivity disorder (ADHD), autism spectrum disorder, and Tourette’s syndrome), and substance use disorders. Secondary outcomes included four dimensions of mental health well-being (life satisfaction, positive affect, neuroticism, and depressive symptoms) and subtypes of mental health disorders (postpartum depression and bipolar disorder I and II) and substance use disorders (cannabis use disorder and alcohol dependence). Secondary outcomes also included the Alcohol Use Disorders Identification Test (AUDIT) total score, score for alcohol consumption, and score for alcohol problems. We obtained genetic associations with relevant outcomes from the FinnGen R12 for replication,^48^ where possible. Genetic associations with binary outcomes obtained using linear regression were transformed into odds ratio (OR) using an established approximation.^49^

### MR analysis

We used the F-statistic to assess instrument strength, approximated by the square of SNP-exposure association divided by the square of its standard error.^50^ An F-statistic larger than 10 suggests weak instrument bias is unlikely. We aligned SNPs on the same allele for exposure and outcome, and used proxy SNPs (r^2^>0.8), where possible, when SNPs were not available in the outcome GWAS. We calculated Wald estimates by dividing genetic association with outcome by genetic association with exposure. We obtained MR estimates by meta-analyzing Wald estimates using inverse variance weighting (IVW) with fixed effects for three SNPs or fewer and random effects for four SNPs or more.^51^ To assess the robustness of IVW estimates, we conducted sensitivity analyses using the weighted median,^52^ MR Egger,^53^ and MR using the robust adjusted profile score (MR-RAPS).^54^ We used the Cochran’s Q statistic^55^ to test heterogeneity in Wald estimates and the MR Egger intercept^53^ to assess directional pleiotropy. We used the Steiger filtering to identify the SNPs explaining more variance in the outcome than in the exposure (*p* value < 0.05).^56^ We conducted sensitivity analyses excluding the SNPs identified to assess whether MR estimates were affected by reverse causation. We used a Bonferroni-corrected significance level of α=0.05/10=0.005 given ten primary outcomes. We considered nominally significant associations (*p* value <0.05) that did not reach Bonferroni-corrected significance as suggestive evidence. We tested differences by sex using a two-sided z-test.^57^

### Colocalization

We performed pairwise colocalization analyses in a Bayesian framework to assess whether any associations found for GLP-1R activation were driven by a shared causal variant affecting both BMI or HbA1c and the outcome or were confounded by linkage disequilibrium.^24^ We included variants (minor allele frequency >0.1%) in or near (+- 1Mb) the *GLP1R* gene. We set the prior probabilities as recommended, that is 10^-4^ for a variant associated with BMI or HbA1c, 10^-4^ for a variant associated with the outcome, and 10^-5^ for a variant associated with both traits.^24^ We assessed the posterior probability of several hypotheses, including H_0_ (no association with either trait), H_1_, (association with BMI or HbA1c only); H_2_, (association with the outcome only); H_3_, (associations of two independent variants and one for each trait); H_4_ (associations of one shared variant with both traits).^24^ A posterior probability of H_4_ larger than 0.80 suggests colocalization.^24^ The power to detect colocalization is low when the variants are not strongly associated with the outcome, so we also calculated the posterior probability of colocalization (H_4_) conditional on the presence of a variant associated with the outcome as H_4_/(H_2_ + H_3_ + H_4_).^58^ We reported the posterior probabilities of H_0_-H_4_ and conditional H_4_ and the variant with the largest posterior probability of being the shared causal variant.

We also used the Hypothesis Prioritisation for multi-trait Colocalization (HyPrColoc)^59^ to assess colocalization across multiple traits identified for GLP-1R activation. We set the prior probability for a variant associated with at least one trait as 10^-4^ and the conditional prior probability for a variant associated with an additional trait given that the variant is associated with at least one other trait as 0.02.^59^ We reported clusters of putatively colocalized traits and the posterior probability of colocalization.

All statistical analyses were conducted using R version 4.2.1 and the packages “ieugwasr”, “TwoSampleMR”, “MendelianRandomization”, “mr.raps, “metafor”, “coloc”, and “hyprcoloc”.

## Results

### Genetic instruments

For primary analyses, we used two *GLP1R* SNPs (rs17757975 and rs4714290) genome-wide significantly associated with BMI to predict the impact of GLP-1R activation as indicated by lower BMI; we used one *GLP1R* SNP (rs10305518) genome-wide significantly associated with HbA1c to predict the impact of GLP-1R activation as indicated by lower HbA1c (eTable 2). For sensitivity analyses, we used three *GLP1R* SNPs for BMI and two *GLP1R* SNPs for HbA1c based on a more lenient significance threshold (*p* value < 1×10^-5^) (eTable 2). The *GLP1R* SNPs for BMI were not associated with HbA1c and vice versa (eTable 3), which substantiated that the two sets of genetic instruments represent distinct mechanisms of GLP-1R activation.^30^ For comparison, we also extracted 516 (overall), 303 (women), and 256 (men) SNPs for BMI and 318 (overall), 200 (women), 174 (men) SNPs for HbA1c from across the genome (eTable 4). The F-statistics of the primary instruments were all >10 (eTables 2 and 4).

In positive control analyses, genetically predicted lower BMI and lower HbA1c via GLP-1R activation were associated with lower CAD risk and all-cause mortality, despite wide confidence intervals (CIs) (eFigure 1). The point estimates for GLP-1R activation were comparable to those for genetically predicted lower BMI and lower HbA1c based on genome-wide variants (eFigure 1 and eTable 5).

### Associations of GLP-1R activation with mental health well-being

Genetically predicted lower BMI via GLP-1R activation was associated with a better well-being spectrum, including life satisfaction, positive affect, neuroticism, and depressive symptoms (Figure 1). The association with depressive symptoms was evident specifically in women (eFigure 2). These associations were stronger than the associations for genetically predicted lower BMI using genome-wide variants (Figure 1). However, genetically predicted lower HbA1c via GLP-1R activation had little association with mental health well-being (Figure 1). Sensitivity analyses gave consistent estimates, and the Steiger filtering identified no SNP that explained more variance in the outcome than in the exposure (eTable 6-7).

**Figure 1.**
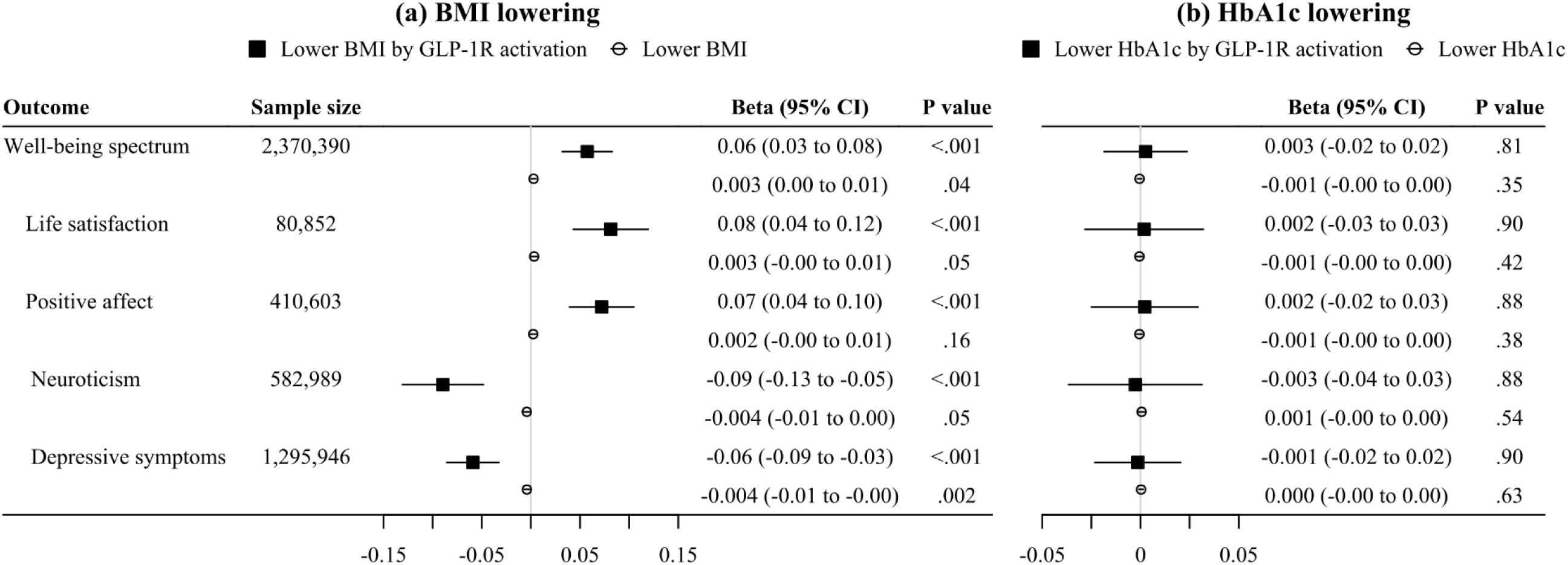
IVW MR estimates for the associations of GLP-1R activation with mental health well-being. BMI, body mass index; GLP-1R, glucagon-like peptide-1 receptor; HbA1c, glycated hemoglobin; IVW, inverse variance weighted; MR, Mendelian randomization. Black squares denote genetically predicted (a) lower BMI or (b) lower HbA1c via GLP-1R activation based on *GLP1R* variants. White circles denote genetically predicted (a) lower BMI or (b) lower HbA1c based on genome-wide variants. Positive associations with the well-being spectrum, life satisfaction, and positive affect and negative associations with neuroticism and depressive symptoms indicate better mental health well-being. Estimates are presented in standard deviation units for mental health well-being per 1-kg/m^2^ decrease in BMI or per 1-mmol/mol decrease in HbA1c.

### Associations of GLP-1R activation with the risk of mental health disorders

Genetically predicted lower BMI via GLP-1R activation was associated with lower risk of depression, postpartum depression, bipolar disorder, bipolar disorder I and II, and ADHD (Figure 2). The association with depression was possibly stronger in women than men, although the *p* value for sex difference was 0.35 (eFigure 3). These associations were stronger than the associations for genetically predicted lower BMI using genome-wide variants (Figure 2). Genetically predicted lower HbA1c via GLP-1R activation was associated with higher risk of anorexia nervosa but lower risk of Tourette syndrome, with stronger associations than the associations for genetically predicted lower HbA1c using genome-wide variants (Figure 2). These results were robust to different MR methods and the exclusion of SNPs identified by the Steiger filtering (eTable 8).

**Figure 2.**
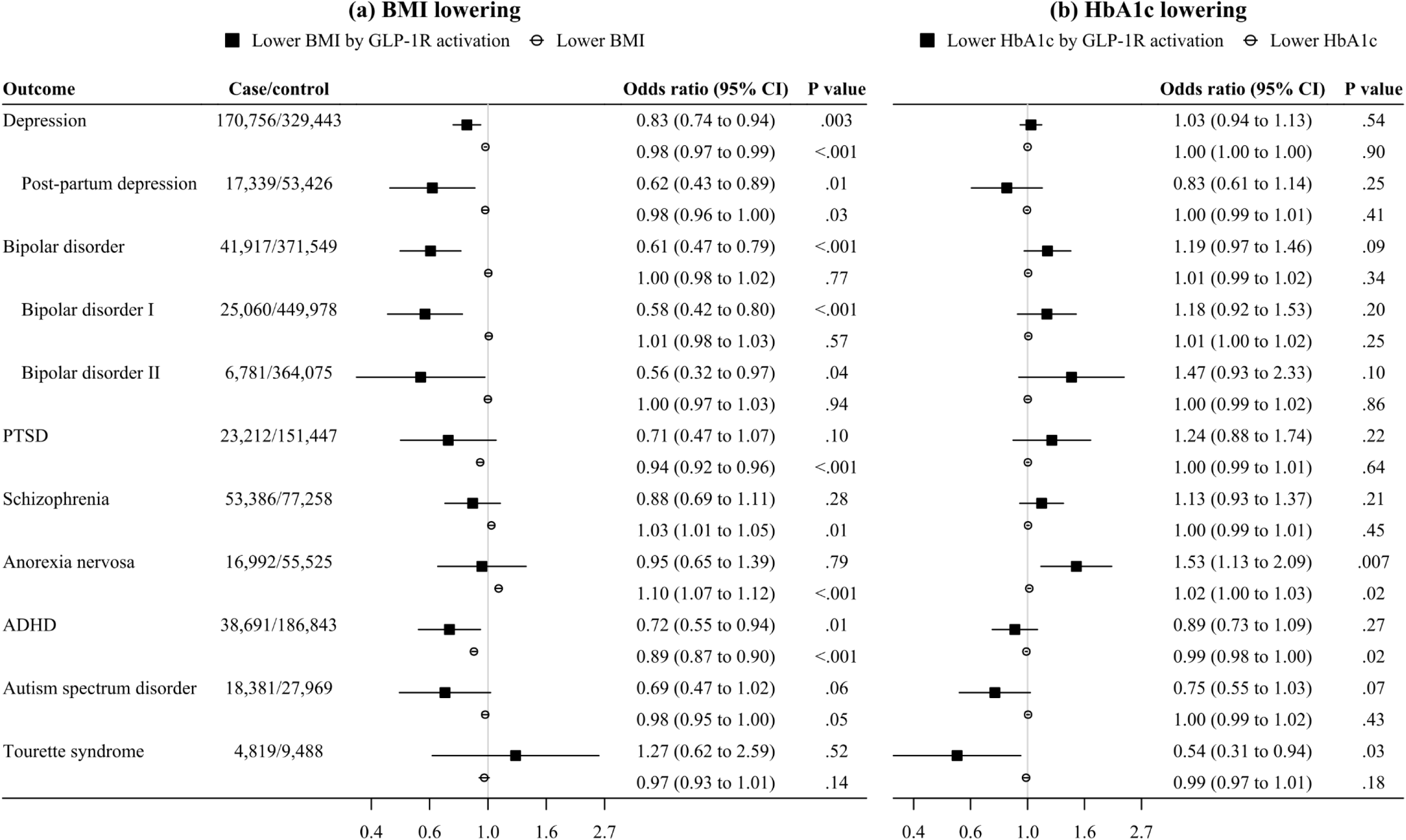
IVW MR estimates for the associations of GLP-1R activation with the risk of mental health disorders. ADHD, attention deficit hyperactivity disorder; BMI, body mass index; GLP-1R, glucagon-like peptide-1 receptor; HbA1c, glycated hemoglobin; IVW, inverse variance weighted; MR, Mendelian randomization; PTSD, post-traumatic stress disorder. Black squares denote genetically predicted (a) lower BMI or (b) lower HbA1c via GLP-1R activation based on *GLP1R* variants. White circles denote genetically predicted (a) lower BMI or (b) lower HbA1c based on genome-wide variants. Estimates are presented as odds ratio per 1-kg/m^2^ decrease in BMI or per 1-mmol/mol decrease in HbA1c.

### Associations of GLP-1R activation with the risk of substance use disorders

Genetically predicted lower BMI via GLP-1R activation was associated with lower risk of substance use disorders and lower AUDIT score for alcohol problems (Figure 3). These associations were stronger than the associations for genetically predicted lower BMI using genome-wide variants. However, genetically predicted lower HbA1c via GLP-1R activation had little association with the risk of substance use disorders (Figure 3). Sensitivity analyses using different MR methods and excluding the SNPs identified by the Steiger filtering gave similar interpretations (eTable 9).

**Figure 3.**
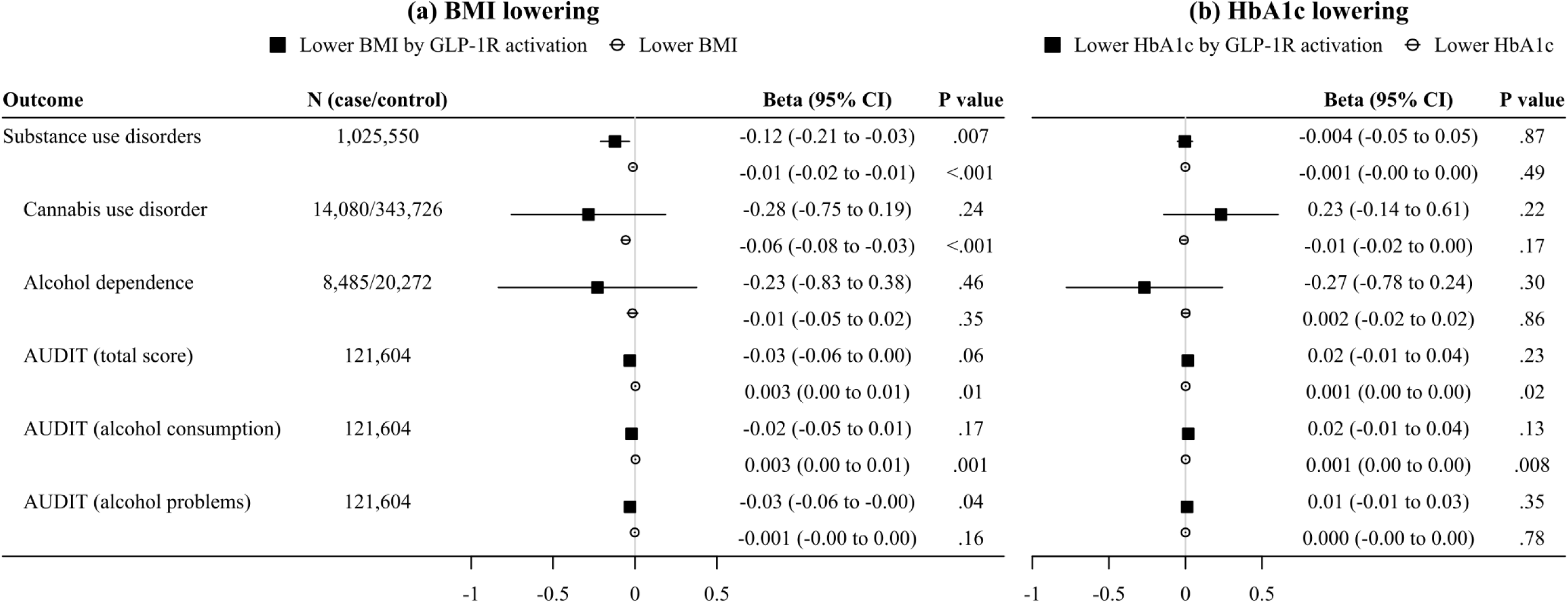
IVW MR estimates for the associations of GLP-1R activation with the risk of substance use disorders and AUDIT scores. AUDIT, Alcohol Use Disorders Identification Test; BMI, body mass index; GLP-1R, glucagon-like peptide-1 receptor; HbA1c, glycated hemoglobin; IVW, inverse variance weighted; MR, Mendelian randomization. Black squares denote genetically predicted (a) lower BMI or (b) lower HbA1c via GLP-1R activation based on *GLP1R* variants. White circles denote genetically predicted (a) lower BMI or (b) lower HbA1c based on genome-wide variants. Positive associations with AUDIT scores indicate higher risk of alcohol use disorders. Estimates are presented in standard deviation unit for substance use disorder risk, as logodds for cannabis use disorder and alcohol dependence, and as log10 transformed score for AUDIT scores per 1-kg/m^2^ decrease in BMI or per 1-mmol/mol decrease in HbA1c.

### Replication

In FinnGen, genetically predicted lower BMI via GLP-1R activation was associated with lower risk of depression, bipolar disorder, PTSD, ADHD, substance abuse, and alcohol dependence (eFigure 4). Genetically predicted lower HbA1c via GLP-1R activation had little association with mental health outcomes except for depression (eFigure 4). Nevertheless, this association with depression was attenuated to null when using the two SNPs extracted based on a more lenient significance threshold (eTable 10).

### Colocalization

After correcting for multiple comparisons, genetically predicted lower BMI via GLP-1R activation remained significantly associated with the well-being spectrum, life satisfaction, positive affect, neuroticism, depressive symptoms, depression, bipolar disorder, and bipolar disorder I (*p* values <0.005). Pairwise colocalization analyses were performed for BMI and each significant outcome in or near the *GLP1R* gene. The posterior probabilities of colocalization were <80% for all the significant outcomes (Figure 4). However, conditioning on the presence of a variant associated with the outcome, the posterior probabilities of colocalization were all >80% except for depression (Figure 4), suggesting that some lack of colocalization was due to limited power. The posterior probabilities of two independent variants associated with each trait were <15% for all the significant outcomes (eTable 11), suggesting that confounding by linkage disequilibrium was unlikely. Across the *GLP1R* gene region, rs4714290, rs847784 (a proxy for rs4714290, r^2^ = 0.89), or rs17757975 had the largest posterior probability of being the shared causal variant, which substantiated their validity as genetic instruments.

**Figure 4.**
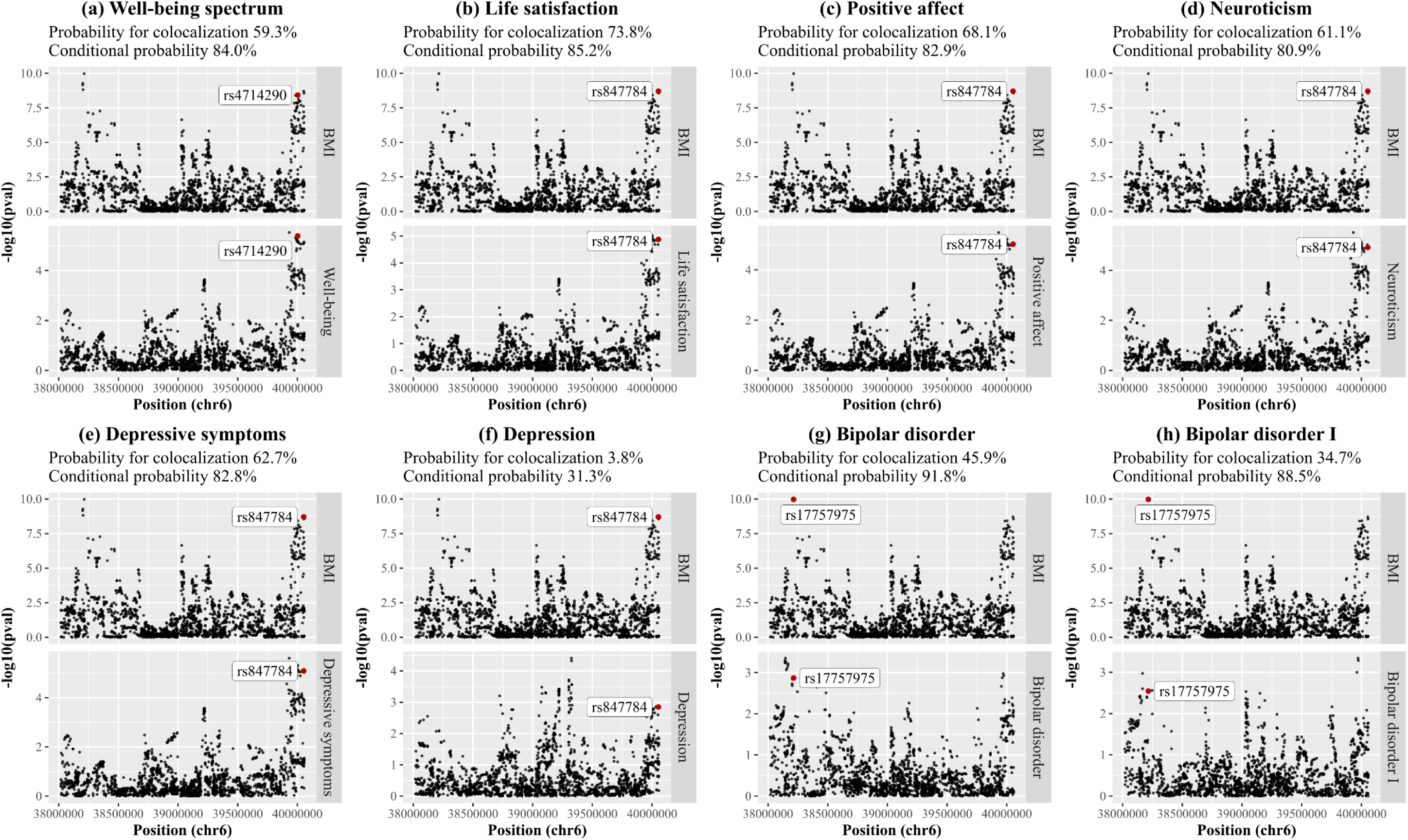
Pairwise colocalization analyses for BMI and each significant outcome at the *GLP1R* gene. BMI, body mass index. Probability for colocalization means the posterior probability of a shared variant associated with both traits; conditional probability means the posterior probability of a shared variant associated with both traits conditional on the presence of a variant associated with the outcome. Given the presence of a shared variant associated with both traits, the labeled variant had the largest probability of being the shared causal variant.

Multi-trait colocalization analyses were performed across BMI and all the significant outcomes in or near the *GLP1R* gene. We identified BMI, the well-being spectrum, life satisfaction, positive affect, neuroticism, depressive symptoms as a cluster of putatively colocalized traits with the posterior probability of colocalization 51.9%.

## Discussion

This drug-target MR study suggests that GLP-1R activation is associated with a better well-being spectrum and lower risk of bipolar disorder. We also provide suggestive evidence that GLP-1R activation is associated with lower risk of substance use disorders. These associations are likely beyond the effects of GLP-1R activation on BMI and HbA1c.

We found that genetically predicted GLP-1R activation was associated with a better well-being spectrum, lower risk of depression, and lower risk of bipolar disorder. Consistently, RCTs have shown that GLP-1R agonists moderately reduce depression symptoms in overweight people^12^ and in people with type 2 diabetes.^13^ A small open-label trial showed that liraglutide, as an adjunct to existing pharmacotherapy, improved cognitive function^60^ and brain volumes^61^ in people with depression or bipolar disorder. These psychiatric benefits of GLP-1R activation could be driven by the neuroprotective effects of GLP-1 including enhanced neurogenesis, improved barrier function, reduced neuroinflammation, and decreased oxidative stress.^62^

Interestingly, the associations of GLP-1R activation with favorable mental health outcomes were more evident when proxied via BMI than via HbA1c, highlighting the importance of BMI-related mechanisms. A recent study showed that GLP-1R agonists target the hindbrain dorsal vagal complex to decrease food intake and body weight.^4^ Actions on the hypothalamus and vagal afferents also contribute to the weight loss on GLP-1R agonists.^5,6^ GLP neurons in the dorsal vagal complex moderate stress responses and stimulate hypothalamic-pituitary-adrenal (HPA) axis activity, which has been implicated in the pathology of depression^63^ and bipolar disorder.^64^ Notably, the HPA axis is also related to cognitive function,^63,64^ consistent with an RCT showing that dulaglutide reduces cognitive impairment in people with type 2 diabetes.^65^ Additional studies are warranted to elucidate the mechanisms underlying potential benefits of GLP-1R agonists for mental health.

We found that genetically predicted GLP-1R activation was possibly associated with lower risk of depression specifically in women and lower risk of postpartum depression. We caution that the power to detect sex differences may have been limited by small or unequal sample sizes of sex-stratified GWAS. Women are about twice as likely as men to have depression during their lifetime with the difference starting at puberty.^25^ A meta-analysis of RCTs showed that GLP-1R agonists decreased testosterone and increased sex hormone-binding globulin in women with polycystic ovary syndrome,^66^ but their effects on estrogen are less clear. Sex hormones might contribute to GLP-1R activation improving depression in women, given their potential roles in modulating serotonergic neurotransmission, cortisol, and oxytocin.^67^

Genetically predicted GLP-1R activation was suggestively associated with lower risk of ADHD and Tourette syndrome but had a null association with autism spectrum disorder. Although RCTs have shown the short-term (no more than 26 weeks) safety of GLP-1R agonists in children and adolescents,^68^ RCTs typically do not capture the long latency of neurodevelopmental disorders. Our study adds to the evidence by showing little adverse effect of lifelong GLP-1R activation on these disorders. However, GLP-1R activation was associated with higher risk of anorexia nervosa when proxied via HbA1c. GLP-1R activation increases insulin secretion and sensitivity and lowers glucose.^69^ A previous MR study suggested an inverse association of fasting insulin with the risk of anorexia nervosa,^70^ indicating a role of insulin sensitivity.

We provide suggestive evidence that GLP-1R activation was associated with lower risk of substance use disorders and lower AUDIT score for alcohol problems, which were replicated in FinnGen. Our finding aligns with a recent RCT showed that eight-week treatment with semaglutide reduced alcohol consumption and craving in people with alcohol use disorder.^20^ In contrast, another RCT including people with alcohol use disorder showed that 26 weeks of treatment with exenatide did not significantly reduce heavy drinking days, although exenatide attenuated fMRI alcohol cue reactivity in brain areas for drug reward and addiction.^19^ Further evidence from RCTs is needed to determine the repurposing opportunities of GLP-1R agonists for treating substance use disorders.

We presented the MR estimates of GLP-1R activation in effect sizes of BMI or HbA1c reduction for interpretability. However, this scaling does not imply that weight loss or glucose lowering is the sole mechanism of action for the psychiatric effects of GLP-1R activation. Indeed, the associations of GLP-1R activation with mental health outcomes appeared stronger than those expected from lowering BMI and HbA1c. Correspondingly, RCTs have showed that semaglutide reduces cardiovascular events and mortality more than expected from the reduction in body weight,^12^ and that GLP-1R agonists outperform other anti-diabetes drugs.^71^ Apart from reducing body weight and glucose, GLP-1R agonists also decrease blood pressure, low-density lipoprotein cholesterol, and inflammation and enhance kidney function.^2,3^ The multifaceted effects of GLP-1R agonists could be partially explained by 5’ adenosine monophosphate-activated protein kinase (AMPK) activation.^72^ Taken together, our findings suggest that the psychiatric benefits of GLP-1R activation likely extend beyond its effect on BMI and HbA1c.

This drug-target MR study comprehensively explored the associations of GLP-1R activation with mental health outcomes. Our investigation has implications for the psychiatric safety and repurposing opportunities of GLP-1R agonists, particularly in people with mental health disorders, who usually have higher risk of obesity and type 2 diabetes but are undertreated. This study has several limitations. First, MR relies on three rigorous assumptions, that is genetic instruments should be related to the exposure, share no common cause with the outcome, and be independent of the outcome given the exposure.^23^ The F-statistics were >10, suggesting minimal weak instrument bias. We validated the genetic instruments using CAD and all-cause mortality as positive control outcomes. We used MR methods with different assumptions about instrumental validity, which gave consistent estimates. Second, we selected genetic instruments for GLP-1R activation based on their associations with BMI and HbA1c, but additional underlying factors may explain their psychiatric effects. Instrumenting on BMI and HbA1c could have masked some effects of GLP-1R activation. Third, the small number of genetic instruments for GLP-1R activation, even when applying a more lenient significance threshold for instrument selection, may have limited the detection of possible causal effects. Fourth, we obtained genetic associations with BMI from a meta-analysis of the UK Biobank and GIANT and with HbA1c from the UK Biobank, which overlaps with some outcome GWASs. Two-sample MR methods in a one-sample setting perform well in terms of bias and precision in large biobanks, except for MR Egger which can result in bias reflecting the direction and magnitude of confounding.^73^ We replicated the results using outcome GWASs from FinnGen, which has no overlap with the UK Biobank or GIANT. Fifth, the power to detect colocalization was insufficient. However, the posterior probabilities of two independent variants associated with each trait were <15% for all the significant outcomes, suggesting that the associations found were unlikely driven by confounding due to linkage disequilibrium. Sixth, MR can be open to selection bias from selecting on genetic makeup and mental health disorders. Nevertheless, such selection should bias towards a harmful effect of GLP-1R activation, which could not explain its associations with favorable mental health outcomes. Finally, mental health disorders sometimes originate during childhood or adolescence, but GLP-1R agonists are primarily used in adults. Our MR study assessed lifelong effects of GLP-1R activation, which cannot directly inform the short-term effects of GLP-1R agonists. RCTs are warranted to determine the clinical relevance of these findings in the future.

## Conclusions

This drug-target MR study provides genetic evidence that GLP-1R activation is associated with better mental health well-being and lower risk of bipolar disorder, possibly beyond its effect on BMI and HbA1c.

## Supporting information

eFigures 1-4 and eTables 1-11

## Data Availability

Summary statistics analyzed are available in the website https://doi.org/10.5281/zenodo.1251813 for BMI, http://www.nealelab.is/uk-biobank/ for HbA1c, https://www.cardiogramplusc4d.org/data-downloads/ for CAD, https://www.ebi.ac.uk/gwas/publications/30642433 for parental mortality, https://www.ebi.ac.uk/gwas/publications/30643256 for mental health well-being, and https://pgc.unc.edu/for-researchers/download-results/ and https://www.finngen.fi/en/access_results for mental health disorders and substance use disorders.

## Acknowledgements

The authors acknowledge Pulit SL, et al., the Neale lab, the CARDIoGRAMplusC4D Consortium, Timmers PR, et al., Baselmans BML, et al., the Psychiatric Genomics Consortium, and FinnGen for their publicly available summary data.

## Availability of data and materials

Summary statistics analyzed are available in the website

https://doi.org/10.5281/zenodo.1251813 for BMI, http://www.nealelab.is/uk-biobank/ for HbA1c, https://www.cardiogramplusc4d.org/data-downloads/ for CAD, https://www.ebi.ac.uk/gwas/publications/30642433 for parental mortality, https://www.ebi.ac.uk/gwas/publications/30643256 for mental health well-being, and https://pgc.unc.edu/for-researchers/download-results/ and https://www.finngen.fi/en/access_results for mental health disorders and substance use disorders.

## Author contributions

GY and CMS designed the study. GY undertook analyses with feedback from SB and CMS. GY drafted the manuscript with critical feedback and revisions from SB and CMS. All authors read and approved the final version of the manuscript. GY had full access to all the data in the study and takes responsibility for the integrity of the data and the accuracy of the data analysis.

## Artificial intelligence (AI)-assisted technologies

The authors declared that AI-assisted technologies were not used in this study.

