## Supplementary material for "Glucagon-like peptide-1 receptor activation and mental health: a drug-target Mendelian randomization study": eFigures 1-4 and eTables 1-11

**Online-Only Supplements**

**eTable 1.** A summary of genome-wide association studies used in this study.

**eTable 2.** SNP-specific estimates for genetic instruments for GLP-1R activation on BMI (kg/m$^2$) and HbA1c (mmol/mol).

**eTable 3.** MR estimates for the associations of GLP-1R activation with HbA1c (mmol/mol) and BMI (kg/m$^2$).

**eTable 4.** SNP-specific estimates for genetic instruments for BMI (kg/m$^2$) and HbA1c (mmol/mol).

**eFigure 1.** IVW MR estimates for the associations of GLP-1R activation with the risk of coronary artery disease and all-cause mortality in comparison with those for lower BMI and lower HbA1c.

**eTable 5.** Sensitivity MR analyses for the associations of GLP-1R activation, lower BMI, and lower HbA1c with the risk of coronary artery disease and all-cause mortality.

**eFigure 2.** IVW MR estimates for sex-specific associations of GLP-1R activation with the risk of depressive symptoms in comparison with those for lower BMI and lower HbA1c.

**eTable 6.** Sensitivity MR analyses for the associations of GLP-1R activation, lower BMI, and lower HbA1c with mental health well-being.

**eTable 7.** Sensitivity MR analyses for sex-specific associations of GLP-1R activation, lower BMI, and HbA1c with the risk of depression symptoms.

**eFigure 3.** IVW MR estimates for sex-specific associations of GLP-1R activation with the risk of depression, schizophrenia, and ADHD in comparison with those for lower BMI and lower HbA1c.

**eTable 8.** Sensitivity MR analyses for the associations of GLP-1R activation, lower BMI, and lower HbA1c with the risk of mental health disorders.

**eTable 9.** Sensitivity MR analyses for the associations of GLP-1R activation, lower BMI, and lower HbA1c with the risk of substance use disorders.

**eFigure 4.** IVW MR estimates for the associations of GLP-1R activation with the risk of mental health outcomes in comparison with those for lower BMI and lower HbA1c in FinnGen.

**eTable 10.** Sensitivity MR analyses for the associations of GLP-1R activation, lower BMI, and lower HbA1c with mental health outcomes in FinnGen.

**eTable 11.** The posterior probabilities of different hypotheses in pairwise colocalization analyses for BMI and each significant outcome at the *GLP1R* gene.

**eTable 1. A summary of genome-wide association studies used in this study.**

| Category | Phenotype | Study | Ancestry | Sex | *N* (case/control) | Covariates | PMID |
|---|---|---|---|---|---|---|---|
| Genetic instruments | Body mass index | UKB, GIANT | European | Overall | 806,834 | Sex, age at assessment, age at assessment$^2$, and assessment center (UKB); sex, age, age$^2$, and study-specific covariates (GIANT) | 30239722 |
| | Body mass index | UKB, GIANT | European | Women | 434,794 | Age at assessment, age at assessment$^2$, and assessment center (UKB); age, age$^2$, and study-specific covariates (GIANT) | 30239722 |
| | Body mass index | UKB, GIANT | European | Men | 374,756 | Age at assessment, age at assessment$^2$, and assessment center (UKB); age, age$^2$, and study-specific covariates (GIANT) | 30239722 |
| | HbA1c | UKB | European | Overall | 344,182 | Age, age$^2$, inferred sex, age × inferred sex, age$^2$ × inferred sex, and the first 20 PCs | Neale lab |
| | HbA1c | UKB | European | Women | 185,022 | Age, age$^2$, and the first 20 PCs | Neale lab |
| | HbA1c | UKB | European | Men | 159,160 | Age, age$^2$, and the first 20 PCs | Neale lab |
| Positive control outcome | Coronary artery disease | UKB, CARDIoGRAMplusC4D | Mixed (European >95%) | Overall | 181,522/984,168 | Study-specific covariates | 36474045 |
| | Parental mortality | UKB, LifeGen | European | Overall | 609,139/403,101 | Genotyping batch and array, the first 40 PCs of relatedness, and participant sex (UKB); participant sex, the first 10 PCs, and study-specific covariates (LifeGen) | 30642433 |
| | Well-being spectrum | Meta-analysis | European | Overall | 2,370,390 | Study-specific covariates | 30643256 |

| | | | | | | | |
|---|---|---|---|---|---|---|---|
| | Depression | UKB, PGC | European | Overall | 170,756/329,443 | Sex, age, genotyping array, and 8 PCs (UKB); study-specific covariates (PGC) | 30718901 |
| | Depression | UKB | European | Women | 13,492/180,661 | Age, $age^2$, and the first 20 PCs | Neale lab |
| | Depression | UKB | European | Men | 7,156/159,832 | Age, $age^2$, and the first 20 PCs | Neale lab |
| | Bipolar disorder | Meta-analysis | European | Overall | 41,917/371,549 | The first five PCs and any others as required | 34002096 |
| | PTSD | PGC, UKB | European | Overall | 23,212/151,447 | The first five PCs (unrelated subjects); the first five PCs and a genetic relatedness matrix (family and twin studies); the first six PCs, batch, and center indicator variables (UKB) | 31594949 |
| Primary outcome | Schizophrenia | PGC | European | Overall | 53,386/77,258 | The first four PCs and any other PC that was nominally significantly associated with the phenotype | 35396580 |
| | Schizophrenia | PGC | European | Women | 17,710/36,803 | The first four PCs and any other PC that was nominally significantly associated with the phenotype | 35396580 |
| | Schizophrenia | PGC | European | Men | 33,097/35,190 | The first four PCs and any other PC that was nominally significantly associated with the phenotype | 35396580 |
| | Anorexia nervosa | PGC, UKB | European | Overall | 16,992/55,525 | The first five PCs and any other PC that was nominally associated with the phenotype | 31308545 |
| | Attention deficit hyperactivity disorder | iPSYCH, deCODE, PGC | European | Overall | 38,691/186,843 | The first ten PCs (iPSYCH); sex, year of birth, county of origin, population stratification, and | 36702997 |

| | | | | | | |
|---|---|---|---|---|---|---|
| | | | | | relatedness (deCODE); PCs and study-specific covariates (PGC) | |
| Attention deficit hyperactivity disorder | iPSYCH, PGC | European | Women | 4,945/16,246 | The first four PCs and any other PC that was nominally significantly associated with the phenotype (iPSYCH); the first five or ten PCs or indicator variables coding for site ID (PGC) | 29325848 |
| Attention deficit hyperactivity disorder | iPSYCH, PGC | European | Men | 14,154/17,948 | The first four PCs and any other PC that was nominally significantly associated with the phenotype (iPSYCH); the first five or ten PCs or indicator variables coding for site ID (PGC) | 29325848 |
| Autism spectrum disorder | iPSYCH, PGC | European | Overall | 18,381/27,969 | The first four PCs and any other PC that was nominally significantly associated with the phenotype (iPSYCH); study-specific covariates (PGC) | 30804558 |
| Tourette's syndrome | Meta-analysis | European | Overall | 4,819/9,488 | The first four population stratification components and any additional population stratification component that was nominally significantly associated with the phenotype | 30818990 |
| Substance use disorders | Meta-analysis | European | Overall | 1,025,550 | Study-specific covariates | 37250466 |
| Life satisfaction | Meta-analysis | European | Overall | 80,852 | Study-specific covariates | 30643256 |

| | | | | | | | |
|---|---|---|---|---|---|---|---|
| | Positive affect | Meta-analysis | European | Overall | 410,603 | Study-specific covariates | 30643256 |
| | Neuroticism | Meta-analysis | European | Overall | 582,989 | Study-specific covariates | 30643256 |
| | Depressive symptoms | Meta-analysis | European | Overall | 1,295,946 | Study-specific covariates | 30643256 |
| | Depressive symptoms (seen a general practitioner) | UKB | European | Women | 79,976/112,704 | Age, age$^2$, and the first 20 PCs | Neale lab |
| | Depressive symptoms (seen a general practitioner) | UKB | European | Men | 43,552/122,461 | Age, age$^2$, and the first 20 PCs | Neale lab |
| | Depressive symptoms (seen a psychiatrist) | UKB | European | Women | 23,975/169,269 | Age, age$^2$, and the first 20 PCs | Neale lab |
| | Depressive symptoms (seen a psychiatrist) | UKB | European | Men | 17,258/149,033 | Age, age$^2$, and the first 20 PCs | Neale lab |
| Secondary outcome | Post-partum depression | Meta-analysis | European | Women | 17,339/53,426 | Study-specific covariates | 37849304 |
| | Bipolar disorder I | Meta-analysis | European | Overall | 25,060/449,978 | The first five PCs and any others as required | 34002096 |
| | Bipolar disorder II | Meta-analysis | European | Overall | 6,781/364,075 | The first five PCs and any others as required | 34002096 |
| | Cannabis use disorder | iPSYCH, deCODE, PGC | European | Overall | 14,080/343,726 | Five ancestral PCs, data processing waves, and the presence of another psychiatric disorder (iPSYCH); sex, age, county of origin, population stratification, and relatedness (deCODE); sex and five to ten within-ancestry PCs (PGC) | 33096046 |
| | Alcohol dependence | Meta-analysis | European | Overall | 8,485/20,272 | Sex and PCs | 30482948 |

| | | | | | | | |
|---|---|---|---|---|---|---|---|
| | AUDIT (total score) | UKB | European | Overall | 121,604 | Age, sex, genotyping array, and the first 20 PCs | 30336701 |
| | AUDIT (alcohol consumption) | UKB | European | Overall | 121,604 | Age, sex, genotyping array, and the first 20 PCs | 30336701 |
| | AUDIT (alcohol problems) | UKB | European | Overall | 121,604 | Age, sex, genotyping array, and the first 20 PCs | 30336701 |
| Replication outcome | Depression | FinnGen R12 | European | Overall | 59,333/434,831 | Age, sex, FinnGen chip version 1 or 2, legacy genotyping batch, and the first 10 PCs | 36653562 |
| | Bipolar disorder | FinnGen R12 | European | Overall | 8,946/434,831 | Age, sex, FinnGen chip version 1 or 2, legacy genotyping batch, and the first 10 PCs | 36653562 |
| | PTSD | FinnGen R12 | European | Overall | 3,444/444,414 | Age, sex, FinnGen chip version 1 or 2, legacy genotyping batch, and the first 10 PCs | 36653562 |
| | Schizophrenia | FinnGen R12 | European | Overall | 7,234/484,776 | Age, sex, FinnGen chip version 1 or 2, legacy genotyping batch, and the first 10 PCs | 36653562 |
| | Anorexia nervosa (including atypical) | FinnGen R12 | European | Overall | 2,621/484,901 | Age, sex, FinnGen chip version 1 or 2, legacy genotyping batch, and the first 10 PCs | 36653562 |
| | Attention deficit hyperactivity disorder | FinnGen R12 | European | Overall | 4,452/490,708 | Age, sex, FinnGen chip version 1 or 2, legacy genotyping batch, and the first 10 PCs | 36653562 |
| | Substance abuse | FinnGen R12 | European | Overall | 30,806/362,304 | Age, sex, FinnGen chip version 1 or 2, legacy genotyping batch, and the first 10 PCs | 36653562 |

| | | | | | | |
|---|---|---|---|---|---|---|
| Alcohol dependence | FinnGen R12 | European | Overall | 12,599/468,475 | Age, sex, FinnGen chip version 1 or 2, legacy genotyping batch, and the first 10 PCs | 36653562 |

AUDIT, Alcohol Use Disorders Identification Test; CARDIoGRAMplusC4D, Coronary ARtery DIsease Genome wide Replication and Meta-analysis plus The Coronary Artery Disease Genetics; GIANT, Genetic Investigation of ANthropometric Traits; HbA1c, glycated hemoglobin; iPSYCH, Integrative Psychiatric Research; PC, principal component; PGC, Psychiatric Genomics Consortium; PTSD, post-traumatic stress disorder; UKB, UK Biobank; WGS, whole genome sequencing.

**eTable 2. SNP-specific estimates for genetic instruments for GLP-1R activation on BMI (kg/m$^2$) and HbA1c (mmol/mol).**

| Category | Phenotype | Sex | SNP | Effect allele | Other allele | Effect allele frequency | Beta | SE | *P* value | F-statistic |
|---|---|---|---|---|---|---|---|---|---|---|
| Primary analyses | BMI | Overall | rs17757975 | T | C | 0.846 | 0.075 | 0.012 | 1.6E-10 | 41.7 |
| | BMI | Overall | rs4714290 | T | C | 0.705 | 0.054 | 0.009 | 2.4E-09 | 34.7 |
| | BMI | Women | rs17757975 | T | C | 0.845 | 0.066 | 0.016 | 2.3E-05 | 17.5 |
| | BMI | Women | rs4714290 | T | C | 0.705 | 0.056 | 0.012 | 4.6E-06 | 21.5 |
| | BMI | Men | rs17757975 | T | C | 0.844 | 0.089 | 0.017 | 1.8E-07 | 26.4 |
| | BMI | Men | rs4714290 | T | C | 0.704 | 0.058 | 0.013 | 1.1E-05 | 18.7 |
| | HbA1c | Overall | rs10305518 | G | T | 0.057 | 0.195 | 0.034 | 7.2E-09 | 33.5 |
| | HbA1c | Women | rs10305518 | G | T | 0.057 | 0.174 | 0.045 | 1.1E-04 | 14.9 |
| | HbA1c | Men | rs10305518 | G | T | 0.057 | 0.218 | 0.051 | 1.6E-05 | 18.6 |
| Sensitivity analyses | BMI | Overall | rs17757975 | T | C | 0.846 | 0.075 | 0.012 | 1.6E-10 | 41.7 |
| | BMI | Overall | rs4714290 | T | C | 0.705 | 0.054 | 0.009 | 2.4E-09 | 34.7 |
| | BMI | Overall | rs9394581 | A | G | 0.177 | 0.048 | 0.011 | 5.5E-06 | 20.3 |
| | BMI | Women | rs17757975 | T | C | 0.845 | 0.066 | 0.016 | 2.3E-05 | 17.5 |
| | BMI | Women | rs4714290 | T | C | 0.705 | 0.056 | 0.012 | 4.6E-06 | 21.5 |
| | BMI | Women | rs9394581 | A | G | 0.175 | 0.028 | 0.014 | 4.6E-02 | 3.9 |
| | BMI | Men | rs17757975 | T | C | 0.844 | 0.089 | 0.017 | 1.8E-07 | 26.4 |
| | BMI | Men | rs4714290 | T | C | 0.704 | 0.058 | 0.013 | 1.1E-05 | 18.7 |
| | BMI | Men | rs9394581 | A | G | 0.176 | 0.068 | 0.015 | 1.1E-05 | 19.4 |
| | HbA1c | Overall | rs10305518 | G | T | 0.057 | 0.195 | 0.034 | 7.2E-09 | 33.5 |
| | HbA1c | Overall | rs112385083 | C | T | 0.023 | 0.239 | 0.052 | 4.1E-06 | 21.2 |
| | HbA1c | Women | rs10305518 | G | T | 0.057 | 0.174 | 0.045 | 1.1E-04 | 14.9 |
| | HbA1c | Women | rs112385083 | C | T | 0.023 | 0.297 | 0.069 | 1.8E-05 | 18.4 |
| | HbA1c | Men | rs10305518 | G | T | 0.057 | 0.218 | 0.051 | 1.6E-05 | 18.6 |
| | HbA1c | Men | rs112385083 | C | T | 0.023 | 0.177 | 0.078 | 2.3E-02 | 5.2 |

BMI, body mass index; GLP-1R, glucagon-like peptide-1 receptor; HbA1c, glycated hemoglobin.

**eTable 3. MR estimates for the associations of GLP-1R activation with HbA1c (mmol/mol) and BMI (kg/m$^2$).**

| Exposure | Outcome | SNPs | Method | Beta (95% CI) | *P* value | *P* value (intercept) | *P* value (Q) |
|---|---|---|---|---|---|---|---|
| Lower BMI via GLP-1R activation | HbA1c | 2 | IVW | -0.32 (-0.74 to 0.11) | 0.14 | | 0.10 |
| Lower BMI via GLP-1R activation | HbA1c | 3 | IVW | -0.19 (-0.56 to 0.19) | 0.33 | | 0.11 |
| Lower BMI via GLP-1R activation | HbA1c | 3 | Weighted median | -0.13 (-0.62 to 0.36) | 0.60 | | |
| Lower BMI via GLP-1R activation | HbA1c | 3 | MR Egger | 0.12 (-4.18 to 4.42) | 0.96 | 0.89 | |
| Lower BMI via GLP-1R activation | HbA1c | 3 | MR-RAPS | -0.15 (-0.55 to 0.25) | 0.46 | | |
| Lower HbA1c via GLP-1R activation | BMI | 1 | IVW | -0.04 (-0.22 to 0.14) | 0.67 | | |
| Lower HbA1c via GLP-1R activation | BMI | 2 | IVW | 0.05 (-0.07 to 0.18) | 0.40 | | 0.14 |

BMI, body mass index; GLP-1R, glucagon-like peptide-1 receptor; HbA1c, glycated hemoglobin; IVW, inverse variance weighted; MR, Mendelian randomization; RAPS, robust adjusted profile score.

**eTable 4. SNP-specific estimates for genetic instruments for BMI (kg/m$^2$) and HbA1c (mmol/mol).**

| Phenotype | Sex | SNP | Effect allele | Other allele | Effect allele frequency | Beta | SE | *P* value | F-statistic |
|---|---|---|---|---|---|---|---|---|---|
| BMI | Overall | rs2803316 | A | G | 0.455 | -0.068 | 0.008 | 1.9E-16 | 69.8 |
| BMI | Overall | rs7535528 | A | G | 0.372 | -0.074 | 0.009 | 1.6E-17 | 73.2 |
| BMI | Overall | rs6577584 | T | G | 0.659 | -0.057 | 0.009 | 3.4E-11 | 43.0 |
| BMI | Overall | rs2791653 | A | G | 0.240 | 0.064 | 0.009 | 1.0E-12 | 49.0 |
| BMI | Overall | rs9435739 | A | G | 0.661 | -0.052 | 0.009 | 9.9E-10 | 36.7 |
| BMI | Overall | rs4655141 | T | C | 0.836 | -0.083 | 0.011 | 6.2E-15 | 61.8 |
| BMI | Overall | rs10798888 | T | G | 0.189 | 0.062 | 0.011 | 5.8E-09 | 34.9 |
| BMI | Overall | rs12022461 | A | G | 0.167 | -0.080 | 0.011 | 9.8E-14 | 57.6 |
| BMI | Overall | rs7512146 | T | G | 0.527 | -0.047 | 0.008 | 1.3E-08 | 32.6 |
| BMI | Overall | rs11586036 | A | C | 0.935 | -0.095 | 0.017 | 1.1E-08 | 32.0 |
| BMI | Overall | rs11206436 | T | C | 0.893 | 0.071 | 0.013 | 4.7E-08 | 29.6 |
| BMI | Overall | rs1707322 | A | G | 0.316 | -0.065 | 0.009 | 1.6E-14 | 57.1 |
| BMI | Overall | rs2984618 | T | G | 0.441 | 0.079 | 0.008 | 3.8E-24 | 106.3 |
| BMI | Overall | rs657452 | A | G | 0.404 | 0.090 | 0.008 | 3.2E-30 | 138.1 |
| BMI | Overall | rs17425707 | T | C | 0.900 | -0.083 | 0.013 | 6.2E-10 | 38.2 |
| BMI | Overall | rs2481665 | T | C | 0.582 | 0.077 | 0.008 | 3.2E-23 | 101.3 |
| BMI | Overall | rs2503185 | A | G | 0.510 | 0.062 | 0.008 | 1.3E-14 | 58.5 |
| BMI | Overall | rs3101336 | T | C | 0.383 | -0.122 | 0.008 | 4.8E-54 | 252.0 |
| BMI | Overall | rs12036473 | A | T | 0.618 | -0.082 | 0.008 | 2.8E-24 | 101.2 |
| BMI | Overall | rs12049202 | T | C | 0.188 | 0.115 | 0.010 | 3.3E-29 | 129.5 |
| BMI | Overall | rs2166171 | T | C | 0.624 | -0.050 | 0.009 | 5.5E-09 | 32.7 |
| BMI | Overall | rs12034762 | T | C | 0.391 | 0.060 | 0.008 | 6.3E-13 | 54.1 |
| BMI | Overall | rs11165643 | T | C | 0.580 | 0.089 | 0.008 | 4.5E-30 | 133.7 |
| BMI | Overall | rs10747488 | A | C | 0.780 | -0.064 | 0.010 | 1.5E-11 | 44.9 |
| BMI | Overall | rs17024393 | T | C | 0.969 | -0.310 | 0.024 | 7.1E-39 | 172.7 |
| BMI | Overall | rs197374 | T | C | 0.403 | 0.068 | 0.008 | 3.2E-16 | 68.8 |
| BMI | Overall | rs12120851 | T | C | 0.631 | -0.056 | 0.010 | 2.9E-08 | 30.5 |
| BMI | Overall | rs7534091 | A | G | 0.718 | -0.058 | 0.009 | 6.7E-11 | 44.4 |
| BMI | Overall | rs6587552 | A | G | 0.241 | 0.075 | 0.010 | 2.2E-15 | 61.6 |
| BMI | Overall | rs3856261 | A | G | 0.498 | 0.056 | 0.008 | 1.1E-11 | 46.6 |
| BMI | Overall | rs1750307 | A | T | 0.366 | 0.062 | 0.009 | 2.8E-13 | 51.4 |
| BMI | Overall | rs10733051 | A | G | 0.512 | 0.045 | 0.008 | 7.0E-09 | 33.8 |
| BMI | Overall | rs12564992 | A | G | 0.888 | -0.091 | 0.012 | 2.6E-13 | 52.8 |
| BMI | Overall | rs543874 | A | G | 0.779 | -0.230 | 0.010 | 3.1E-125 | 573.6 |
| BMI | Overall | rs10920678 | A | G | 0.425 | 0.072 | 0.008 | 7.1E-20 | 86.7 |
| BMI | Overall | rs2400414 | T | C | 0.346 | -0.061 | 0.009 | 5.5E-13 | 49.0 |
| BMI | Overall | rs2820295 | A | G | 0.327 | 0.113 | 0.009 | 5.6E-39 | 170.4 |
| BMI | Overall | rs10920336 | A | G | 0.533 | -0.049 | 0.008 | 4.2E-09 | 35.3 |
| BMI | Overall | rs1006317 | T | G | 0.127 | 0.077 | 0.012 | 3.7E-10 | 37.9 |
| BMI | Overall | rs6661316 | T | C | 0.589 | 0.058 | 0.008 | 1.7E-13 | 56.3 |
| BMI | Overall | rs10489615 | A | G | 0.400 | 0.048 | 0.008 | 1.3E-08 | 33.9 |
| BMI | Overall | rs946824 | T | C | 0.136 | 0.095 | 0.012 | 4.8E-15 | 62.1 |
| BMI | Overall | rs4577313 | A | G | 0.241 | 0.076 | 0.010 | 2.6E-15 | 62.4 |
| BMI | Overall | rs13021737 | A | G | 0.157 | -0.278 | 0.010 | 2.9E-161 | 757.6 |
| BMI | Overall | rs10929925 | A | C | 0.411 | -0.068 | 0.008 | 3.0E-18 | 78.8 |

| | | | | | | | | | |
|---|---|---|---|---|---|---|---|---|---|
| BMI | Overall | rs11902450 | T | C | 0.105 | 0.080 | 0.013 | 1.1E-09 | 38.3 |
| BMI | Overall | rs10182181 | A | G | 0.505 | -0.157 | 0.008 | 2.5E-91 | 417.7 |
| BMI | Overall | rs1260326 | T | C | 0.391 | -0.051 | 0.008 | 1.2E-10 | 39.6 |
| BMI | Overall | rs17327461 | T | C | 0.446 | 0.059 | 0.008 | 7.6E-14 | 58.1 |
| BMI | Overall | rs10185199 | A | G | 0.286 | -0.069 | 0.010 | 3.2E-13 | 51.1 |
| BMI | Overall | rs10169594 | T | C | 0.648 | -0.058 | 0.009 | 1.3E-11 | 44.4 |
| BMI | Overall | rs7561278 | T | C | 0.767 | 0.081 | 0.010 | 4.9E-16 | 64.8 |
| BMI | Overall | rs930295 | A | C | 0.155 | 0.100 | 0.011 | 2.0E-19 | 81.8 |
| BMI | Overall | rs805412 | A | G | 0.434 | -0.047 | 0.008 | 1.1E-08 | 33.2 |
| BMI | Overall | rs4671328 | T | G | 0.450 | 0.103 | 0.008 | 3.4E-36 | 158.5 |
| BMI | Overall | rs6545714 | A | G | 0.611 | -0.093 | 0.008 | 4.0E-32 | 147.0 |
| BMI | Overall | rs2861683 | A | C | 0.597 | 0.068 | 0.008 | 1.9E-16 | 68.8 |
| BMI | Overall | rs17020497 | A | G | 0.134 | 0.067 | 0.012 | 2.0E-08 | 31.4 |
| BMI | Overall | rs12714199 | T | C | 0.615 | -0.068 | 0.008 | 3.2E-16 | 68.8 |
| BMI | Overall | rs4303732 | T | C | 0.614 | 0.081 | 0.008 | 1.3E-22 | 98.8 |
| BMI | Overall | rs264941 | A | C | 0.468 | -0.060 | 0.008 | 1.8E-13 | 53.2 |
| BMI | Overall | rs10197031 | T | C | 0.726 | -0.077 | 0.009 | 5.0E-18 | 71.8 |
| BMI | Overall | rs11884795 | A | G | 0.268 | 0.054 | 0.009 | 1.6E-09 | 38.7 |
| BMI | Overall | rs4988235 | A | G | 0.721 | 0.060 | 0.008 | 7.1E-13 | 53.2 |
| BMI | Overall | rs17814208 | A | G | 0.758 | -0.062 | 0.010 | 7.2E-11 | 41.6 |
| BMI | Overall | rs16824165 | A | G | 0.116 | 0.077 | 0.013 | 2.2E-09 | 35.1 |
| BMI | Overall | rs429343 | A | G | 0.434 | 0.070 | 0.008 | 8.4E-18 | 73.8 |
| BMI | Overall | rs2119753 | A | G | 0.623 | 0.048 | 0.008 | 1.0E-08 | 34.6 |
| BMI | Overall | rs3764835 | A | G | 0.151 | -0.062 | 0.012 | 3.8E-08 | 29.3 |
| BMI | Overall | rs12692596 | T | C | 0.362 | 0.058 | 0.008 | 1.0E-12 | 49.8 |
| BMI | Overall | rs1521527 | C | G | 0.518 | -0.049 | 0.008 | 1.8E-09 | 36.0 |
| BMI | Overall | rs3754963 | A | T | 0.763 | 0.062 | 0.009 | 2.4E-11 | 46.1 |
| BMI | Overall | rs2138348 | T | G | 0.284 | -0.064 | 0.009 | 1.4E-12 | 49.7 |
| BMI | Overall | rs2119137 | A | G | 0.662 | -0.063 | 0.009 | 2.3E-13 | 53.0 |
| BMI | Overall | rs7588437 | A | G | 0.366 | -0.079 | 0.008 | 2.3E-22 | 94.2 |
| BMI | Overall | rs6757852 | A | G | 0.499 | 0.051 | 0.008 | 3.8E-10 | 38.9 |
| BMI | Overall | rs7593917 | A | G | 0.454 | -0.055 | 0.008 | 9.7E-13 | 51.7 |
| BMI | Overall | rs4482463 | A | C | 0.917 | -0.149 | 0.015 | 4.8E-23 | 100.0 |
| BMI | Overall | rs6715020 | T | C | 0.381 | -0.050 | 0.008 | 1.3E-09 | 38.1 |
| BMI | Overall | rs11692326 | T | C | 0.230 | 0.071 | 0.009 | 1.7E-14 | 59.9 |
| BMI | Overall | rs7599312 | A | G | 0.274 | -0.087 | 0.009 | 1.5E-23 | 102.2 |
| BMI | Overall | rs12987009 | A | T | 0.585 | -0.052 | 0.008 | 1.8E-10 | 41.1 |
| BMI | Overall | rs6725931 | T | C | 0.848 | 0.090 | 0.012 | 2.3E-15 | 60.7 |
| BMI | Overall | rs4973618 | A | G | 0.662 | -0.071 | 0.009 | 1.3E-16 | 67.6 |
| BMI | Overall | rs6720868 | T | C | 0.308 | 0.074 | 0.009 | 1.8E-17 | 73.2 |
| BMI | Overall | rs7568228 | C | G | 0.540 | -0.050 | 0.008 | 1.0E-09 | 37.4 |
| BMI | Overall | rs17535749 | A | G | 0.108 | 0.076 | 0.013 | 3.5E-09 | 34.7 |
| BMI | Overall | rs10510419 | T | G | 0.147 | -0.081 | 0.011 | 2.2E-13 | 53.4 |
| BMI | Overall | rs2600226 | T | C | 0.671 | -0.056 | 0.009 | 1.4E-10 | 41.5 |
| BMI | Overall | rs1048637 | T | G | 0.547 | -0.045 | 0.008 | 2.9E-08 | 30.6 |
| BMI | Overall | rs4858193 | T | C | 0.720 | 0.064 | 0.009 | 2.4E-12 | 49.0 |
| BMI | Overall | rs6804842 | A | G | 0.430 | -0.068 | 0.008 | 7.6E-18 | 77.7 |
| BMI | Overall | rs11921432 | T | C | 0.891 | -0.091 | 0.013 | 5.4E-12 | 49.0 |

| | | | | | | | | | |
|---|---|---|---|---|---|---|---|---|---|
| BMI | Overall | rs1799923 | A | G | 0.112 | -0.108 | 0.012 | 1.1E-17 | 74.2 |
| BMI | Overall | rs28350 | A | G | 0.172 | 0.083 | 0.011 | 1.1E-14 | 61.1 |
| BMI | Overall | rs4017425 | T | C | 0.468 | -0.057 | 0.008 | 2.9E-12 | 48.2 |
| BMI | Overall | rs11713193 | A | G | 0.516 | 0.118 | 0.008 | 3.0E-48 | 209.4 |
| BMI | Overall | rs2365389 | T | C | 0.400 | -0.081 | 0.008 | 6.5E-25 | 110.3 |
| BMI | Overall | rs925018 | C | G | 0.667 | -0.062 | 0.008 | 4.3E-14 | 58.5 |
| BMI | Overall | rs11915371 | A | C | 0.802 | -0.074 | 0.010 | 2.3E-13 | 53.8 |
| BMI | Overall | rs12636480 | T | G | 0.351 | 0.062 | 0.009 | 7.0E-13 | 50.6 |
| BMI | Overall | rs9827823 | T | C | 0.855 | 0.087 | 0.011 | 3.4E-15 | 62.6 |
| BMI | Overall | rs9818122 | T | C | 0.793 | -0.110 | 0.010 | 4.0E-30 | 130.0 |
| BMI | Overall | rs11128021 | A | G | 0.162 | -0.087 | 0.012 | 1.6E-14 | 56.9 |
| BMI | Overall | rs1492014 | T | C | 0.567 | -0.082 | 0.008 | 1.4E-23 | 101.2 |
| BMI | Overall | rs1436344 | C | G | 0.570 | 0.071 | 0.008 | 1.1E-17 | 74.8 |
| BMI | Overall | rs7640424 | T | C | 0.311 | -0.065 | 0.009 | 1.2E-14 | 56.3 |
| BMI | Overall | rs17681451 | A | G | 0.076 | -0.108 | 0.015 | 7.4E-13 | 52.7 |
| BMI | Overall | rs6808814 | T | C | 0.734 | 0.057 | 0.009 | 1.3E-09 | 38.6 |
| BMI | Overall | rs7631156 | A | G | 0.309 | 0.103 | 0.009 | 3.3E-32 | 142.7 |
| BMI | Overall | rs687339 | T | C | 0.770 | 0.090 | 0.009 | 4.3E-22 | 97.9 |
| BMI | Overall | rs16851483 | T | G | 0.072 | 0.169 | 0.016 | 4.9E-25 | 107.2 |
| BMI | Overall | rs355777 | C | G | 0.398 | 0.073 | 0.008 | 2.1E-18 | 78.9 |
| BMI | Overall | rs9826775 | A | G | 0.851 | 0.075 | 0.012 | 6.6E-11 | 41.7 |
| BMI | Overall | rs2047648 | A | T | 0.747 | -0.064 | 0.010 | 7.9E-12 | 44.2 |
| BMI | Overall | rs3732927 | T | C | 0.291 | 0.048 | 0.008 | 1.1E-08 | 33.9 |
| BMI | Overall | rs12635553 | A | T | 0.486 | 0.048 | 0.008 | 4.1E-09 | 33.9 |
| BMI | Overall | rs39654 | A | G | 0.451 | -0.078 | 0.008 | 1.7E-21 | 91.9 |
| BMI | Overall | rs6443750 | T | C | 0.199 | -0.073 | 0.010 | 7.3E-13 | 52.4 |
| BMI | Overall | rs865809 | A | G | 0.225 | 0.060 | 0.010 | 6.5E-10 | 38.4 |
| BMI | Overall | rs9816226 | A | T | 0.176 | -0.151 | 0.010 | 1.4E-50 | 225.0 |
| BMI | Overall | rs7616009 | A | G | 0.160 | -0.075 | 0.012 | 4.3E-11 | 42.8 |
| BMI | Overall | rs2051559 | T | C | 0.859 | -0.080 | 0.012 | 3.8E-11 | 44.6 |
| BMI | Overall | rs6818414 | T | C | 0.516 | -0.048 | 0.008 | 4.6E-09 | 34.6 |
| BMI | Overall | rs1477890 | A | G | 0.488 | -0.062 | 0.008 | 6.1E-14 | 56.7 |
| BMI | Overall | rs9291467 | T | C | 0.465 | 0.068 | 0.008 | 4.6E-17 | 69.8 |
| BMI | Overall | rs6448587 | A | C | 0.806 | 0.075 | 0.011 | 1.6E-12 | 50.3 |
| BMI | Overall | rs337637 | A | G | 0.359 | -0.066 | 0.008 | 6.2E-16 | 64.9 |
| BMI | Overall | rs10938397 | A | G | 0.570 | -0.155 | 0.008 | 2.4E-86 | 405.0 |
| BMI | Overall | rs2271046 | A | T | 0.695 | -0.055 | 0.009 | 3.0E-10 | 40.8 |
| BMI | Overall | rs1492767 | T | C | 0.468 | 0.046 | 0.008 | 3.5E-09 | 35.3 |
| BMI | Overall | rs2192158 | A | G | 0.449 | 0.066 | 0.008 | 7.4E-16 | 64.9 |
| BMI | Overall | rs1346841 | A | G | 0.412 | -0.061 | 0.008 | 3.2E-13 | 54.9 |
| BMI | Overall | rs10002111 | A | G | 0.223 | 0.060 | 0.010 | 1.3E-09 | 35.4 |
| BMI | Overall | rs10033843 | A | G | 0.215 | 0.067 | 0.010 | 1.1E-11 | 44.4 |
| BMI | Overall | rs17507682 | T | G | 0.154 | 0.065 | 0.012 | 1.9E-08 | 31.6 |
| BMI | Overall | rs4148155 | A | G | 0.887 | 0.092 | 0.012 | 1.3E-13 | 54.5 |
| BMI | Overall | rs4286488 | A | G | 0.761 | 0.058 | 0.010 | 1.7E-09 | 36.6 |
| BMI | Overall | rs7678054 | A | G | 0.479 | -0.048 | 0.008 | 4.6E-09 | 33.9 |
| BMI | Overall | rs3796432 | T | G | 0.367 | -0.054 | 0.009 | 2.2E-10 | 39.4 |
| BMI | Overall | rs13107325 | T | C | 0.082 | 0.225 | 0.015 | 3.8E-47 | 213.9 |

| | | | | | | | | | |
|---|---|---|---|---|---|---|---|---|---|
| BMI | Overall | rs326893 | T | C | 0.581 | 0.058 | 0.008 | 1.9E-12 | 50.7 |
| BMI | Overall | rs7696649 | A | G | 0.278 | 0.056 | 0.009 | 4.7E-10 | 37.3 |
| BMI | Overall | rs4864201 | T | C | 0.347 | 0.066 | 0.008 | 4.3E-16 | 64.9 |
| BMI | Overall | rs1296328 | A | C | 0.446 | 0.080 | 0.008 | 3.5E-22 | 95.3 |
| BMI | Overall | rs17367750 | T | C | 0.312 | -0.059 | 0.009 | 2.0E-11 | 45.9 |
| BMI | Overall | rs9992189 | C | G | 0.608 | -0.047 | 0.008 | 2.0E-08 | 32.6 |
| BMI | Overall | rs3914628 | T | C | 0.859 | 0.079 | 0.011 | 6.9E-13 | 51.5 |
| BMI | Overall | rs750090 | T | C | 0.626 | 0.054 | 0.009 | 2.5E-10 | 39.4 |
| BMI | Overall | rs13110266 | A | G | 0.404 | -0.060 | 0.008 | 4.0E-14 | 60.1 |
| BMI | Overall | rs7685628 | A | T | 0.401 | 0.049 | 0.008 | 6.1E-09 | 35.3 |
| BMI | Overall | rs1522569 | T | G | 0.818 | 0.068 | 0.011 | 1.6E-10 | 41.1 |
| BMI | Overall | rs1437842 | A | G | 0.491 | -0.051 | 0.008 | 8.5E-10 | 38.9 |
| BMI | Overall | rs4565118 | A | C | 0.502 | -0.051 | 0.009 | 7.3E-09 | 34.7 |
| BMI | Overall | rs698147 | A | G | 0.447 | 0.056 | 0.008 | 9.7E-12 | 46.6 |
| BMI | Overall | rs6890310 | A | G | 0.292 | -0.057 | 0.009 | 3.3E-10 | 39.2 |
| BMI | Overall | rs7730004 | T | C | 0.661 | 0.067 | 0.009 | 1.5E-14 | 59.6 |
| BMI | Overall | rs7704281 | A | G | 0.046 | 0.119 | 0.020 | 1.2E-09 | 36.6 |
| BMI | Overall | rs13186194 | T | C | 0.617 | 0.048 | 0.008 | 2.2E-09 | 34.6 |
| BMI | Overall | rs4700608 | T | C | 0.524 | -0.075 | 0.008 | 4.3E-20 | 83.1 |
| BMI | Overall | rs6886740 | T | C | 0.662 | 0.055 | 0.009 | 3.1E-10 | 40.1 |
| BMI | Overall | rs1159692 | A | C | 0.475 | 0.065 | 0.008 | 5.5E-15 | 63.1 |
| BMI | Overall | rs2112347 | T | G | 0.630 | 0.133 | 0.008 | 1.2E-61 | 263.6 |
| BMI | Overall | rs10942267 | A | G | 0.693 | 0.071 | 0.009 | 6.5E-16 | 67.6 |
| BMI | Overall | rs2962334 | T | G | 0.027 | 0.190 | 0.028 | 1.8E-11 | 45.0 |
| BMI | Overall | rs1501673 | A | G | 0.136 | 0.139 | 0.012 | 2.7E-31 | 133.6 |
| BMI | Overall | rs12652212 | A | G | 0.561 | -0.063 | 0.008 | 1.6E-15 | 67.0 |
| BMI | Overall | rs7713317 | A | G | 0.720 | -0.080 | 0.009 | 2.0E-20 | 85.0 |
| BMI | Overall | rs6882366 | T | C | 0.396 | -0.063 | 0.008 | 4.4E-14 | 59.4 |
| BMI | Overall | rs11739877 | T | C | 0.623 | 0.056 | 0.009 | 4.0E-11 | 41.5 |
| BMI | Overall | rs6888194 | T | C | 0.844 | -0.061 | 0.011 | 3.6E-08 | 30.5 |
| BMI | Overall | rs40067 | A | G | 0.170 | -0.121 | 0.011 | 9.7E-29 | 120.0 |
| BMI | Overall | rs459552 | A | T | 0.775 | -0.064 | 0.009 | 8.4E-12 | 49.0 |
| BMI | Overall | rs6893539 | A | C | 0.698 | -0.059 | 0.009 | 6.2E-11 | 41.2 |
| BMI | Overall | rs6864049 | A | G | 0.489 | -0.058 | 0.008 | 1.5E-13 | 57.2 |
| BMI | Overall | rs6886072 | T | C | 0.467 | -0.048 | 0.008 | 4.1E-09 | 34.6 |
| BMI | Overall | rs13174863 | A | G | 0.854 | -0.095 | 0.011 | 1.9E-17 | 73.4 |
| BMI | Overall | rs17405603 | A | T | 0.706 | -0.060 | 0.009 | 4.4E-11 | 43.3 |
| BMI | Overall | rs2973157 | A | G | 0.671 | -0.054 | 0.010 | 4.2E-08 | 28.4 |
| BMI | Overall | rs7715256 | T | G | 0.564 | -0.076 | 0.008 | 4.0E-22 | 97.5 |
| BMI | Overall | rs7734385 | A | G | 0.445 | -0.049 | 0.008 | 6.1E-10 | 39.8 |
| BMI | Overall | rs2861089 | A | T | 0.381 | 0.050 | 0.008 | 1.5E-09 | 38.1 |
| BMI | Overall | rs7727781 | T | C | 0.516 | 0.045 | 0.008 | 4.3E-08 | 29.9 |
| BMI | Overall | rs2053682 | A | C | 0.678 | 0.082 | 0.009 | 2.6E-20 | 89.2 |
| BMI | Overall | rs6556301 | T | G | 0.371 | -0.054 | 0.008 | 8.1E-11 | 44.2 |
| BMI | Overall | rs2228213 | A | G | 0.337 | -0.069 | 0.008 | 5.5E-17 | 71.8 |
| BMI | Overall | rs11757278 | T | C | 0.696 | 0.064 | 0.009 | 6.9E-13 | 49.0 |
| BMI | Overall | rs3806114 | A | G | 0.692 | -0.058 | 0.009 | 1.5E-11 | 44.4 |
| BMI | Overall | rs2066295 | A | G | 0.758 | 0.068 | 0.010 | 2.3E-12 | 50.4 |

| | | | | | | | | | |
|---|---|---|---|---|---|---|---|---|---|
| BMI | Overall | rs3115667 | T | C | 0.282 | -0.086 | 0.009 | 5.9E-21 | 88.8 |
| BMI | Overall | rs2281819 | A | T | 0.229 | -0.074 | 0.010 | 2.1E-14 | 59.3 |
| BMI | Overall | rs2744974 | T | C | 0.324 | 0.125 | 0.008 | 1.3E-51 | 235.7 |
| BMI | Overall | rs2436728 | A | G | 0.407 | 0.091 | 0.008 | 2.0E-29 | 123.6 |
| BMI | Overall | rs1358980 | T | C | 0.474 | -0.062 | 0.008 | 5.2E-15 | 57.6 |
| BMI | Overall | rs2206277 | T | C | 0.156 | 0.196 | 0.010 | 1.8E-83 | 377.5 |
| BMI | Overall | rs1327259 | A | G | 0.610 | 0.075 | 0.008 | 1.5E-19 | 85.3 |
| BMI | Overall | rs6915002 | T | C | 0.411 | 0.048 | 0.008 | 8.7E-09 | 33.9 |
| BMI | Overall | rs9370410 | A | G | 0.729 | 0.050 | 0.009 | 2.6E-08 | 30.5 |
| BMI | Overall | rs2622274 | T | G | 0.455 | -0.051 | 0.008 | 3.2E-10 | 39.6 |
| BMI | Overall | rs13213867 | A | G | 0.803 | 0.059 | 0.011 | 1.0E-08 | 31.3 |
| BMI | Overall | rs6921533 | T | C | 0.293 | 0.050 | 0.009 | 1.4E-08 | 33.4 |
| BMI | Overall | rs9294260 | A | G | 0.469 | 0.067 | 0.008 | 8.2E-18 | 76.6 |
| BMI | Overall | rs6909685 | T | C | 0.330 | -0.072 | 0.009 | 2.8E-16 | 68.5 |
| BMI | Overall | rs9320823 | T | C | 0.412 | -0.079 | 0.008 | 2.1E-21 | 94.2 |
| BMI | Overall | rs12530388 | A | C | 0.474 | 0.048 | 0.008 | 4.2E-09 | 34.6 |
| BMI | Overall | rs156201 | C | G | 0.746 | 0.060 | 0.009 | 1.6E-10 | 42.6 |
| BMI | Overall | rs768023 | A | G | 0.603 | 0.077 | 0.008 | 1.1E-22 | 101.3 |
| BMI | Overall | rs2357760 | A | G | 0.674 | 0.069 | 0.008 | 2.1E-16 | 70.8 |
| BMI | Overall | rs2875762 | C | G | 0.252 | 0.062 | 0.010 | 1.1E-10 | 41.6 |
| BMI | Overall | rs1268065 | A | G | 0.500 | -0.048 | 0.008 | 7.1E-10 | 39.1 |
| BMI | Overall | rs9375702 | T | C | 0.679 | -0.051 | 0.009 | 5.5E-09 | 34.7 |
| BMI | Overall | rs2246012 | T | C | 0.851 | -0.077 | 0.011 | 1.2E-13 | 53.6 |
| BMI | Overall | rs6922607 | A | G | 0.808 | -0.062 | 0.011 | 1.9E-09 | 34.9 |
| BMI | Overall | rs765875 | T | C | 0.469 | -0.063 | 0.008 | 1.1E-14 | 60.3 |
| BMI | Overall | rs7760482 | A | G | 0.616 | -0.049 | 0.009 | 6.2E-09 | 32.1 |
| BMI | Overall | rs12527426 | A | G | 0.300 | 0.072 | 0.009 | 5.5E-16 | 62.3 |
| BMI | Overall | rs9478496 | T | C | 0.845 | -0.075 | 0.011 | 8.2E-12 | 46.6 |
| BMI | Overall | rs13191362 | A | G | 0.857 | 0.113 | 0.012 | 4.1E-21 | 88.4 |
| BMI | Overall | rs9458814 | T | C | 0.771 | -0.054 | 0.010 | 2.3E-08 | 31.4 |
| BMI | Overall | rs6461115 | A | G | 0.762 | 0.064 | 0.009 | 1.5E-12 | 49.7 |
| BMI | Overall | rs6463489 | T | C | 0.096 | 0.080 | 0.012 | 2.5E-10 | 41.3 |
| BMI | Overall | rs7779296 | A | G | 0.716 | -0.050 | 0.009 | 4.6E-09 | 32.7 |
| BMI | Overall | rs4307239 | A | G | 0.536 | -0.055 | 0.008 | 1.5E-11 | 45.8 |
| BMI | Overall | rs213518 | T | C | 0.850 | -0.074 | 0.012 | 2.5E-10 | 40.6 |
| BMI | Overall | rs215669 | A | G | 0.602 | -0.072 | 0.008 | 8.9E-18 | 76.8 |
| BMI | Overall | rs2108719 | A | G | 0.727 | 0.051 | 0.009 | 2.3E-08 | 31.7 |
| BMI | Overall | rs217433 | T | C | 0.803 | -0.055 | 0.010 | 3.0E-08 | 30.0 |
| BMI | Overall | rs2289379 | T | C | 0.392 | -0.066 | 0.009 | 7.0E-15 | 57.9 |
| BMI | Overall | rs10499694 | A | G | 0.494 | 0.062 | 0.008 | 1.3E-15 | 66.0 |
| BMI | Overall | rs11772246 | T | C | 0.823 | 0.070 | 0.011 | 3.7E-11 | 43.4 |
| BMI | Overall | rs17207196 | T | C | 0.420 | -0.106 | 0.008 | 1.6E-36 | 167.5 |
| BMI | Overall | rs17149254 | T | C | 0.205 | 0.114 | 0.011 | 3.0E-25 | 107.1 |
| BMI | Overall | rs6973656 | A | G | 0.584 | -0.049 | 0.008 | 6.3E-09 | 35.3 |
| BMI | Overall | rs1965529 | A | G | 0.773 | 0.077 | 0.011 | 7.7E-14 | 53.6 |
| BMI | Overall | rs274628 | A | C | 0.337 | -0.049 | 0.009 | 1.4E-08 | 32.1 |
| BMI | Overall | rs2283006 | A | G | 0.488 | 0.063 | 0.008 | 8.1E-15 | 60.3 |
| BMI | Overall | rs13240600 | A | G | 0.834 | 0.087 | 0.011 | 7.9E-16 | 66.9 |

| | | | | | | | | | |
|---|---|---|---|---|---|---|---|---|---|
| BMI | Overall | rs1721447 | T | G | 0.510 | -0.048 | 0.008 | 3.8E-09 | 34.6 |
| BMI | Overall | rs2396625 | A | T | 0.418 | -0.085 | 0.008 | 2.8E-24 | 107.2 |
| BMI | Overall | rs13245051 | A | G | 0.456 | 0.072 | 0.008 | 1.1E-18 | 77.9 |
| BMI | Overall | rs10261050 | T | C | 0.475 | 0.054 | 0.008 | 4.5E-11 | 44.2 |
| BMI | Overall | rs2283093 | T | C | 0.195 | 0.058 | 0.010 | 1.1E-08 | 33.2 |
| BMI | Overall | rs7802342 | T | G | 0.705 | -0.060 | 0.009 | 6.2E-11 | 42.6 |
| BMI | Overall | rs17160760 | A | T | 0.105 | -0.079 | 0.013 | 1.3E-09 | 37.3 |
| BMI | Overall | rs11773362 | T | C | 0.340 | -0.050 | 0.009 | 6.4E-09 | 34.0 |
| BMI | Overall | rs2907948 | A | G | 0.244 | -0.070 | 0.009 | 1.9E-14 | 58.2 |
| BMI | Overall | rs1700082 | C | G | 0.659 | 0.045 | 0.008 | 3.4E-08 | 30.6 |
| BMI | Overall | rs4240673 | T | C | 0.455 | 0.084 | 0.008 | 1.6E-26 | 119.6 |
| BMI | Overall | rs13263601 | A | C | 0.659 | -0.070 | 0.009 | 4.9E-16 | 64.9 |
| BMI | Overall | rs10110189 | T | C | 0.106 | -0.075 | 0.013 | 2.2E-08 | 31.0 |
| BMI | Overall | rs10101364 | T | C | 0.680 | 0.058 | 0.009 | 5.6E-11 | 44.4 |
| BMI | Overall | rs1421334 | A | C | 0.455 | 0.065 | 0.008 | 3.1E-15 | 63.1 |
| BMI | Overall | rs2466103 | T | G | 0.693 | -0.058 | 0.009 | 8.0E-12 | 45.2 |
| BMI | Overall | rs6468266 | A | T | 0.418 | -0.054 | 0.008 | 7.7E-11 | 44.2 |
| BMI | Overall | rs12681792 | A | C | 0.202 | 0.072 | 0.010 | 2.9E-12 | 51.0 |
| BMI | Overall | rs12334877 | A | G | 0.191 | -0.069 | 0.010 | 2.2E-11 | 47.0 |
| BMI | Overall | rs1431659 | A | G | 0.274 | 0.092 | 0.009 | 2.3E-23 | 101.1 |
| BMI | Overall | rs17405819 | T | C | 0.685 | 0.101 | 0.009 | 6.0E-33 | 137.4 |
| BMI | Overall | rs2196618 | A | G | 0.263 | -0.066 | 0.009 | 1.5E-12 | 52.0 |
| BMI | Overall | rs2120710 | A | G | 0.653 | 0.049 | 0.009 | 2.0E-08 | 31.5 |
| BMI | Overall | rs12680842 | A | G | 0.684 | 0.068 | 0.008 | 3.4E-16 | 69.8 |
| BMI | Overall | rs6469351 | T | C | 0.672 | 0.053 | 0.009 | 3.7E-10 | 38.0 |
| BMI | Overall | rs3808477 | T | C | 0.274 | -0.087 | 0.009 | 8.7E-22 | 91.8 |
| BMI | Overall | rs11781699 | T | C | 0.813 | -0.068 | 0.010 | 1.6E-11 | 45.1 |
| BMI | Overall | rs6470144 | T | G | 0.650 | 0.047 | 0.009 | 3.1E-08 | 29.6 |
| BMI | Overall | rs7842934 | T | C | 0.921 | -0.086 | 0.015 | 1.5E-08 | 33.0 |
| BMI | Overall | rs305256 | T | C | 0.226 | -0.055 | 0.010 | 2.8E-08 | 30.0 |
| BMI | Overall | rs16906838 | T | C | 0.049 | -0.119 | 0.019 | 4.3E-10 | 38.1 |
| BMI | Overall | rs11782074 | T | G | 0.368 | 0.060 | 0.009 | 4.2E-12 | 47.5 |
| BMI | Overall | rs10099330 | A | G | 0.538 | -0.057 | 0.008 | 3.2E-12 | 49.0 |
| BMI | Overall | rs10975933 | C | G | 0.663 | 0.055 | 0.009 | 2.1E-10 | 40.1 |
| BMI | Overall | rs1948080 | T | G | 0.630 | 0.065 | 0.009 | 1.1E-14 | 57.1 |
| BMI | Overall | rs10961649 | T | C | 0.324 | 0.050 | 0.009 | 1.5E-08 | 33.4 |
| BMI | Overall | rs4740619 | T | C | 0.544 | 0.091 | 0.008 | 3.1E-31 | 139.5 |
| BMI | Overall | rs10962550 | C | G | 0.176 | 0.089 | 0.011 | 5.7E-17 | 71.5 |
| BMI | Overall | rs10811868 | A | G | 0.320 | -0.048 | 0.009 | 3.2E-08 | 30.9 |
| BMI | Overall | rs10968114 | A | C | 0.533 | 0.054 | 0.008 | 3.3E-11 | 44.2 |
| BMI | Overall | rs1412235 | C | G | 0.315 | 0.114 | 0.008 | 2.3E-42 | 194.4 |
| BMI | Overall | rs10121187 | C | G | 0.509 | 0.049 | 0.008 | 2.0E-09 | 36.0 |
| BMI | Overall | rs2275003 | A | G | 0.501 | 0.053 | 0.008 | 9.4E-12 | 48.1 |
| BMI | Overall | rs13296413 | T | C | 0.381 | -0.071 | 0.008 | 2.5E-17 | 75.8 |
| BMI | Overall | rs2134858 | T | C | 0.512 | -0.056 | 0.008 | 5.9E-12 | 47.4 |
| BMI | Overall | rs7861160 | T | C | 0.594 | 0.046 | 0.008 | 3.7E-08 | 31.2 |
| BMI | Overall | rs1999433 | T | C | 0.440 | -0.051 | 0.008 | 2.9E-10 | 39.6 |
| BMI | Overall | rs2777768 | A | G | 0.723 | 0.057 | 0.009 | 6.4E-10 | 39.2 |

| | | | | | | | | | |
|---|---|---|---|---|---|---|---|---|---|
| BMI | Overall | rs7357754 | A | G | 0.499 | -0.058 | 0.008 | 1.8E-12 | 49.8 |
| BMI | Overall | rs10992867 | A | G | 0.269 | 0.078 | 0.009 | 3.2E-17 | 72.7 |
| BMI | Overall | rs7025938 | C | G | 0.680 | -0.078 | 0.009 | 1.5E-19 | 81.0 |
| BMI | Overall | rs7024334 | T | G | 0.225 | 0.065 | 0.010 | 4.7E-12 | 45.6 |
| BMI | Overall | rs1928295 | T | C | 0.571 | 0.064 | 0.008 | 2.2E-16 | 70.1 |
| BMI | Overall | rs1877875 | T | C | 0.434 | -0.052 | 0.008 | 2.5E-10 | 41.1 |
| BMI | Overall | rs10733682 | A | G | 0.459 | 0.071 | 0.008 | 1.8E-19 | 85.6 |
| BMI | Overall | rs2267958 | A | G | 0.509 | -0.062 | 0.009 | 2.0E-13 | 52.2 |
| BMI | Overall | rs4740383 | A | G | 0.419 | 0.063 | 0.009 | 7.0E-14 | 53.0 |
| BMI | Overall | rs10858334 | C | G | 0.848 | -0.071 | 0.012 | 4.9E-09 | 35.0 |
| BMI | Overall | rs7907470 | A | G | 0.916 | -0.085 | 0.015 | 7.4E-09 | 32.6 |
| BMI | Overall | rs7893571 | T | G | 0.674 | 0.060 | 0.009 | 5.8E-12 | 48.2 |
| BMI | Overall | rs7084454 | A | G | 0.309 | 0.095 | 0.009 | 4.5E-27 | 121.0 |
| BMI | Overall | rs10829164 | T | C | 0.147 | 0.073 | 0.012 | 2.4E-10 | 39.6 |
| BMI | Overall | rs4097319 | T | G | 0.564 | 0.051 | 0.008 | 4.3E-10 | 39.6 |
| BMI | Overall | rs12765914 | T | C | 0.080 | 0.109 | 0.015 | 2.0E-13 | 53.1 |
| BMI | Overall | rs1624134 | C | G | 0.393 | 0.048 | 0.008 | 6.5E-09 | 34.6 |
| BMI | Overall | rs12259464 | A | G | 0.481 | 0.052 | 0.008 | 1.5E-10 | 41.1 |
| BMI | Overall | rs10761785 | T | G | 0.514 | -0.064 | 0.008 | 3.5E-16 | 69.1 |
| BMI | Overall | rs12098284 | T | C | 0.121 | 0.088 | 0.012 | 9.9E-13 | 50.1 |
| BMI | Overall | rs11001259 | A | T | 0.177 | -0.066 | 0.012 | 1.2E-08 | 33.1 |
| BMI | Overall | rs7899106 | A | G | 0.950 | -0.157 | 0.018 | 1.7E-18 | 78.1 |
| BMI | Overall | rs10788494 | C | G | 0.484 | 0.063 | 0.008 | 5.5E-15 | 60.3 |
| BMI | Overall | rs2439823 | A | G | 0.451 | -0.079 | 0.008 | 6.5E-22 | 94.2 |
| BMI | Overall | rs17094222 | T | C | 0.791 | -0.083 | 0.010 | 4.0E-18 | 74.8 |
| BMI | Overall | rs1886276 | A | G | 0.571 | -0.052 | 0.008 | 3.0E-10 | 40.4 |
| BMI | Overall | rs7903146 | T | C | 0.279 | -0.086 | 0.009 | 1.7E-23 | 97.8 |
| BMI | Overall | rs2257791 | A | G | 0.756 | -0.067 | 0.010 | 1.6E-12 | 48.3 |
| BMI | Overall | rs845084 | A | G | 0.267 | 0.065 | 0.009 | 3.2E-12 | 51.2 |
| BMI | Overall | rs17636031 | T | C | 0.724 | -0.074 | 0.009 | 3.9E-17 | 73.2 |
| BMI | Overall | rs4880341 | T | C | 0.573 | -0.062 | 0.008 | 3.1E-14 | 58.5 |
| BMI | Overall | rs12416812 | A | G | 0.507 | 0.051 | 0.008 | 5.2E-11 | 43.9 |
| BMI | Overall | rs4256980 | C | G | 0.331 | -0.090 | 0.008 | 8.6E-29 | 121.0 |
| BMI | Overall | rs900144 | T | C | 0.578 | 0.071 | 0.008 | 1.5E-18 | 75.8 |
| BMI | Overall | rs2074314 | T | C | 0.636 | 0.050 | 0.008 | 1.4E-09 | 38.1 |
| BMI | Overall | rs6265 | T | C | 0.184 | -0.199 | 0.010 | 7.4E-89 | 386.8 |
| BMI | Overall | rs570463 | A | C | 0.327 | -0.058 | 0.009 | 4.3E-11 | 45.2 |
| BMI | Overall | rs11030618 | T | C | 0.579 | 0.053 | 0.008 | 1.7E-10 | 41.9 |
| BMI | Overall | rs2065418 | T | G | 0.646 | 0.067 | 0.009 | 6.1E-15 | 59.6 |
| BMI | Overall | rs2862996 | T | G | 0.701 | -0.104 | 0.008 | 3.6E-35 | 161.4 |
| BMI | Overall | rs10838202 | C | G | 0.556 | 0.054 | 0.008 | 5.5E-11 | 44.2 |
| BMI | Overall | rs10742752 | T | C | 0.376 | -0.059 | 0.008 | 1.2E-13 | 52.3 |
| BMI | Overall | rs7124681 | A | C | 0.416 | 0.124 | 0.008 | 4.0E-55 | 258.0 |
| BMI | Overall | rs6591407 | A | C | 0.198 | -0.060 | 0.010 | 3.6E-09 | 34.9 |
| BMI | Overall | rs562664 | T | C | 0.187 | -0.066 | 0.011 | 4.2E-10 | 39.3 |
| BMI | Overall | rs7102454 | T | C | 0.644 | -0.081 | 0.009 | 3.8E-21 | 87.1 |
| BMI | Overall | rs592483 | T | C | 0.593 | -0.066 | 0.008 | 1.5E-16 | 64.9 |
| BMI | Overall | rs737185 | A | G | 0.781 | 0.066 | 0.010 | 4.2E-11 | 42.6 |

| | | | | | | | | | |
|---|---|---|---|---|---|---|---|---|---|
| BMI | Overall | rs349088 | A | C | 0.478 | -0.062 | 0.008 | 3.5E-14 | 58.5 |
| BMI | Overall | rs10741329 | A | G | 0.694 | 0.055 | 0.009 | 3.9E-10 | 40.8 |
| BMI | Overall | rs3019466 | T | C | 0.168 | -0.062 | 0.012 | 4.9E-08 | 28.4 |
| BMI | Overall | rs2605603 | A | G | 0.481 | -0.050 | 0.008 | 2.0E-10 | 41.4 |
| BMI | Overall | rs4937870 | A | G | 0.694 | 0.054 | 0.009 | 1.6E-09 | 34.7 |
| BMI | Overall | rs10891549 | T | C | 0.465 | 0.046 | 0.008 | 1.9E-08 | 31.2 |
| BMI | Overall | rs12286929 | A | G | 0.501 | -0.085 | 0.008 | 1.9E-27 | 122.4 |
| BMI | Overall | rs3825061 | T | C | 0.386 | 0.067 | 0.008 | 6.1E-16 | 67.8 |
| BMI | Overall | rs6589936 | A | C | 0.387 | 0.054 | 0.009 | 2.2E-10 | 38.7 |
| BMI | Overall | rs7944782 | T | G | 0.502 | -0.069 | 0.008 | 3.6E-17 | 71.8 |
| BMI | Overall | rs2007518 | A | G | 0.562 | -0.062 | 0.008 | 2.0E-14 | 58.5 |
| BMI | Overall | rs1941213 | A | C | 0.711 | 0.052 | 0.009 | 1.6E-08 | 32.3 |
| BMI | Overall | rs329651 | T | G | 0.806 | 0.077 | 0.010 | 2.1E-14 | 58.0 |
| BMI | Overall | rs12364470 | T | G | 0.854 | -0.090 | 0.011 | 2.2E-17 | 72.3 |
| BMI | Overall | rs11611246 | T | G | 0.199 | 0.107 | 0.010 | 2.0E-28 | 124.3 |
| BMI | Overall | rs765125 | T | C | 0.586 | -0.047 | 0.008 | 1.3E-08 | 33.2 |
| BMI | Overall | rs10772983 | T | C | 0.539 | -0.048 | 0.008 | 6.2E-10 | 39.1 |
| BMI | Overall | rs11044430 | A | T | 0.844 | 0.077 | 0.011 | 3.8E-12 | 49.0 |
| BMI | Overall | rs11046972 | T | C | 0.071 | 0.085 | 0.015 | 3.7E-08 | 30.3 |
| BMI | Overall | rs10842240 | C | G | 0.126 | 0.094 | 0.012 | 1.2E-14 | 60.8 |
| BMI | Overall | rs11170468 | A | C | 0.783 | 0.062 | 0.009 | 1.1E-11 | 46.8 |
| BMI | Overall | rs1350430 | T | C | 0.533 | -0.060 | 0.008 | 2.1E-13 | 54.1 |
| BMI | Overall | rs7138803 | A | G | 0.387 | 0.143 | 0.008 | 3.1E-71 | 305.2 |
| BMI | Overall | rs2271189 | A | G | 0.396 | -0.068 | 0.009 | 9.3E-16 | 61.4 |
| BMI | Overall | rs7975187 | A | G | 0.771 | -0.066 | 0.010 | 3.9E-11 | 42.6 |
| BMI | Overall | rs650198 | T | C | 0.729 | -0.066 | 0.009 | 6.4E-13 | 52.0 |
| BMI | Overall | rs11115176 | T | C | 0.784 | 0.063 | 0.009 | 6.8E-12 | 47.5 |
| BMI | Overall | rs10506971 | A | G | 0.552 | -0.068 | 0.008 | 4.7E-17 | 69.8 |
| BMI | Overall | rs11105839 | A | T | 0.373 | -0.054 | 0.008 | 1.2E-11 | 44.2 |
| BMI | Overall | rs2712665 | T | C | 0.697 | -0.052 | 0.009 | 5.4E-09 | 32.3 |
| BMI | Overall | rs6539064 | C | G | 0.747 | 0.093 | 0.009 | 1.1E-23 | 104.3 |
| BMI | Overall | rs1860561 | A | G | 0.209 | 0.076 | 0.009 | 1.7E-16 | 70.0 |
| BMI | Overall | rs11066188 | A | G | 0.376 | -0.055 | 0.008 | 3.1E-12 | 50.8 |
| BMI | Overall | rs2707183 | T | G | 0.532 | -0.045 | 0.008 | 4.9E-08 | 29.9 |
| BMI | Overall | rs11615578 | T | C | 0.258 | 0.056 | 0.010 | 3.0E-09 | 34.2 |
| BMI | Overall | rs12369179 | T | C | 0.085 | -0.163 | 0.015 | 2.3E-28 | 120.3 |
| BMI | Overall | rs7306544 | T | C | 0.881 | -0.068 | 0.012 | 4.0E-08 | 29.8 |
| BMI | Overall | rs11614340 | T | C | 0.707 | -0.056 | 0.009 | 1.9E-10 | 42.3 |
| BMI | Overall | rs9512648 | A | G | 0.474 | 0.046 | 0.008 | 4.1E-08 | 31.2 |
| BMI | Overall | rs7323 | C | G | 0.268 | -0.080 | 0.009 | 2.2E-18 | 76.3 |
| BMI | Overall | rs4771218 | A | G | 0.624 | -0.068 | 0.009 | 3.2E-15 | 61.4 |
| BMI | Overall | rs9595908 | T | C | 0.643 | 0.074 | 0.008 | 3.7E-20 | 82.1 |
| BMI | Overall | rs9603697 | T | C | 0.318 | 0.064 | 0.009 | 1.7E-13 | 55.4 |
| BMI | Overall | rs12429545 | A | G | 0.122 | 0.150 | 0.012 | 1.4E-37 | 170.1 |
| BMI | Overall | rs962796 | T | C | 0.194 | 0.067 | 0.010 | 1.9E-11 | 44.4 |
| BMI | Overall | rs9569777 | T | G | 0.187 | -0.097 | 0.010 | 3.4E-21 | 91.6 |
| BMI | Overall | rs9527895 | T | C | 0.819 | -0.076 | 0.011 | 3.7E-12 | 47.2 |
| BMI | Overall | rs8181823 | A | C | 0.234 | -0.060 | 0.010 | 4.4E-10 | 39.1 |

| | | | | | | | | | |
|---|---|---|---|---|---|---|---|---|---|
| BMI | Overall | rs9599161 | T | C | 0.570 | 0.047 | 0.008 | 2.3E-09 | 37.5 |
| BMI | Overall | rs1441264 | A | G | 0.585 | 0.084 | 0.008 | 7.6E-25 | 104.8 |
| BMI | Overall | rs9602902 | T | C | 0.379 | 0.064 | 0.008 | 1.6E-14 | 62.1 |
| BMI | Overall | rs1927790 | T | C | 0.606 | -0.067 | 0.008 | 1.6E-17 | 76.6 |
| BMI | Overall | rs9168 | A | C | 0.275 | -0.067 | 0.009 | 1.1E-13 | 53.5 |
| BMI | Overall | rs9559022 | A | G | 0.816 | -0.058 | 0.011 | 4.9E-08 | 30.3 |
| BMI | Overall | rs1536053 | T | C | 0.310 | -0.059 | 0.009 | 8.0E-11 | 41.2 |
| BMI | Overall | rs12868881 | A | T | 0.407 | 0.066 | 0.008 | 1.4E-15 | 65.9 |
| BMI | Overall | rs10132280 | A | C | 0.311 | -0.103 | 0.009 | 2.3E-33 | 141.3 |
| BMI | Overall | rs4981693 | A | G | 0.774 | 0.097 | 0.010 | 7.9E-24 | 102.0 |
| BMI | Overall | rs225882 | T | C | 0.751 | 0.054 | 0.009 | 8.6E-10 | 39.4 |
| BMI | Overall | rs1958898 | C | G | 0.210 | -0.070 | 0.010 | 4.2E-12 | 48.3 |
| BMI | Overall | rs4900714 | T | G | 0.478 | 0.072 | 0.008 | 1.0E-18 | 77.9 |
| BMI | Overall | rs217671 | A | G | 0.726 | -0.072 | 0.009 | 4.1E-15 | 62.3 |
| BMI | Overall | rs4430672 | T | C | 0.196 | 0.060 | 0.010 | 5.0E-09 | 34.9 |
| BMI | Overall | rs3902951 | T | G | 0.769 | -0.064 | 0.009 | 2.9E-12 | 49.7 |
| BMI | Overall | rs17182027 | A | G | 0.565 | -0.053 | 0.008 | 1.5E-10 | 41.9 |
| BMI | Overall | rs7144011 | T | G | 0.233 | 0.126 | 0.010 | 2.4E-40 | 172.9 |
| BMI | Overall | rs4517716 | C | G | 0.792 | -0.056 | 0.010 | 4.3E-09 | 34.2 |
| BMI | Overall | rs12888545 | A | G | 0.746 | -0.064 | 0.010 | 1.8E-11 | 44.2 |
| BMI | Overall | rs1951455 | T | C | 0.281 | -0.071 | 0.009 | 6.0E-15 | 60.7 |
| BMI | Overall | rs9989141 | T | C | 0.618 | 0.081 | 0.008 | 8.2E-23 | 97.7 |
| BMI | Overall | rs8008285 | T | C | 0.849 | -0.065 | 0.012 | 1.7E-08 | 31.6 |
| BMI | Overall | rs7161194 | A | G | 0.344 | 0.091 | 0.009 | 2.2E-24 | 100.0 |
| BMI | Overall | rs4906263 | C | G | 0.656 | -0.085 | 0.009 | 8.1E-23 | 95.6 |
| BMI | Overall | rs2273175 | T | C | 0.679 | -0.058 | 0.009 | 3.7E-11 | 45.2 |
| BMI | Overall | rs12594043 | C | G | 0.521 | 0.049 | 0.008 | 1.3E-09 | 36.0 |
| BMI | Overall | rs7172627 | A | G | 0.523 | -0.055 | 0.008 | 1.8E-11 | 45.0 |
| BMI | Overall | rs11636611 | T | C | 0.498 | 0.050 | 0.008 | 8.9E-10 | 37.4 |
| BMI | Overall | rs9944219 | A | G | 0.604 | -0.057 | 0.008 | 8.1E-13 | 49.0 |
| BMI | Overall | rs12912198 | T | C | 0.273 | -0.048 | 0.009 | 3.4E-08 | 30.9 |
| BMI | Overall | rs6493498 | T | C | 0.447 | 0.066 | 0.008 | 4.8E-17 | 73.3 |
| BMI | Overall | rs10518694 | A | C | 0.139 | 0.069 | 0.012 | 3.4E-09 | 36.0 |
| BMI | Overall | rs12439632 | C | G | 0.168 | 0.070 | 0.011 | 3.7E-10 | 39.7 |
| BMI | Overall | rs339991 | A | G | 0.420 | -0.060 | 0.008 | 3.5E-13 | 54.1 |
| BMI | Overall | rs1559673 | A | C | 0.969 | 0.173 | 0.024 | 2.2E-13 | 53.7 |
| BMI | Overall | rs2241423 | A | G | 0.229 | -0.143 | 0.009 | 3.6E-54 | 246.0 |
| BMI | Overall | rs7171864 | A | G | 0.694 | 0.063 | 0.008 | 2.6E-14 | 60.3 |
| BMI | Overall | rs2470893 | T | C | 0.305 | 0.051 | 0.008 | 9.4E-10 | 39.6 |
| BMI | Overall | rs12914623 | C | G | 0.272 | -0.076 | 0.009 | 2.0E-16 | 70.0 |
| BMI | Overall | rs16946314 | A | G | 0.212 | -0.066 | 0.010 | 6.9E-11 | 42.6 |
| BMI | Overall | rs11633626 | A | C | 0.631 | -0.075 | 0.009 | 7.3E-19 | 76.1 |
| BMI | Overall | rs6496248 | A | T | 0.635 | 0.050 | 0.009 | 2.9E-09 | 33.4 |
| BMI | Overall | rs2715423 | A | G | 0.281 | -0.055 | 0.009 | 1.9E-09 | 36.6 |
| BMI | Overall | rs214249 | T | G | 0.607 | 0.066 | 0.008 | 2.7E-15 | 65.9 |
| BMI | Overall | rs12448257 | A | G | 0.219 | 0.077 | 0.010 | 9.0E-16 | 64.8 |
| BMI | Overall | rs879620 | T | C | 0.597 | 0.109 | 0.008 | 8.4E-39 | 176.7 |
| BMI | Overall | rs2058527 | T | G | 0.272 | -0.055 | 0.009 | 1.8E-09 | 36.6 |

| | | | | | | | | | |
|---|---|---|---|---|---|---|---|---|---|
| BMI | Overall | rs9926784 | T | C | 0.801 | 0.114 | 0.010 | 1.1E-30 | 127.4 |
| BMI | Overall | rs194809 | A | G | 0.187 | 0.061 | 0.011 | 4.9E-09 | 32.8 |
| BMI | Overall | rs2342892 | T | G | 0.486 | 0.061 | 0.008 | 1.3E-13 | 54.9 |
| BMI | Overall | rs7498665 | A | G | 0.618 | -0.137 | 0.008 | 1.1E-66 | 281.1 |
| BMI | Overall | rs3814883 | T | C | 0.480 | 0.109 | 0.008 | 1.5E-40 | 178.3 |
| BMI | Overall | rs2193101 | C | G | 0.195 | 0.068 | 0.010 | 3.3E-11 | 45.7 |
| BMI | Overall | rs1477199 | A | G | 0.843 | -0.106 | 0.011 | 5.1E-21 | 92.3 |
| BMI | Overall | rs9922708 | T | C | 0.440 | 0.321 | 0.008 | 0.0E+00 | 1743.1 |
| BMI | Overall | rs2962449 | T | C | 0.471 | 0.053 | 0.008 | 6.7E-11 | 42.6 |
| BMI | Overall | rs889398 | T | C | 0.415 | -0.094 | 0.008 | 3.2E-32 | 148.5 |
| BMI | Overall | rs17604662 | A | G | 0.890 | -0.086 | 0.013 | 1.0E-10 | 40.9 |
| BMI | Overall | rs756717 | A | G | 0.394 | -0.064 | 0.008 | 2.4E-15 | 62.1 |
| BMI | Overall | rs4390583 | A | C | 0.571 | -0.048 | 0.009 | 1.6E-08 | 30.9 |
| BMI | Overall | rs12922346 | C | G | 0.264 | 0.064 | 0.010 | 1.5E-11 | 44.2 |
| BMI | Overall | rs7206608 | C | G | 0.675 | -0.062 | 0.009 | 1.2E-12 | 52.2 |
| BMI | Overall | rs3923783 | A | C | 0.178 | -0.107 | 0.011 | 4.1E-23 | 101.8 |
| BMI | Overall | rs11078883 | C | G | 0.654 | -0.061 | 0.009 | 1.3E-12 | 49.0 |
| BMI | Overall | rs5862 | A | G | 0.542 | -0.049 | 0.008 | 2.3E-09 | 36.0 |
| BMI | Overall | rs1075901 | T | C | 0.452 | -0.057 | 0.008 | 4.4E-13 | 54.4 |
| BMI | Overall | rs4986044 | T | C | 0.455 | -0.085 | 0.008 | 1.3E-27 | 122.4 |
| BMI | Overall | rs8065172 | A | G | 0.238 | -0.060 | 0.010 | 5.3E-10 | 38.4 |
| BMI | Overall | rs12150665 | T | C | 0.593 | 0.081 | 0.008 | 1.7E-24 | 110.3 |
| BMI | Overall | rs6607337 | T | C | 0.298 | -0.060 | 0.009 | 2.5E-11 | 42.6 |
| BMI | Overall | rs7222349 | A | G | 0.356 | 0.048 | 0.009 | 1.7E-08 | 30.9 |
| BMI | Overall | rs208015 | T | C | 0.070 | 0.169 | 0.016 | 7.2E-26 | 113.8 |
| BMI | Overall | rs7207087 | C | G | 0.684 | 0.062 | 0.009 | 1.8E-12 | 52.2 |
| BMI | Overall | rs11649864 | A | G | 0.086 | 0.092 | 0.014 | 2.5E-10 | 41.0 |
| BMI | Overall | rs12602912 | T | C | 0.212 | 0.080 | 0.010 | 2.9E-16 | 68.9 |
| BMI | Overall | rs312750 | A | G | 0.499 | 0.047 | 0.008 | 2.6E-09 | 36.8 |
| BMI | Overall | rs12939549 | A | G | 0.557 | 0.087 | 0.008 | 3.7E-28 | 126.6 |
| BMI | Overall | rs4075482 | A | C | 0.368 | -0.063 | 0.009 | 2.0E-13 | 53.8 |
| BMI | Overall | rs8097672 | A | T | 0.855 | -0.100 | 0.012 | 3.3E-18 | 75.8 |
| BMI | Overall | rs477805 | A | G | 0.204 | -0.064 | 0.010 | 3.3E-10 | 40.7 |
| BMI | Overall | rs891387 | T | C | 0.495 | 0.100 | 0.008 | 9.3E-35 | 149.7 |
| BMI | Overall | rs1945160 | A | G | 0.382 | -0.050 | 0.009 | 5.7E-09 | 33.4 |
| BMI | Overall | rs1941696 | A | G | 0.517 | 0.054 | 0.008 | 4.7E-11 | 43.4 |
| BMI | Overall | rs474605 | A | G | 0.468 | -0.060 | 0.008 | 2.7E-13 | 54.1 |
| BMI | Overall | rs1356506 | T | C | 0.629 | 0.066 | 0.009 | 8.4E-15 | 57.9 |
| BMI | Overall | rs2612576 | A | T | 0.295 | -0.052 | 0.009 | 8.5E-09 | 32.3 |
| BMI | Overall | rs784257 | T | C | 0.188 | -0.071 | 0.011 | 3.1E-11 | 44.6 |
| BMI | Overall | rs9951619 | T | G | 0.224 | -0.073 | 0.009 | 2.3E-15 | 64.0 |
| BMI | Overall | rs6567160 | T | C | 0.752 | -0.265 | 0.009 | 7.8E-184 | 844.1 |
| BMI | Overall | rs17066856 | T | C | 0.892 | 0.168 | 0.013 | 7.1E-38 | 168.0 |
| BMI | Overall | rs543892 | A | G | 0.608 | 0.049 | 0.009 | 7.0E-09 | 31.5 |
| BMI | Overall | rs17783165 | T | C | 0.671 | -0.062 | 0.008 | 2.7E-13 | 56.7 |
| BMI | Overall | rs11150911 | A | C | 0.285 | 0.057 | 0.009 | 3.7E-11 | 43.0 |
| BMI | Overall | rs12981256 | A | G | 0.528 | 0.073 | 0.008 | 1.4E-18 | 78.9 |
| BMI | Overall | rs895330 | C | G | 0.806 | 0.094 | 0.011 | 1.6E-19 | 79.4 |

| | | | | | | | | | |
|---|---|---|---|---|---|---|---|---|---|
| BMI | Overall | rs273512 | T | C | 0.409 | 0.075 | 0.009 | 4.5E-19 | 75.1 |
| BMI | Overall | rs17724992 | A | G | 0.720 | 0.083 | 0.009 | 5.2E-21 | 91.3 |
| BMI | Overall | rs7258722 | A | T | 0.413 | -0.096 | 0.008 | 1.8E-30 | 138.4 |
| BMI | Overall | rs12611148 | A | C | 0.143 | -0.066 | 0.012 | 1.4E-08 | 32.6 |
| BMI | Overall | rs12462975 | A | G | 0.324 | 0.093 | 0.009 | 1.5E-25 | 115.0 |
| BMI | Overall | rs7245985 | T | G | 0.796 | 0.057 | 0.010 | 1.9E-08 | 32.1 |
| BMI | Overall | rs11882409 | A | C | 0.303 | 0.058 | 0.009 | 3.3E-10 | 40.6 |
| BMI | Overall | rs185350 | T | C | 0.490 | 0.066 | 0.008 | 9.1E-17 | 73.3 |
| BMI | Overall | rs769449 | A | G | 0.107 | -0.124 | 0.013 | 4.4E-21 | 91.3 |
| BMI | Overall | rs11672660 | T | C | 0.187 | -0.162 | 0.010 | 6.8E-60 | 259.1 |
| BMI | Overall | rs9304665 | A | T | 0.742 | 0.111 | 0.010 | 2.2E-31 | 132.3 |
| BMI | Overall | rs12608738 | A | G | 0.712 | 0.061 | 0.009 | 3.5E-11 | 44.7 |
| BMI | Overall | rs1884389 | T | C | 0.439 | -0.052 | 0.008 | 3.7E-10 | 40.4 |
| BMI | Overall | rs4813619 | T | G | 0.531 | -0.051 | 0.009 | 1.7E-09 | 34.7 |
| BMI | Overall | rs1884897 | A | G | 0.375 | -0.088 | 0.008 | 2.7E-28 | 117.1 |
| BMI | Overall | rs2423668 | T | C | 0.420 | 0.051 | 0.009 | 7.8E-09 | 34.7 |
| BMI | Overall | rs852056 | T | C | 0.254 | 0.059 | 0.009 | 2.1E-10 | 41.9 |
| BMI | Overall | rs1409818 | T | C | 0.107 | 0.094 | 0.013 | 2.6E-12 | 48.5 |
| BMI | Overall | rs8122855 | A | G | 0.338 | 0.066 | 0.009 | 4.1E-14 | 57.9 |
| BMI | Overall | rs6059578 | C | G | 0.668 | 0.050 | 0.009 | 6.0E-09 | 33.4 |
| BMI | Overall | rs16989232 | A | G | 0.386 | 0.055 | 0.008 | 6.2E-12 | 45.8 |
| BMI | Overall | rs11904898 | A | G | 0.234 | -0.065 | 0.010 | 3.2E-11 | 46.2 |
| BMI | Overall | rs2143624 | A | G | 0.361 | 0.045 | 0.008 | 2.1E-08 | 30.6 |
| BMI | Overall | rs17201143 | T | C | 0.305 | -0.054 | 0.009 | 1.1E-09 | 38.7 |
| BMI | Overall | rs17806224 | A | G | 0.181 | -0.125 | 0.011 | 7.9E-32 | 139.7 |
| BMI | Overall | rs1304549 | A | G | 0.226 | -0.057 | 0.010 | 1.4E-08 | 31.6 |
| BMI | Overall | rs6010784 | T | C | 0.506 | 0.051 | 0.008 | 6.9E-11 | 43.9 |
| BMI | Overall | rs6512302 | C | G | 0.739 | 0.064 | 0.010 | 1.5E-11 | 44.9 |
| BMI | Overall | rs2064044 | A | C | 0.806 | -0.059 | 0.010 | 1.0E-08 | 34.3 |
| BMI | Overall | rs2832283 | A | G | 0.225 | 0.055 | 0.010 | 4.7E-09 | 33.1 |
| BMI | Overall | rs13047416 | C | G | 0.626 | 0.073 | 0.009 | 6.9E-18 | 71.3 |
| BMI | Overall | rs13049280 | C | G | 0.643 | 0.050 | 0.009 | 7.0E-09 | 32.7 |
| BMI | Overall | rs2183588 | A | G | 0.345 | -0.055 | 0.009 | 1.9E-10 | 40.1 |
| BMI | Overall | rs427943 | A | C | 0.429 | -0.085 | 0.008 | 3.6E-25 | 108.4 |
| BMI | Overall | rs11538 | A | G | 0.828 | -0.066 | 0.011 | 1.1E-09 | 36.0 |
| BMI | Overall | rs2238799 | A | G | 0.619 | 0.049 | 0.009 | 8.6E-09 | 32.1 |
| BMI | Overall | rs12628891 | T | C | 0.319 | -0.055 | 0.009 | 5.9E-10 | 36.6 |
| BMI | Overall | rs12628051 | T | C | 0.641 | 0.077 | 0.009 | 2.9E-19 | 80.0 |
| BMI | Overall | rs738140 | A | G | 0.710 | 0.062 | 0.009 | 3.1E-12 | 51.4 |
| BMI | Women | rs12044597 | A | G | 0.496 | -0.066 | 0.011 | 2.3E-10 | 39.3 |
| BMI | Women | rs4648360 | T | C | 0.447 | -0.061 | 0.011 | 6.3E-09 | 33.3 |
| BMI | Women | rs309517 | A | T | 0.670 | -0.064 | 0.012 | 2.5E-08 | 30.7 |
| BMI | Women | rs12027258 | T | C | 0.835 | 0.080 | 0.014 | 4.7E-08 | 30.6 |
| BMI | Women | rs13374459 | T | C | 0.610 | -0.065 | 0.011 | 8.2E-09 | 34.5 |
| BMI | Women | rs2984618 | T | G | 0.443 | 0.080 | 0.011 | 2.2E-14 | 57.6 |
| BMI | Women | rs657452 | A | G | 0.405 | 0.098 | 0.011 | 1.1E-19 | 85.1 |
| BMI | Women | rs17424278 | A | C | 0.903 | -0.101 | 0.018 | 3.7E-08 | 30.5 |
| BMI | Women | rs1013293 | A | G | 0.411 | -0.074 | 0.011 | 2.2E-11 | 44.8 |

| | | | | | | | | | |
|---|---|---|---|---|---|---|---|---|---|
| BMI | Women | rs2568958 | A | G | 0.619 | 0.109 | 0.011 | 1.3E-24 | 106.5 |
| BMI | Women | rs6669189 | T | C | 0.405 | 0.101 | 0.011 | 4.7E-21 | 91.1 |
| BMI | Women | rs12729914 | T | C | 0.830 | -0.115 | 0.013 | 8.6E-18 | 73.5 |
| BMI | Women | rs2166171 | T | C | 0.623 | -0.066 | 0.012 | 7.2E-09 | 32.6 |
| BMI | Women | rs11165643 | T | C | 0.579 | 0.087 | 0.011 | 1.7E-16 | 67.7 |
| BMI | Women | rs4421623 | T | G | 0.210 | 0.088 | 0.013 | 1.1E-11 | 46.4 |
| BMI | Women | rs7550711 | T | C | 0.028 | 0.308 | 0.033 | 3.4E-21 | 88.9 |
| BMI | Women | rs197374 | T | C | 0.405 | 0.082 | 0.011 | 2.8E-13 | 55.3 |
| BMI | Women | rs3738476 | A | C | 0.877 | -0.094 | 0.017 | 2.2E-08 | 31.0 |
| BMI | Women | rs11264483 | C | G | 0.588 | 0.074 | 0.012 | 7.6E-11 | 41.2 |
| BMI | Women | rs12564992 | A | G | 0.888 | -0.100 | 0.017 | 2.4E-09 | 35.0 |
| BMI | Women | rs543874 | A | G | 0.777 | -0.277 | 0.013 | 8.7E-100 | 455.1 |
| BMI | Women | rs10920678 | A | G | 0.424 | 0.078 | 0.011 | 7.5E-14 | 54.9 |
| BMI | Women | rs10801377 | C | G | 0.420 | 0.071 | 0.011 | 1.9E-10 | 41.4 |
| BMI | Women | rs2820295 | A | G | 0.325 | 0.103 | 0.012 | 8.0E-19 | 79.5 |
| BMI | Women | rs17014375 | T | G | 0.870 | -0.091 | 0.016 | 1.5E-08 | 31.2 |
| BMI | Women | rs12042959 | A | G | 0.865 | 0.093 | 0.015 | 2.3E-09 | 36.8 |
| BMI | Women | rs6548237 | A | C | 0.160 | -0.277 | 0.014 | 4.4E-89 | 394.5 |
| BMI | Women | rs7571496 | A | G | 0.743 | 0.070 | 0.012 | 1.4E-08 | 31.1 |
| BMI | Women | rs7601870 | A | G | 0.453 | -0.063 | 0.011 | 1.2E-08 | 32.9 |
| BMI | Women | rs10182181 | A | G | 0.505 | -0.174 | 0.011 | 5.0E-63 | 270.8 |
| BMI | Women | rs10048652 | T | C | 0.450 | 0.065 | 0.011 | 3.5E-09 | 34.5 |
| BMI | Women | rs579119 | T | C | 0.670 | 0.069 | 0.011 | 6.7E-10 | 39.2 |
| BMI | Women | rs2346882 | A | G | 0.335 | -0.076 | 0.012 | 6.6E-11 | 43.3 |
| BMI | Women | rs7582359 | A | G | 0.331 | -0.080 | 0.012 | 1.0E-11 | 44.6 |
| BMI | Women | rs1861412 | A | G | 0.423 | -0.103 | 0.011 | 9.1E-21 | 87.4 |
| BMI | Women | rs6728037 | A | G | 0.386 | 0.100 | 0.011 | 6.5E-19 | 81.0 |
| BMI | Women | rs11678302 | T | G | 0.787 | -0.081 | 0.014 | 7.2E-09 | 33.6 |
| BMI | Women | rs12714199 | T | C | 0.615 | -0.070 | 0.011 | 5.1E-10 | 40.3 |
| BMI | Women | rs11677607 | T | C | 0.246 | -0.115 | 0.013 | 2.9E-19 | 79.0 |
| BMI | Women | rs1457457 | A | T | 0.532 | 0.068 | 0.011 | 3.8E-10 | 38.1 |
| BMI | Women | rs6739199 | T | C | 0.270 | 0.076 | 0.012 | 3.2E-10 | 40.4 |
| BMI | Women | rs1984559 | A | G | 0.808 | 0.078 | 0.014 | 3.3E-08 | 29.5 |
| BMI | Women | rs13002158 | A | G | 0.813 | -0.100 | 0.014 | 1.2E-12 | 51.4 |
| BMI | Women | rs12692596 | T | C | 0.361 | 0.070 | 0.011 | 9.7E-11 | 40.3 |
| BMI | Women | rs12692738 | T | C | 0.764 | -0.078 | 0.012 | 2.8E-10 | 38.8 |
| BMI | Women | rs7588437 | A | G | 0.367 | -0.085 | 0.011 | 8.4E-15 | 58.6 |
| BMI | Women | rs7594237 | A | T | 0.277 | -0.069 | 0.012 | 1.8E-08 | 32.7 |
| BMI | Women | rs4482463 | A | C | 0.917 | -0.175 | 0.020 | 5.2E-18 | 75.5 |
| BMI | Women | rs11692326 | T | C | 0.230 | 0.075 | 0.012 | 2.0E-09 | 36.0 |
| BMI | Women | rs4673553 | T | G | 0.555 | -0.074 | 0.011 | 2.6E-11 | 44.3 |
| BMI | Women | rs12612009 | T | G | 0.663 | -0.068 | 0.012 | 3.8E-09 | 34.5 |
| BMI | Women | rs6720868 | T | C | 0.306 | 0.081 | 0.012 | 4.6E-12 | 49.6 |
| BMI | Women | rs1801282 | C | G | 0.897 | -0.087 | 0.016 | 4.7E-08 | 30.4 |
| BMI | Women | rs2470520 | T | C | 0.577 | -0.061 | 0.011 | 4.2E-08 | 30.0 |
| BMI | Women | rs4858223 | T | C | 0.326 | -0.077 | 0.012 | 8.3E-11 | 41.5 |
| BMI | Women | rs13062093 | T | G | 0.632 | -0.066 | 0.012 | 8.0E-09 | 32.6 |
| BMI | Women | rs1799923 | A | G | 0.111 | -0.117 | 0.017 | 5.1E-12 | 48.2 |

| | | | | | | | | | |
|---|---|---|---|---|---|---|---|---|---|
| BMI | Women | rs2681781 | A | G | 0.480 | -0.110 | 0.011 | 1.6E-25 | 107.4 |
| BMI | Women | rs1916801 | A | T | 0.620 | 0.082 | 0.011 | 1.8E-14 | 59.7 |
| BMI | Women | rs1452075 | T | C | 0.732 | 0.076 | 0.012 | 1.5E-10 | 40.4 |
| BMI | Women | rs925018 | C | G | 0.665 | -0.068 | 0.011 | 9.8E-10 | 38.1 |
| BMI | Women | rs7647242 | T | C | 0.394 | 0.070 | 0.011 | 3.9E-10 | 39.7 |
| BMI | Women | rs13068138 | T | C | 0.192 | 0.111 | 0.013 | 3.6E-17 | 73.2 |
| BMI | Women | rs1949197 | A | G | 0.401 | 0.067 | 0.011 | 3.0E-09 | 36.5 |
| BMI | Women | rs1492014 | T | C | 0.568 | -0.070 | 0.011 | 2.8E-10 | 39.7 |
| BMI | Women | rs1436348 | A | G | 0.432 | -0.064 | 0.011 | 6.3E-09 | 33.9 |
| BMI | Women | rs1289736 | T | C | 0.543 | 0.067 | 0.011 | 1.4E-09 | 36.5 |
| BMI | Women | rs17619973 | A | G | 0.936 | 0.116 | 0.021 | 1.3E-08 | 31.7 |
| BMI | Women | rs7428670 | C | G | 0.269 | 0.097 | 0.012 | 3.1E-15 | 60.4 |
| BMI | Women | rs7621025 | T | C | 0.261 | -0.096 | 0.012 | 1.6E-15 | 64.0 |
| BMI | Women | rs16851483 | T | G | 0.073 | 0.174 | 0.022 | 2.3E-15 | 62.3 |
| BMI | Women | rs355754 | T | C | 0.604 | -0.068 | 0.011 | 4.3E-10 | 38.1 |
| BMI | Women | rs247975 | T | C | 0.466 | -0.087 | 0.011 | 3.6E-15 | 62.6 |
| BMI | Women | rs10513801 | T | G | 0.878 | 0.169 | 0.015 | 1.3E-27 | 121.0 |
| BMI | Women | rs4677812 | A | C | 0.293 | -0.069 | 0.012 | 9.4E-09 | 33.2 |
| BMI | Women | rs1477887 | A | G | 0.441 | -0.075 | 0.011 | 1.7E-11 | 46.0 |
| BMI | Women | rs9291467 | T | C | 0.465 | 0.072 | 0.011 | 5.7E-11 | 42.5 |
| BMI | Women | rs6448587 | A | C | 0.802 | 0.081 | 0.014 | 1.6E-08 | 31.4 |
| BMI | Women | rs337637 | A | G | 0.358 | -0.060 | 0.011 | 3.0E-08 | 29.5 |
| BMI | Women | rs10938397 | A | G | 0.571 | -0.151 | 0.011 | 1.7E-45 | 205.0 |
| BMI | Women | rs2271046 | A | T | 0.694 | -0.066 | 0.012 | 2.4E-08 | 30.0 |
| BMI | Women | rs2192158 | A | G | 0.449 | 0.070 | 0.011 | 2.1E-10 | 40.3 |
| BMI | Women | rs11945861 | A | G | 0.247 | -0.072 | 0.013 | 3.1E-08 | 30.9 |
| BMI | Women | rs10002111 | A | G | 0.224 | 0.081 | 0.013 | 9.3E-10 | 36.4 |
| BMI | Women | rs4148155 | A | G | 0.887 | 0.098 | 0.017 | 6.2E-09 | 33.6 |
| BMI | Women | rs12645001 | A | G | 0.356 | -0.068 | 0.012 | 4.1E-09 | 34.5 |
| BMI | Women | rs13107325 | T | C | 0.083 | 0.217 | 0.021 | 9.3E-25 | 105.5 |
| BMI | Women | rs4834272 | T | C | 0.687 | -0.069 | 0.011 | 8.2E-10 | 38.7 |
| BMI | Women | rs1403846 | T | C | 0.833 | 0.078 | 0.014 | 1.6E-08 | 31.2 |
| BMI | Women | rs4864201 | T | C | 0.346 | 0.063 | 0.011 | 6.3E-09 | 32.4 |
| BMI | Women | rs10000502 | T | G | 0.367 | -0.073 | 0.012 | 1.8E-10 | 39.6 |
| BMI | Women | rs750090 | T | C | 0.624 | 0.075 | 0.012 | 7.4E-11 | 42.3 |
| BMI | Women | rs13110266 | A | G | 0.403 | -0.068 | 0.011 | 1.5E-10 | 41.7 |
| BMI | Women | rs1491333 | T | G | 0.263 | -0.071 | 0.012 | 9.9E-09 | 32.0 |
| BMI | Women | rs6890310 | A | G | 0.294 | -0.074 | 0.012 | 1.4E-09 | 37.9 |
| BMI | Women | rs782971 | A | G | 0.754 | -0.083 | 0.012 | 4.9E-12 | 47.3 |
| BMI | Women | rs10471636 | A | G | 0.525 | -0.082 | 0.012 | 4.7E-13 | 50.2 |
| BMI | Women | rs2112347 | T | G | 0.630 | 0.149 | 0.011 | 2.1E-42 | 180.5 |
| BMI | Women | rs6870983 | T | C | 0.206 | -0.102 | 0.012 | 9.6E-16 | 66.5 |
| BMI | Women | rs2161228 | T | C | 0.101 | 0.135 | 0.018 | 5.5E-14 | 57.7 |
| BMI | Women | rs10473922 | A | G | 0.458 | 0.064 | 0.011 | 7.0E-09 | 33.4 |
| BMI | Women | rs6885199 | A | G | 0.036 | 0.187 | 0.031 | 8.5E-10 | 37.1 |
| BMI | Women | rs2161097 | T | C | 0.442 | 0.062 | 0.011 | 2.3E-08 | 31.5 |
| BMI | Women | rs158186 | A | G | 0.166 | -0.138 | 0.015 | 3.2E-20 | 85.7 |
| BMI | Women | rs4895231 | C | G | 0.468 | 0.069 | 0.011 | 9.3E-10 | 38.7 |

| | | | | | | | | | |
|---|---|---|---|---|---|---|---|---|---|
| BMI | Women | rs13174863 | A | G | 0.854 | -0.130 | 0.015 | 4.5E-18 | 75.9 |
| BMI | Women | rs4569924 | T | C | 0.435 | 0.076 | 0.011 | 7.4E-13 | 51.6 |
| BMI | Women | rs6556810 | T | C | 0.534 | -0.060 | 0.011 | 5.0E-08 | 29.1 |
| BMI | Women | rs2053682 | A | C | 0.678 | 0.088 | 0.012 | 1.9E-13 | 53.6 |
| BMI | Women | rs2228213 | A | G | 0.337 | -0.071 | 0.011 | 9.2E-11 | 41.4 |
| BMI | Women | rs11757278 | T | C | 0.695 | 0.065 | 0.012 | 4.8E-08 | 29.6 |
| BMI | Women | rs3806114 | A | G | 0.694 | -0.068 | 0.012 | 4.5E-09 | 34.5 |
| BMI | Women | rs2178899 | A | T | 0.885 | 0.119 | 0.016 | 7.2E-14 | 56.5 |
| BMI | Women | rs11751591 | A | G | 0.153 | -0.089 | 0.015 | 1.2E-08 | 33.4 |
| BMI | Women | rs2814992 | A | G | 0.667 | -0.138 | 0.011 | 1.2E-35 | 155.7 |
| BMI | Women | rs1579557 | T | C | 0.288 | 0.102 | 0.012 | 3.1E-17 | 71.9 |
| BMI | Women | rs943005 | T | C | 0.147 | 0.191 | 0.014 | 2.0E-43 | 188.4 |
| BMI | Women | rs2465043 | A | G | 0.351 | -0.068 | 0.012 | 3.2E-09 | 35.0 |
| BMI | Women | rs209416 | A | T | 0.324 | 0.080 | 0.012 | 8.7E-12 | 47.8 |
| BMI | Women | rs13209872 | C | G | 0.345 | -0.068 | 0.012 | 6.1E-09 | 35.0 |
| BMI | Women | rs12202969 | A | G | 0.479 | -0.070 | 0.011 | 1.9E-10 | 39.7 |
| BMI | Women | rs1417665 | T | C | 0.802 | 0.096 | 0.014 | 5.7E-12 | 47.1 |
| BMI | Women | rs768023 | A | G | 0.603 | 0.089 | 0.011 | 1.4E-16 | 70.7 |
| BMI | Women | rs2357760 | A | G | 0.673 | 0.076 | 0.011 | 1.1E-11 | 47.8 |
| BMI | Women | rs11754747 | T | C | 0.271 | 0.070 | 0.012 | 1.3E-08 | 31.5 |
| BMI | Women | rs198665 | A | C | 0.394 | 0.064 | 0.011 | 4.4E-09 | 33.9 |
| BMI | Women | rs2185027 | A | C | 0.698 | -0.074 | 0.012 | 1.1E-10 | 41.2 |
| BMI | Women | rs9478496 | T | C | 0.847 | -0.082 | 0.015 | 4.6E-08 | 30.1 |
| BMI | Women | rs13213408 | A | T | 0.847 | 0.095 | 0.016 | 4.3E-09 | 35.6 |
| BMI | Women | rs11765458 | T | C | 0.781 | 0.076 | 0.014 | 4.1E-08 | 29.7 |
| BMI | Women | rs836525 | T | C | 0.173 | 0.085 | 0.015 | 9.8E-09 | 32.2 |
| BMI | Women | rs4307239 | A | G | 0.536 | -0.068 | 0.011 | 1.3E-09 | 37.6 |
| BMI | Women | rs11971098 | A | G | 0.909 | -0.114 | 0.020 | 6.5E-09 | 33.4 |
| BMI | Women | rs10499694 | A | G | 0.494 | 0.065 | 0.011 | 4.0E-10 | 38.2 |
| BMI | Women | rs10237317 | A | G | 0.588 | -0.066 | 0.011 | 6.8E-10 | 39.3 |
| BMI | Women | rs2267812 | A | C | 0.807 | -0.083 | 0.014 | 1.7E-09 | 35.2 |
| BMI | Women | rs1167800 | A | G | 0.553 | 0.096 | 0.011 | 3.7E-19 | 82.6 |
| BMI | Women | rs7805441 | T | C | 0.547 | 0.070 | 0.011 | 2.0E-10 | 40.3 |
| BMI | Women | rs2528531 | A | C | 0.651 | -0.070 | 0.012 | 1.3E-09 | 36.5 |
| BMI | Women | rs13240600 | A | G | 0.835 | 0.083 | 0.014 | 9.7E-09 | 33.3 |
| BMI | Women | rs2299383 | T | C | 0.395 | 0.087 | 0.011 | 2.7E-16 | 68.4 |
| BMI | Women | rs7788008 | A | G | 0.430 | -0.082 | 0.011 | 1.1E-13 | 55.3 |
| BMI | Women | rs2045293 | T | C | 0.417 | 0.091 | 0.011 | 4.1E-16 | 67.5 |
| BMI | Women | rs2289705 | A | G | 0.796 | -0.072 | 0.012 | 1.5E-08 | 33.3 |
| BMI | Women | rs2968864 | T | C | 0.756 | 0.078 | 0.013 | 9.3E-10 | 36.4 |
| BMI | Women | rs4240673 | T | C | 0.456 | 0.085 | 0.011 | 1.0E-15 | 64.0 |
| BMI | Women | rs2959592 | T | C | 0.809 | 0.082 | 0.013 | 6.3E-10 | 36.9 |
| BMI | Women | rs1362910 | A | G | 0.413 | 0.064 | 0.011 | 3.0E-09 | 33.9 |
| BMI | Women | rs4320543 | A | G | 0.419 | -0.062 | 0.011 | 2.9E-08 | 31.5 |
| BMI | Women | rs12681792 | A | C | 0.204 | 0.088 | 0.014 | 2.6E-10 | 39.8 |
| BMI | Women | rs1431659 | A | G | 0.274 | 0.094 | 0.012 | 2.9E-14 | 56.8 |
| BMI | Women | rs17405819 | T | C | 0.683 | 0.105 | 0.012 | 7.8E-20 | 82.5 |
| BMI | Women | rs12680842 | A | G | 0.684 | 0.062 | 0.012 | 3.4E-08 | 29.3 |

| | | | | | | | | | |
|---|---|---|---|---|---|---|---|---|---|
| BMI | Women | rs2721965 | A | C | 0.669 | 0.089 | 0.012 | 2.1E-14 | 60.1 |
| BMI | Women | rs10099330 | A | G | 0.538 | -0.063 | 0.011 | 9.9E-09 | 32.4 |
| BMI | Women | rs10124645 | A | G | 0.592 | 0.065 | 0.011 | 5.0E-09 | 35.0 |
| BMI | Women | rs10756714 | A | G | 0.555 | 0.100 | 0.011 | 1.5E-19 | 81.8 |
| BMI | Women | rs10962550 | C | G | 0.174 | 0.092 | 0.014 | 1.7E-10 | 41.0 |
| BMI | Women | rs16912921 | A | C | 0.313 | 0.120 | 0.011 | 1.6E-26 | 117.2 |
| BMI | Women | rs973345 | T | C | 0.611 | 0.068 | 0.011 | 1.1E-09 | 38.1 |
| BMI | Women | rs563132 | A | T | 0.605 | 0.087 | 0.012 | 2.0E-14 | 56.3 |
| BMI | Women | rs10797115 | T | C | 0.532 | 0.077 | 0.011 | 4.1E-12 | 48.4 |
| BMI | Women | rs10992867 | A | G | 0.269 | 0.089 | 0.012 | 8.9E-13 | 50.6 |
| BMI | Women | rs10118701 | A | G | 0.672 | -0.085 | 0.012 | 2.2E-13 | 53.8 |
| BMI | Women | rs7024334 | T | G | 0.226 | 0.076 | 0.012 | 1.6E-09 | 37.4 |
| BMI | Women | rs1928295 | T | C | 0.571 | 0.077 | 0.011 | 2.8E-13 | 52.9 |
| BMI | Women | rs3829849 | T | C | 0.350 | 0.065 | 0.011 | 1.9E-09 | 35.0 |
| BMI | Women | rs10733682 | A | G | 0.458 | 0.076 | 0.011 | 1.9E-12 | 51.6 |
| BMI | Women | rs2267958 | A | G | 0.510 | -0.077 | 0.012 | 1.9E-11 | 45.0 |
| BMI | Women | rs1009473 | T | C | 0.526 | -0.061 | 0.011 | 3.4E-08 | 30.5 |
| BMI | Women | rs7084454 | A | G | 0.306 | 0.108 | 0.012 | 7.4E-20 | 81.0 |
| BMI | Women | rs12762034 | T | C | 0.928 | -0.123 | 0.021 | 2.3E-09 | 35.2 |
| BMI | Women | rs1624134 | C | G | 0.392 | 0.062 | 0.011 | 3.5E-08 | 31.0 |
| BMI | Women | rs10823897 | A | T | 0.416 | 0.071 | 0.011 | 2.1E-10 | 40.8 |
| BMI | Women | rs10761785 | T | G | 0.514 | -0.083 | 0.011 | 4.2E-15 | 61.1 |
| BMI | Women | rs10824347 | A | T | 0.771 | 0.085 | 0.013 | 5.5E-11 | 42.5 |
| BMI | Women | rs7899106 | A | G | 0.950 | -0.180 | 0.024 | 7.6E-14 | 56.3 |
| BMI | Women | rs563296 | A | G | 0.558 | 0.084 | 0.012 | 8.8E-14 | 53.2 |
| BMI | Women | rs3977755 | T | C | 0.287 | -0.069 | 0.012 | 8.5E-09 | 32.7 |
| BMI | Women | rs6585198 | A | G | 0.444 | -0.074 | 0.011 | 1.2E-12 | 49.0 |
| BMI | Women | rs1681743 | T | C | 0.401 | -0.063 | 0.012 | 3.7E-08 | 29.8 |
| BMI | Women | rs1561589 | A | G | 0.361 | 0.076 | 0.011 | 4.2E-12 | 47.2 |
| BMI | Women | rs4880341 | T | C | 0.572 | -0.069 | 0.011 | 4.5E-10 | 38.7 |
| BMI | Women | rs7395632 | T | C | 0.357 | -0.062 | 0.012 | 3.9E-08 | 29.3 |
| BMI | Women | rs4929923 | T | C | 0.337 | -0.106 | 0.011 | 2.5E-22 | 92.3 |
| BMI | Women | rs11022762 | T | C | 0.368 | -0.074 | 0.012 | 5.6E-11 | 41.2 |
| BMI | Women | rs6265 | T | C | 0.183 | -0.177 | 0.013 | 8.0E-40 | 172.7 |
| BMI | Women | rs2862961 | A | G | 0.699 | -0.115 | 0.012 | 5.3E-22 | 91.4 |
| BMI | Women | rs10742752 | T | C | 0.375 | -0.063 | 0.011 | 4.0E-09 | 36.0 |
| BMI | Women | rs7124681 | A | C | 0.418 | 0.119 | 0.011 | 9.3E-29 | 126.1 |
| BMI | Women | rs893006 | A | C | 0.721 | 0.083 | 0.012 | 4.0E-13 | 52.0 |
| BMI | Women | rs7102454 | T | C | 0.643 | -0.080 | 0.012 | 5.1E-12 | 47.8 |
| BMI | Women | rs2605603 | A | G | 0.481 | -0.063 | 0.011 | 1.4E-09 | 36.0 |
| BMI | Women | rs7105462 | A | G | 0.581 | 0.069 | 0.011 | 4.4E-10 | 39.2 |
| BMI | Women | rs1048932 | A | C | 0.440 | -0.087 | 0.011 | 2.5E-16 | 68.4 |
| BMI | Women | rs1003081 | T | C | 0.435 | 0.069 | 0.011 | 6.3E-11 | 42.8 |
| BMI | Women | rs4459316 | T | C | 0.551 | -0.074 | 0.011 | 2.3E-11 | 44.8 |
| BMI | Women | rs4936175 | T | C | 0.559 | -0.076 | 0.011 | 6.7E-12 | 47.2 |
| BMI | Women | rs329651 | T | G | 0.805 | 0.087 | 0.013 | 2.0E-10 | 41.8 |
| BMI | Women | rs12364470 | T | G | 0.855 | -0.096 | 0.014 | 2.5E-11 | 44.4 |
| BMI | Women | rs11611246 | T | G | 0.199 | 0.102 | 0.013 | 1.3E-14 | 61.7 |

| | | | | | | | | | |
|---|---|---|---|---|---|---|---|---|---|
| BMI | Women | rs7976757 | T | C | 0.177 | -0.092 | 0.014 | 2.9E-10 | 41.0 |
| BMI | Women | rs16926318 | A | G | 0.921 | -0.109 | 0.020 | 4.2E-08 | 30.7 |
| BMI | Women | rs16926778 | C | G | 0.077 | 0.125 | 0.021 | 1.4E-09 | 36.6 |
| BMI | Women | rs7138803 | A | G | 0.388 | 0.140 | 0.011 | 9.7E-38 | 160.1 |
| BMI | Women | rs2271189 | A | G | 0.396 | -0.070 | 0.012 | 6.4E-10 | 37.0 |
| BMI | Women | rs11173522 | A | C | 0.233 | 0.081 | 0.013 | 9.0E-10 | 36.4 |
| BMI | Women | rs645026 | A | G | 0.744 | -0.077 | 0.012 | 4.8E-10 | 37.9 |
| BMI | Women | rs2279574 | A | C | 0.549 | -0.079 | 0.011 | 2.0E-12 | 51.5 |
| BMI | Women | rs2712643 | A | C | 0.239 | -0.074 | 0.012 | 1.5E-09 | 37.9 |
| BMI | Women | rs17033633 | T | C | 0.169 | -0.100 | 0.014 | 7.3E-12 | 48.5 |
| BMI | Women | rs6606686 | C | G | 0.698 | -0.086 | 0.011 | 2.5E-14 | 59.9 |
| BMI | Women | rs1466852 | A | G | 0.227 | 0.078 | 0.013 | 3.6E-09 | 33.9 |
| BMI | Women | rs6489156 | T | C | 0.734 | 0.107 | 0.012 | 1.0E-17 | 72.9 |
| BMI | Women | rs7133378 | A | G | 0.334 | 0.076 | 0.012 | 1.9E-11 | 43.3 |
| BMI | Women | rs1967772 | A | G | 0.271 | -0.079 | 0.012 | 1.1E-10 | 40.3 |
| BMI | Women | rs1045411 | T | C | 0.283 | -0.067 | 0.012 | 2.6E-08 | 30.9 |
| BMI | Women | rs4942925 | T | C | 0.574 | 0.062 | 0.011 | 2.8E-09 | 34.9 |
| BMI | Women | rs12429545 | A | G | 0.120 | 0.140 | 0.016 | 1.1E-18 | 77.8 |
| BMI | Women | rs6561937 | A | T | 0.767 | -0.095 | 0.013 | 6.0E-13 | 53.2 |
| BMI | Women | rs9538162 | T | C | 0.579 | 0.069 | 0.011 | 6.3E-10 | 38.7 |
| BMI | Women | rs9571687 | A | C | 0.342 | -0.070 | 0.012 | 3.2E-09 | 36.5 |
| BMI | Women | rs1441264 | A | G | 0.583 | 0.082 | 0.011 | 5.2E-14 | 55.3 |
| BMI | Women | rs9531786 | C | G | 0.371 | -0.066 | 0.012 | 8.4E-09 | 33.1 |
| BMI | Women | rs4343164 | T | C | 0.276 | 0.072 | 0.012 | 4.0E-09 | 33.3 |
| BMI | Women | rs1927790 | T | C | 0.608 | -0.065 | 0.011 | 1.3E-09 | 37.7 |
| BMI | Women | rs4586287 | C | G | 0.674 | 0.066 | 0.012 | 2.9E-08 | 30.5 |
| BMI | Women | rs7335249 | T | C | 0.558 | 0.065 | 0.011 | 4.9E-09 | 34.5 |
| BMI | Women | rs2479958 | A | G | 0.486 | 0.065 | 0.012 | 1.4E-08 | 31.6 |
| BMI | Women | rs2754079 | A | G | 0.571 | -0.075 | 0.011 | 2.3E-11 | 46.0 |
| BMI | Women | rs1569979 | A | G | 0.774 | 0.111 | 0.013 | 1.6E-17 | 72.6 |
| BMI | Women | rs17522122 | T | G | 0.467 | 0.080 | 0.011 | 6.5E-14 | 57.6 |
| BMI | Women | rs2806034 | T | G | 0.668 | 0.066 | 0.011 | 5.3E-09 | 35.5 |
| BMI | Women | rs7141420 | T | C | 0.549 | 0.101 | 0.011 | 7.8E-22 | 91.1 |
| BMI | Women | rs1285997 | C | G | 0.288 | -0.072 | 0.012 | 3.2E-09 | 36.0 |
| BMI | Women | rs9989141 | T | C | 0.616 | 0.081 | 0.011 | 3.2E-13 | 53.4 |
| BMI | Women | rs3803286 | A | G | 0.341 | 0.083 | 0.012 | 8.3E-13 | 51.4 |
| BMI | Women | rs709400 | A | G | 0.621 | 0.088 | 0.011 | 4.4E-16 | 64.0 |
| BMI | Women | rs12101625 | C | G | 0.614 | 0.066 | 0.012 | 5.5E-09 | 33.1 |
| BMI | Women | rs8038522 | T | G | 0.413 | -0.067 | 0.011 | 1.3E-09 | 37.1 |
| BMI | Women | rs6493498 | T | C | 0.446 | 0.078 | 0.011 | 1.5E-13 | 54.9 |
| BMI | Women | rs8024806 | T | C | 0.937 | 0.131 | 0.023 | 9.8E-09 | 33.5 |
| BMI | Women | rs8033510 | T | C | 0.364 | 0.066 | 0.012 | 1.1E-08 | 32.6 |
| BMI | Women | rs4776982 | A | G | 0.769 | 0.149 | 0.012 | 4.2E-33 | 143.1 |
| BMI | Women | rs12914489 | A | G | 0.081 | 0.109 | 0.017 | 1.6E-10 | 39.8 |
| BMI | Women | rs12593036 | A | G | 0.690 | 0.074 | 0.012 | 6.6E-10 | 37.9 |
| BMI | Women | rs11073383 | A | G | 0.480 | -0.076 | 0.011 | 3.3E-12 | 47.8 |
| BMI | Women | rs214249 | T | G | 0.607 | 0.080 | 0.012 | 1.4E-12 | 48.4 |
| BMI | Women | rs12448257 | A | G | 0.219 | 0.072 | 0.013 | 3.1E-08 | 30.9 |

| | | | | | | | | | |
|---|---|---|---|---|---|---|---|---|---|
| BMI | Women | rs879620 | T | C | 0.596 | 0.119 | 0.011 | 5.4E-26 | 115.3 |
| BMI | Women | rs9926784 | T | C | 0.800 | 0.121 | 0.013 | 6.0E-20 | 81.0 |
| BMI | Women | rs17767765 | T | C | 0.515 | -0.061 | 0.011 | 2.6E-08 | 30.5 |
| BMI | Women | rs7498665 | A | G | 0.619 | -0.127 | 0.011 | 1.9E-32 | 144.0 |
| BMI | Women | rs2289292 | T | C | 0.332 | -0.091 | 0.012 | 1.1E-14 | 57.8 |
| BMI | Women | rs4889606 | A | G | 0.631 | 0.129 | 0.011 | 1.3E-32 | 135.8 |
| BMI | Women | rs1558902 | A | T | 0.409 | 0.337 | 0.011 | 1.1E-215 | 1018.2 |
| BMI | Women | rs4783718 | T | C | 0.419 | 0.098 | 0.011 | 1.8E-19 | 78.7 |
| BMI | Women | rs811054 | T | C | 0.533 | 0.078 | 0.011 | 3.2E-13 | 54.2 |
| BMI | Women | rs2012502 | A | C | 0.384 | 0.074 | 0.011 | 3.0E-11 | 44.3 |
| BMI | Women | rs4790292 | A | C | 0.147 | -0.126 | 0.016 | 1.7E-15 | 63.5 |
| BMI | Women | rs3826408 | T | C | 0.418 | 0.058 | 0.011 | 4.5E-08 | 30.3 |
| BMI | Women | rs1075901 | T | C | 0.452 | -0.069 | 0.011 | 4.4E-11 | 42.8 |
| BMI | Women | rs4986044 | T | C | 0.453 | -0.083 | 0.011 | 2.2E-15 | 61.8 |
| BMI | Women | rs12150665 | T | C | 0.592 | 0.080 | 0.011 | 5.9E-14 | 57.6 |
| BMI | Women | rs9906944 | T | C | 0.355 | -0.091 | 0.012 | 7.3E-15 | 62.7 |
| BMI | Women | rs7211966 | T | C | 0.029 | -0.226 | 0.038 | 4.9E-09 | 34.5 |
| BMI | Women | rs8075273 | A | C | 0.290 | -0.077 | 0.012 | 5.6E-11 | 44.4 |
| BMI | Women | rs2619976 | T | C | 0.399 | 0.070 | 0.012 | 6.2E-10 | 37.0 |
| BMI | Women | rs12949279 | T | C | 0.563 | 0.086 | 0.011 | 6.1E-16 | 66.2 |
| BMI | Women | rs4969387 | C | G | 0.738 | -0.074 | 0.013 | 6.9E-09 | 32.5 |
| BMI | Women | rs9955276 | T | C | 0.144 | 0.107 | 0.015 | 8.0E-12 | 48.1 |
| BMI | Women | rs1808579 | T | C | 0.481 | -0.095 | 0.011 | 9.4E-20 | 81.0 |
| BMI | Women | rs16975921 | A | T | 0.695 | -0.078 | 0.012 | 3.9E-11 | 42.5 |
| BMI | Women | rs1518159 | T | C | 0.250 | -0.091 | 0.012 | 6.7E-13 | 52.8 |
| BMI | Women | rs7239114 | A | G | 0.542 | 0.075 | 0.011 | 3.9E-12 | 49.6 |
| BMI | Women | rs6567160 | T | C | 0.751 | -0.273 | 0.012 | 8.7E-108 | 475.6 |
| BMI | Women | rs17066856 | T | C | 0.891 | 0.191 | 0.018 | 1.4E-27 | 115.7 |
| BMI | Women | rs12981256 | A | G | 0.527 | 0.087 | 0.011 | 3.9E-15 | 62.6 |
| BMI | Women | rs895330 | C | G | 0.807 | 0.087 | 0.014 | 6.6E-10 | 39.0 |
| BMI | Women | rs273504 | A | G | 0.576 | -0.073 | 0.011 | 1.3E-10 | 43.1 |
| BMI | Women | rs17724992 | A | G | 0.719 | 0.082 | 0.012 | 6.7E-12 | 46.2 |
| BMI | Women | rs11880870 | A | G | 0.519 | 0.095 | 0.011 | 3.3E-18 | 73.4 |
| BMI | Women | rs17751061 | T | C | 0.159 | -0.087 | 0.015 | 1.3E-08 | 32.0 |
| BMI | Women | rs12462975 | A | G | 0.325 | 0.098 | 0.012 | 2.6E-16 | 65.9 |
| BMI | Women | rs11667280 | C | G | 0.797 | 0.076 | 0.014 | 3.1E-08 | 29.7 |
| BMI | Women | rs7250833 | T | C | 0.311 | 0.072 | 0.012 | 7.2E-10 | 38.5 |
| BMI | Women | rs185350 | T | C | 0.490 | 0.062 | 0.011 | 7.1E-09 | 33.9 |
| BMI | Women | rs11672660 | T | C | 0.187 | -0.154 | 0.013 | 5.9E-31 | 131.4 |
| BMI | Women | rs9304665 | A | T | 0.740 | 0.098 | 0.013 | 1.8E-14 | 57.1 |
| BMI | Women | rs1884389 | T | C | 0.440 | -0.067 | 0.011 | 1.2E-09 | 37.1 |
| BMI | Women | rs1884897 | A | G | 0.375 | -0.081 | 0.011 | 9.1E-14 | 54.0 |
| BMI | Women | rs1409818 | T | C | 0.108 | 0.121 | 0.018 | 1.6E-11 | 46.4 |
| BMI | Women | rs6050446 | A | G | 0.032 | -0.179 | 0.031 | 6.2E-09 | 33.8 |
| BMI | Women | rs6142096 | A | G | 0.520 | 0.064 | 0.011 | 4.2E-09 | 33.9 |
| BMI | Women | rs16989232 | A | G | 0.385 | 0.062 | 0.011 | 1.5E-08 | 31.0 |
| BMI | Women | rs17806224 | A | G | 0.181 | -0.139 | 0.014 | 3.5E-22 | 92.8 |
| BMI | Women | rs6010784 | T | C | 0.508 | 0.061 | 0.011 | 1.1E-08 | 32.8 |

| | | | | | | | | | |
|---|---|---|---|---|---|---|---|---|---|
| BMI | Women | rs2836754 | T | C | 0.369 | -0.072 | 0.011 | 3.7E-11 | 45.9 |
| BMI | Women | rs8126575 | T | G | 0.868 | 0.088 | 0.016 | 3.0E-08 | 31.1 |
| BMI | Women | rs427943 | A | C | 0.428 | -0.085 | 0.011 | 1.4E-14 | 59.2 |
| BMI | Women | rs4820408 | T | G | 0.375 | 0.065 | 0.011 | 1.4E-09 | 38.2 |
| BMI | Women | rs13055841 | A | G | 0.693 | 0.066 | 0.012 | 3.7E-08 | 30.5 |
| BMI | Women | rs13053342 | A | G | 0.400 | 0.074 | 0.012 | 1.3E-10 | 40.6 |
| BMI | Men | rs2803316 | A | G | 0.457 | -0.072 | 0.012 | 4.0E-09 | 35.5 |
| BMI | Men | rs3762444 | T | C | 0.461 | -0.093 | 0.012 | 1.5E-14 | 60.2 |
| BMI | Men | rs11121210 | T | C | 0.342 | -0.072 | 0.012 | 7.9E-09 | 32.8 |
| BMI | Men | rs11581010 | A | G | 0.763 | -0.086 | 0.014 | 1.0E-09 | 37.7 |
| BMI | Men | rs2235549 | T | G | 0.840 | -0.102 | 0.016 | 8.3E-11 | 41.7 |
| BMI | Men | rs2271928 | A | G | 0.375 | -0.065 | 0.012 | 9.9E-09 | 32.1 |
| BMI | Men | rs12061406 | T | C | 0.078 | -0.131 | 0.022 | 1.7E-09 | 36.8 |
| BMI | Men | rs1768808 | T | C | 0.624 | 0.069 | 0.012 | 3.5E-08 | 30.3 |
| BMI | Men | rs977747 | T | G | 0.439 | 0.076 | 0.012 | 1.3E-11 | 43.9 |
| BMI | Men | rs7531656 | A | G | 0.330 | 0.095 | 0.012 | 2.3E-15 | 62.7 |
| BMI | Men | rs2481665 | T | C | 0.583 | 0.090 | 0.012 | 2.0E-15 | 60.7 |
| BMI | Men | rs2310754 | A | G | 0.510 | 0.075 | 0.012 | 4.4E-10 | 38.4 |
| BMI | Men | rs1993709 | A | G | 0.178 | -0.199 | 0.014 | 1.5E-42 | 189.5 |
| BMI | Men | rs12566985 | A | G | 0.558 | -0.087 | 0.011 | 1.1E-14 | 61.9 |
| BMI | Men | rs12049202 | T | C | 0.186 | 0.109 | 0.015 | 5.5E-13 | 53.1 |
| BMI | Men | rs12034762 | T | C | 0.392 | 0.081 | 0.012 | 4.8E-11 | 41.8 |
| BMI | Men | rs11165643 | T | C | 0.579 | 0.097 | 0.012 | 1.7E-17 | 70.1 |
| BMI | Men | rs17024393 | T | C | 0.968 | -0.342 | 0.034 | 1.7E-23 | 100.3 |
| BMI | Men | rs6587552 | A | G | 0.241 | 0.086 | 0.014 | 1.0E-09 | 37.7 |
| BMI | Men | rs543874 | A | G | 0.775 | -0.173 | 0.014 | 3.7E-35 | 154.1 |
| BMI | Men | rs10920678 | A | G | 0.422 | 0.064 | 0.012 | 1.9E-08 | 30.7 |
| BMI | Men | rs2172935 | T | C | 0.321 | 0.128 | 0.012 | 3.6E-24 | 105.5 |
| BMI | Men | rs823094 | T | G | 0.553 | 0.071 | 0.012 | 3.8E-10 | 38.0 |
| BMI | Men | rs4653942 | A | G | 0.209 | -0.077 | 0.014 | 4.4E-08 | 30.4 |
| BMI | Men | rs2125231 | T | C | 0.323 | 0.082 | 0.012 | 5.5E-11 | 43.3 |
| BMI | Men | rs4639527 | A | G | 0.704 | -0.096 | 0.013 | 2.0E-13 | 54.9 |
| BMI | Men | rs12714415 | T | C | 0.850 | 0.290 | 0.015 | 1.1E-80 | 356.3 |
| BMI | Men | rs2693826 | A | G | 0.429 | -0.079 | 0.012 | 5.0E-11 | 43.0 |
| BMI | Men | rs10182181 | A | G | 0.505 | -0.131 | 0.011 | 5.5E-32 | 140.9 |
| BMI | Men | rs1260326 | T | C | 0.392 | -0.088 | 0.012 | 2.1E-14 | 58.1 |
| BMI | Men | rs2888172 | A | G | 0.547 | -0.068 | 0.012 | 1.4E-08 | 31.8 |
| BMI | Men | rs10185199 | A | G | 0.287 | -0.076 | 0.014 | 3.8E-08 | 30.1 |
| BMI | Men | rs7561278 | T | C | 0.764 | 0.084 | 0.015 | 1.1E-08 | 31.9 |
| BMI | Men | rs12986742 | T | C | 0.517 | -0.095 | 0.012 | 3.2E-15 | 62.1 |
| BMI | Men | rs887912 | T | C | 0.299 | 0.110 | 0.012 | 2.7E-19 | 77.6 |
| BMI | Men | rs11685481 | T | C | 0.850 | 0.095 | 0.016 | 9.0E-09 | 33.6 |
| BMI | Men | rs4581940 | T | C | 0.586 | 0.075 | 0.012 | 8.4E-10 | 38.4 |
| BMI | Men | rs7557796 | T | C | 0.361 | 0.075 | 0.012 | 2.2E-09 | 35.5 |
| BMI | Men | rs1437377 | T | G | 0.854 | -0.117 | 0.017 | 1.5E-11 | 45.9 |
| BMI | Men | rs10197031 | T | C | 0.729 | -0.081 | 0.013 | 7.7E-10 | 38.7 |
| BMI | Men | rs902695 | A | G | 0.492 | -0.072 | 0.012 | 2.3E-09 | 35.5 |
| BMI | Men | rs11687120 | A | G | 0.446 | 0.066 | 0.012 | 4.7E-08 | 30.0 |

| | | | | | | | | | |
|---|---|---|---|---|---|---|---|---|---|
| BMI | Men | rs961360 | A | G | 0.890 | 0.102 | 0.016 | 3.8E-10 | 39.2 |
| BMI | Men | rs453520 | T | C | 0.566 | -0.077 | 0.012 | 1.2E-10 | 41.5 |
| BMI | Men | rs908671 | C | G | 0.709 | 0.077 | 0.013 | 5.8E-09 | 33.1 |
| BMI | Men | rs1019612 | T | C | 0.664 | 0.086 | 0.012 | 1.4E-11 | 47.4 |
| BMI | Men | rs10497807 | C | G | 0.502 | -0.070 | 0.011 | 5.0E-10 | 40.3 |
| BMI | Men | rs7569376 | T | C | 0.207 | 0.094 | 0.014 | 1.3E-10 | 42.3 |
| BMI | Men | rs17203016 | A | G | 0.802 | -0.082 | 0.014 | 9.9E-09 | 32.1 |
| BMI | Men | rs7599312 | A | G | 0.275 | -0.105 | 0.012 | 1.5E-16 | 70.3 |
| BMI | Men | rs1541777 | A | G | 0.551 | -0.075 | 0.011 | 1.8E-11 | 46.0 |
| BMI | Men | rs7600417 | A | G | 0.853 | 0.100 | 0.017 | 2.9E-09 | 35.3 |
| BMI | Men | rs11692572 | C | G | 0.596 | -0.067 | 0.012 | 3.7E-08 | 30.9 |
| BMI | Men | rs2596902 | A | G | 0.650 | -0.072 | 0.012 | 1.3E-08 | 33.3 |
| BMI | Men | rs6804842 | A | G | 0.430 | -0.087 | 0.012 | 1.3E-14 | 57.5 |
| BMI | Men | rs762318 | A | C | 0.628 | 0.074 | 0.012 | 2.1E-09 | 34.6 |
| BMI | Men | rs1524887 | T | C | 0.419 | -0.067 | 0.012 | 3.6E-08 | 30.9 |
| BMI | Men | rs6803741 | T | C | 0.664 | -0.072 | 0.013 | 2.0E-08 | 30.9 |
| BMI | Men | rs2188151 | T | G | 0.415 | 0.137 | 0.012 | 2.7E-29 | 129.0 |
| BMI | Men | rs6787805 | A | G | 0.139 | -0.099 | 0.017 | 2.3E-09 | 34.6 |
| BMI | Men | rs2365389 | T | C | 0.395 | -0.085 | 0.012 | 5.3E-14 | 54.4 |
| BMI | Men | rs11915371 | A | C | 0.804 | -0.105 | 0.015 | 1.1E-12 | 49.5 |
| BMI | Men | rs13078509 | T | C | 0.028 | 0.231 | 0.040 | 8.4E-09 | 32.8 |
| BMI | Men | rs3849570 | A | C | 0.354 | 0.072 | 0.012 | 2.6E-09 | 35.5 |
| BMI | Men | rs12495178 | T | C | 0.641 | 0.109 | 0.012 | 1.5E-20 | 88.7 |
| BMI | Men | rs7628689 | A | G | 0.169 | -0.110 | 0.016 | 1.4E-11 | 45.4 |
| BMI | Men | rs1492014 | T | C | 0.568 | -0.102 | 0.012 | 1.6E-17 | 72.6 |
| BMI | Men | rs1436351 | T | G | 0.737 | 0.086 | 0.013 | 3.4E-11 | 43.5 |
| BMI | Men | rs6768228 | A | G | 0.632 | -0.072 | 0.012 | 6.8E-09 | 32.8 |
| BMI | Men | rs2918217 | T | C | 0.153 | 0.106 | 0.017 | 7.2E-10 | 37.7 |
| BMI | Men | rs1320903 | A | G | 0.309 | 0.123 | 0.013 | 1.7E-21 | 89.2 |
| BMI | Men | rs645040 | T | G | 0.770 | 0.090 | 0.013 | 3.2E-11 | 44.6 |
| BMI | Men | rs2035935 | A | G | 0.924 | -0.160 | 0.022 | 4.0E-13 | 52.4 |
| BMI | Men | rs355777 | C | G | 0.397 | 0.082 | 0.012 | 1.8E-11 | 46.2 |
| BMI | Men | rs9870575 | A | G | 0.141 | -0.114 | 0.017 | 1.5E-11 | 45.9 |
| BMI | Men | rs5398 | A | G | 0.283 | 0.081 | 0.012 | 5.5E-11 | 41.8 |
| BMI | Men | rs2606227 | T | C | 0.368 | 0.068 | 0.012 | 4.6E-08 | 29.8 |
| BMI | Men | rs9816226 | A | T | 0.174 | -0.164 | 0.015 | 5.1E-29 | 121.0 |
| BMI | Men | rs6841761 | T | G | 0.521 | -0.064 | 0.011 | 9.0E-09 | 33.9 |
| BMI | Men | rs2242189 | T | C | 0.633 | 0.082 | 0.012 | 5.6E-11 | 42.8 |
| BMI | Men | rs13130484 | T | C | 0.431 | 0.169 | 0.012 | 6.4E-50 | 215.1 |
| BMI | Men | rs2237025 | T | C | 0.430 | 0.068 | 0.012 | 3.1E-08 | 31.8 |
| BMI | Men | rs13107325 | T | C | 0.085 | 0.238 | 0.023 | 3.2E-26 | 110.9 |
| BMI | Men | rs2613726 | A | G | 0.370 | -0.078 | 0.012 | 4.3E-10 | 38.8 |
| BMI | Men | rs2391518 | T | C | 0.315 | 0.076 | 0.012 | 1.7E-10 | 40.4 |
| BMI | Men | rs1296328 | A | C | 0.443 | 0.096 | 0.012 | 2.1E-15 | 64.0 |
| BMI | Men | rs6537285 | A | G | 0.280 | -0.086 | 0.013 | 1.2E-10 | 40.4 |
| BMI | Men | rs6827083 | A | G | 0.579 | -0.073 | 0.012 | 1.3E-10 | 40.1 |
| BMI | Men | rs828544 | T | G | 0.435 | 0.066 | 0.012 | 4.5E-08 | 30.0 |
| BMI | Men | rs1503526 | T | C | 0.530 | -0.067 | 0.012 | 5.7E-09 | 34.0 |

| | | | | | | | | | |
|---|---|---|---|---|---|---|---|---|---|
| BMI | Men | rs253414 | T | C | 0.657 | 0.125 | 0.012 | 1.1E-24 | 107.3 |
| BMI | Men | rs4532349 | A | G | 0.783 | 0.078 | 0.013 | 6.6E-09 | 33.5 |
| BMI | Men | rs7733438 | T | G | 0.859 | -0.166 | 0.017 | 2.1E-21 | 91.8 |
| BMI | Men | rs6235 | C | G | 0.734 | -0.100 | 0.013 | 1.9E-13 | 54.7 |
| BMI | Men | rs11951673 | T | C | 0.395 | -0.090 | 0.012 | 7.8E-15 | 60.7 |
| BMI | Men | rs289228 | C | G | 0.836 | 0.107 | 0.016 | 9.8E-12 | 45.3 |
| BMI | Men | rs7711753 | A | G | 0.416 | -0.063 | 0.012 | 2.6E-08 | 30.3 |
| BMI | Men | rs7715256 | T | G | 0.563 | -0.078 | 0.012 | 4.5E-12 | 46.1 |
| BMI | Men | rs7730898 | A | G | 0.731 | 0.081 | 0.012 | 9.9E-11 | 42.3 |
| BMI | Men | rs6556301 | T | G | 0.370 | -0.069 | 0.012 | 1.6E-08 | 32.7 |
| BMI | Men | rs2228213 | A | G | 0.337 | -0.073 | 0.012 | 6.8E-10 | 36.5 |
| BMI | Men | rs9366639 | C | G | 0.804 | 0.093 | 0.016 | 3.8E-09 | 34.6 |
| BMI | Men | rs2178899 | A | T | 0.886 | 0.118 | 0.017 | 5.4E-12 | 46.7 |
| BMI | Men | rs14398 | A | G | 0.661 | -0.107 | 0.017 | 7.1E-10 | 38.0 |
| BMI | Men | rs2744974 | T | C | 0.322 | 0.110 | 0.012 | 8.5E-20 | 83.9 |
| BMI | Men | rs17681686 | C | G | 0.295 | 0.101 | 0.013 | 5.3E-15 | 60.5 |
| BMI | Men | rs1358980 | T | C | 0.472 | -0.075 | 0.012 | 6.1E-11 | 42.8 |
| BMI | Men | rs2206277 | T | C | 0.151 | 0.218 | 0.015 | 5.4E-50 | 214.5 |
| BMI | Men | rs1327259 | A | G | 0.610 | 0.076 | 0.012 | 6.0E-10 | 37.4 |
| BMI | Men | rs7773045 | T | G | 0.922 | 0.127 | 0.021 | 1.6E-09 | 36.0 |
| BMI | Men | rs12663742 | A | G | 0.372 | -0.087 | 0.012 | 1.7E-12 | 48.5 |
| BMI | Men | rs10872224 | T | G | 0.585 | 0.097 | 0.012 | 3.8E-15 | 59.8 |
| BMI | Men | rs17789218 | T | C | 0.753 | -0.073 | 0.013 | 3.3E-08 | 29.5 |
| BMI | Men | rs768023 | A | G | 0.602 | 0.064 | 0.012 | 3.0E-08 | 30.7 |
| BMI | Men | rs1159974 | T | C | 0.492 | -0.067 | 0.012 | 2.0E-08 | 30.9 |
| BMI | Men | rs6569648 | T | C | 0.765 | -0.073 | 0.013 | 3.7E-08 | 29.5 |
| BMI | Men | rs765875 | T | C | 0.466 | -0.066 | 0.012 | 3.4E-08 | 30.5 |
| BMI | Men | rs13191362 | A | G | 0.853 | 0.127 | 0.017 | 2.3E-13 | 54.2 |
| BMI | Men | rs9364687 | T | G | 0.398 | -0.067 | 0.012 | 4.0E-09 | 34.0 |
| BMI | Men | rs9638713 | A | G | 0.027 | 0.216 | 0.036 | 2.3E-09 | 36.0 |
| BMI | Men | rs215635 | T | C | 0.622 | -0.088 | 0.012 | 1.2E-12 | 49.5 |
| BMI | Men | rs7779853 | A | G | 0.461 | -0.063 | 0.012 | 2.8E-08 | 29.8 |
| BMI | Men | rs10269783 | A | G | 0.423 | 0.083 | 0.012 | 7.6E-13 | 51.4 |
| BMI | Men | rs1167827 | A | G | 0.439 | -0.104 | 0.012 | 2.9E-19 | 81.8 |
| BMI | Men | rs2245368 | T | C | 0.810 | -0.158 | 0.016 | 4.4E-23 | 98.8 |
| BMI | Men | rs1965529 | A | G | 0.771 | 0.084 | 0.015 | 4.2E-08 | 29.9 |
| BMI | Men | rs1701829 | A | G | 0.336 | 0.103 | 0.012 | 6.8E-16 | 67.7 |
| BMI | Men | rs13228237 | C | G | 0.213 | -0.079 | 0.014 | 4.6E-08 | 29.9 |
| BMI | Men | rs4841504 | A | C | 0.502 | -0.093 | 0.012 | 4.2E-16 | 64.7 |
| BMI | Men | rs4123853 | T | C | 0.373 | 0.080 | 0.012 | 1.1E-10 | 40.8 |
| BMI | Men | rs4258002 | T | G | 0.141 | -0.089 | 0.016 | 2.0E-08 | 31.4 |
| BMI | Men | rs7827210 | A | G | 0.397 | 0.086 | 0.012 | 2.3E-12 | 47.4 |
| BMI | Men | rs7012923 | A | C | 0.441 | 0.078 | 0.012 | 4.9E-11 | 42.5 |
| BMI | Men | rs2017657 | T | G | 0.559 | -0.074 | 0.012 | 7.0E-10 | 37.9 |
| BMI | Men | rs2588785 | A | G | 0.284 | -0.101 | 0.012 | 1.7E-15 | 65.9 |
| BMI | Men | rs957447 | A | C | 0.239 | -0.108 | 0.014 | 8.6E-14 | 56.3 |
| BMI | Men | rs800571 | T | C | 0.246 | -0.087 | 0.014 | 6.9E-10 | 39.0 |
| BMI | Men | rs11781699 | T | C | 0.811 | -0.085 | 0.014 | 6.7E-09 | 34.4 |

| | | | | | | | | | |
|---|---|---|---|---|---|---|---|---|---|
| BMI | Men | rs7826388 | T | C | 0.037 | -0.180 | 0.032 | 1.4E-08 | 32.1 |
| BMI | Men | rs4740619 | T | C | 0.542 | 0.090 | 0.011 | 1.5E-15 | 66.1 |
| BMI | Men | rs10738451 | T | C | 0.715 | -0.078 | 0.013 | 2.2E-09 | 36.4 |
| BMI | Men | rs10968114 | A | C | 0.534 | 0.074 | 0.012 | 8.7E-10 | 37.5 |
| BMI | Men | rs1412235 | C | G | 0.314 | 0.112 | 0.012 | 9.9E-21 | 87.6 |
| BMI | Men | rs10746862 | T | G | 0.580 | -0.075 | 0.012 | 8.6E-10 | 38.4 |
| BMI | Men | rs1187351 | A | C | 0.399 | -0.067 | 0.012 | 4.5E-08 | 30.9 |
| BMI | Men | rs6559921 | A | T | 0.771 | 0.087 | 0.014 | 1.2E-09 | 36.4 |
| BMI | Men | rs6477694 | T | C | 0.642 | -0.067 | 0.012 | 8.9E-09 | 34.0 |
| BMI | Men | rs7038943 | T | C | 0.659 | 0.073 | 0.012 | 6.5E-09 | 34.2 |
| BMI | Men | rs10760279 | T | G | 0.383 | 0.074 | 0.013 | 7.5E-09 | 32.5 |
| BMI | Men | rs3902840 | A | G | 0.087 | 0.148 | 0.021 | 2.5E-12 | 48.7 |
| BMI | Men | rs4740383 | A | G | 0.418 | 0.074 | 0.012 | 2.7E-09 | 35.1 |
| BMI | Men | rs11251291 | A | C | 0.467 | -0.068 | 0.012 | 1.7E-08 | 31.8 |
| BMI | Men | rs10828247 | A | G | 0.664 | -0.081 | 0.012 | 1.7E-10 | 42.3 |
| BMI | Men | rs3758438 | T | C | 0.078 | 0.132 | 0.023 | 7.3E-09 | 34.0 |
| BMI | Men | rs7072873 | T | C | 0.535 | -0.071 | 0.012 | 5.5E-09 | 34.6 |
| BMI | Men | rs10788494 | C | G | 0.483 | 0.075 | 0.012 | 5.4E-10 | 38.4 |
| BMI | Men | rs17094222 | T | C | 0.791 | -0.123 | 0.014 | 1.1E-18 | 77.3 |
| BMI | Men | rs284851 | C | G | 0.879 | -0.119 | 0.017 | 2.8E-12 | 50.2 |
| BMI | Men | rs7903146 | T | C | 0.278 | -0.111 | 0.012 | 3.6E-19 | 78.9 |
| BMI | Men | rs845084 | A | G | 0.269 | 0.075 | 0.014 | 3.8E-08 | 29.3 |
| BMI | Men | rs7113874 | T | C | 0.327 | -0.069 | 0.012 | 3.3E-09 | 36.0 |
| BMI | Men | rs900144 | T | C | 0.584 | 0.074 | 0.012 | 5.5E-10 | 37.9 |
| BMI | Men | rs6265 | T | C | 0.183 | -0.230 | 0.014 | 6.3E-58 | 253.9 |
| BMI | Men | rs1765139 | T | C | 0.767 | 0.106 | 0.014 | 1.8E-13 | 54.3 |
| BMI | Men | rs2862996 | T | G | 0.704 | -0.096 | 0.012 | 2.5E-15 | 63.4 |
| BMI | Men | rs7124681 | A | C | 0.419 | 0.132 | 0.012 | 5.1E-31 | 131.3 |
| BMI | Men | rs7102454 | T | C | 0.642 | -0.085 | 0.012 | 1.3E-11 | 46.3 |
| BMI | Men | rs349088 | A | C | 0.473 | -0.072 | 0.012 | 2.0E-09 | 36.0 |
| BMI | Men | rs647248 | A | G | 0.544 | 0.077 | 0.012 | 2.6E-10 | 41.5 |
| BMI | Men | rs12286929 | A | G | 0.507 | -0.086 | 0.011 | 2.3E-14 | 59.9 |
| BMI | Men | rs7944782 | T | G | 0.504 | -0.070 | 0.012 | 7.5E-09 | 34.1 |
| BMI | Men | rs10894670 | A | C | 0.561 | -0.068 | 0.012 | 1.6E-09 | 35.0 |
| BMI | Men | rs11611246 | T | G | 0.196 | 0.118 | 0.014 | 8.0E-17 | 67.2 |
| BMI | Men | rs10772983 | T | C | 0.544 | -0.067 | 0.011 | 2.0E-09 | 37.1 |
| BMI | Men | rs10842240 | C | G | 0.125 | 0.124 | 0.018 | 4.6E-12 | 48.6 |
| BMI | Men | rs11170468 | A | C | 0.784 | 0.077 | 0.013 | 7.8E-09 | 33.1 |
| BMI | Men | rs11181001 | A | G | 0.482 | 0.069 | 0.012 | 1.2E-09 | 36.0 |
| BMI | Men | rs7132908 | A | G | 0.392 | 0.166 | 0.013 | 1.3E-38 | 164.2 |
| BMI | Men | rs705704 | A | G | 0.322 | -0.078 | 0.012 | 1.5E-10 | 39.3 |
| BMI | Men | rs1689437 | A | G | 0.948 | -0.143 | 0.024 | 2.0E-09 | 36.7 |
| BMI | Men | rs7488867 | T | C | 0.257 | -0.106 | 0.014 | 5.7E-14 | 57.6 |
| BMI | Men | rs10850184 | T | G | 0.321 | 0.080 | 0.013 | 3.5E-10 | 38.3 |
| BMI | Men | rs12369179 | T | C | 0.084 | -0.169 | 0.022 | 1.4E-14 | 58.2 |
| BMI | Men | rs7323 | C | G | 0.267 | -0.084 | 0.013 | 4.1E-10 | 39.1 |
| BMI | Men | rs2504236 | T | C | 0.586 | -0.073 | 0.012 | 1.9E-09 | 36.5 |
| BMI | Men | rs9595893 | T | G | 0.360 | -0.096 | 0.012 | 1.2E-14 | 58.6 |

| | | | | | | | | | |
|---|---|---|---|---|---|---|---|---|---|
| BMI | Men | rs9603697 | T | C | 0.317 | 0.095 | 0.013 | 1.3E-13 | 53.2 |
| BMI | Men | rs12429545 | A | G | 0.121 | 0.162 | 0.017 | 2.0E-21 | 92.2 |
| BMI | Men | rs7338127 | A | T | 0.330 | 0.095 | 0.013 | 1.4E-13 | 53.2 |
| BMI | Men | rs1441264 | A | G | 0.582 | 0.087 | 0.012 | 1.4E-13 | 56.9 |
| BMI | Men | rs912690 | C | G | 0.557 | -0.083 | 0.012 | 6.9E-12 | 47.9 |
| BMI | Men | rs9168 | A | C | 0.273 | -0.072 | 0.013 | 4.6E-08 | 30.9 |
| BMI | Men | rs12868881 | A | T | 0.405 | 0.087 | 0.012 | 1.1E-12 | 51.8 |
| BMI | Men | rs10132280 | A | C | 0.313 | -0.144 | 0.012 | 1.4E-31 | 132.3 |
| BMI | Men | rs1950709 | A | G | 0.556 | -0.065 | 0.011 | 7.3E-09 | 35.0 |
| BMI | Men | rs1955540 | T | C | 0.194 | -0.096 | 0.015 | 2.9E-10 | 39.1 |
| BMI | Men | rs4900714 | T | G | 0.473 | 0.096 | 0.012 | 1.2E-15 | 64.0 |
| BMI | Men | rs217671 | A | G | 0.726 | -0.081 | 0.013 | 2.2E-09 | 36.4 |
| BMI | Men | rs3902951 | T | G | 0.771 | -0.073 | 0.013 | 4.4E-08 | 29.1 |
| BMI | Men | rs17109256 | A | G | 0.237 | 0.148 | 0.013 | 2.8E-27 | 121.0 |
| BMI | Men | rs12888545 | A | G | 0.745 | -0.084 | 0.014 | 2.0E-09 | 36.4 |
| BMI | Men | rs2148564 | A | G | 0.285 | -0.095 | 0.013 | 1.1E-12 | 50.0 |
| BMI | Men | rs7161194 | A | G | 0.345 | 0.103 | 0.013 | 3.3E-15 | 63.4 |
| BMI | Men | rs2015407 | A | G | 0.353 | 0.087 | 0.012 | 2.7E-12 | 47.9 |
| BMI | Men | rs11636536 | T | C | 0.497 | 0.071 | 0.012 | 2.8E-09 | 35.0 |
| BMI | Men | rs10851458 | A | G | 0.581 | 0.071 | 0.012 | 6.0E-09 | 35.0 |
| BMI | Men | rs4776970 | A | T | 0.643 | 0.134 | 0.012 | 1.5E-30 | 135.1 |
| BMI | Men | rs4777541 | T | C | 0.784 | 0.101 | 0.014 | 1.1E-12 | 49.0 |
| BMI | Men | rs8024572 | A | G | 0.611 | 0.075 | 0.012 | 7.8E-10 | 38.9 |
| BMI | Men | rs8036171 | A | C | 0.374 | 0.074 | 0.012 | 4.3E-10 | 37.5 |
| BMI | Men | rs12448257 | A | G | 0.219 | 0.089 | 0.014 | 1.2E-10 | 41.1 |
| BMI | Men | rs879620 | T | C | 0.597 | 0.100 | 0.012 | 3.1E-16 | 64.6 |
| BMI | Men | rs9926784 | T | C | 0.801 | 0.108 | 0.014 | 5.7E-14 | 55.8 |
| BMI | Men | rs9921854 | A | G | 0.087 | -0.140 | 0.022 | 1.9E-10 | 40.3 |
| BMI | Men | rs3888190 | A | C | 0.380 | 0.151 | 0.012 | 7.0E-40 | 172.3 |
| BMI | Men | rs7201780 | T | C | 0.454 | -0.107 | 0.012 | 1.4E-18 | 79.6 |
| BMI | Men | rs1558902 | A | T | 0.415 | 0.392 | 0.012 | 1.2E-257 | 1153.2 |
| BMI | Men | rs889398 | T | C | 0.418 | -0.100 | 0.012 | 1.5E-18 | 75.8 |
| BMI | Men | rs756717 | A | G | 0.394 | -0.068 | 0.012 | 8.6E-09 | 34.5 |
| BMI | Men | rs7206608 | C | G | 0.674 | -0.072 | 0.013 | 1.9E-08 | 30.9 |
| BMI | Men | rs3923783 | A | C | 0.178 | -0.109 | 0.016 | 7.5E-12 | 46.9 |
| BMI | Men | rs12450045 | A | G | 0.458 | 0.069 | 0.012 | 1.7E-08 | 32.7 |
| BMI | Men | rs4986044 | T | C | 0.454 | -0.087 | 0.012 | 1.5E-14 | 56.9 |
| BMI | Men | rs8065172 | A | G | 0.238 | -0.080 | 0.014 | 1.2E-08 | 33.2 |
| BMI | Men | rs12150665 | T | C | 0.593 | 0.087 | 0.012 | 3.4E-14 | 56.9 |
| BMI | Men | rs676387 | A | C | 0.303 | 0.078 | 0.013 | 4.1E-09 | 36.0 |
| BMI | Men | rs208015 | T | C | 0.070 | 0.175 | 0.024 | 8.7E-14 | 55.5 |
| BMI | Men | rs12939549 | A | G | 0.556 | 0.096 | 0.011 | 1.3E-17 | 75.6 |
| BMI | Men | rs7226371 | A | G | 0.844 | -0.089 | 0.015 | 7.7E-09 | 33.8 |
| BMI | Men | rs891387 | T | C | 0.496 | 0.109 | 0.012 | 6.8E-20 | 82.4 |
| BMI | Men | rs11874191 | T | C | 0.728 | -0.081 | 0.014 | 3.1E-09 | 34.0 |
| BMI | Men | rs2612575 | A | G | 0.276 | -0.077 | 0.013 | 1.5E-08 | 32.7 |
| BMI | Men | rs9951619 | T | G | 0.222 | -0.089 | 0.013 | 1.7E-11 | 44.1 |
| BMI | Men | rs6567160 | T | C | 0.749 | -0.256 | 0.013 | 2.1E-82 | 362.4 |

| | | | | | | | | | |
|---|---|---|---|---|---|---|---|---|---|
| BMI | Men | rs17066856 | T | C | 0.890 | 0.145 | 0.019 | 2.7E-14 | 59.6 |
| BMI | Men | rs17710386 | T | C | 0.664 | -0.079 | 0.012 | 3.7E-11 | 43.0 |
| BMI | Men | rs1787267 | C | G | 0.063 | -0.157 | 0.025 | 2.5E-10 | 39.5 |
| BMI | Men | rs7258309 | T | G | 0.784 | 0.112 | 0.015 | 6.5E-14 | 56.5 |
| BMI | Men | rs273512 | T | C | 0.410 | 0.079 | 0.012 | 1.7E-10 | 39.8 |
| BMI | Men | rs9636202 | A | G | 0.281 | -0.085 | 0.013 | 5.5E-11 | 43.0 |
| BMI | Men | rs7258722 | A | T | 0.411 | -0.096 | 0.012 | 8.6E-15 | 58.6 |
| BMI | Men | rs17513613 | T | C | 0.695 | -0.086 | 0.012 | 1.5E-12 | 51.3 |
| BMI | Men | rs11084553 | A | G | 0.849 | 0.114 | 0.017 | 9.4E-12 | 45.9 |
| BMI | Men | rs769449 | A | G | 0.102 | -0.116 | 0.019 | 1.6E-09 | 36.6 |
| BMI | Men | rs1800437 | C | G | 0.186 | -0.175 | 0.014 | 1.7E-33 | 148.0 |
| BMI | Men | rs9304665 | A | T | 0.741 | 0.132 | 0.014 | 4.8E-21 | 89.3 |
| BMI | Men | rs7249149 | A | G | 0.696 | 0.073 | 0.013 | 4.4E-08 | 29.5 |
| BMI | Men | rs2145270 | T | C | 0.616 | 0.100 | 0.012 | 7.0E-18 | 75.1 |
| BMI | Men | rs8123881 | A | G | 0.864 | -0.092 | 0.016 | 2.5E-08 | 31.9 |
| BMI | Men | rs7272093 | A | G | 0.700 | -0.083 | 0.013 | 1.1E-09 | 37.7 |
| BMI | Men | rs742760 | A | T | 0.820 | 0.112 | 0.015 | 7.1E-13 | 53.0 |
| BMI | Men | rs6092143 | T | C | 0.368 | 0.072 | 0.012 | 1.1E-08 | 32.8 |
| BMI | Men | rs13047416 | C | G | 0.628 | 0.075 | 0.012 | 9.8E-10 | 36.5 |
| BMI | Men | rs427943 | A | C | 0.427 | -0.084 | 0.012 | 2.2E-12 | 49.0 |
| BMI | Men | rs138383 | T | C | 0.514 | -0.075 | 0.012 | 1.4E-09 | 35.5 |
| BMI | Men | rs733381 | A | G | 0.767 | 0.099 | 0.014 | 5.2E-13 | 50.5 |
| HbA1c | Overall | rs11191561 | G | C | 0.150 | 0.122 | 0.022 | 2.3E-08 | 31.3 |
| HbA1c | Overall | rs11195508 | G | C | 0.149 | -0.119 | 0.022 | 4.3E-08 | 30.0 |
| HbA1c | Overall | rs114322470 | G | T | 0.019 | -0.363 | 0.057 | 2.2E-10 | 40.3 |
| HbA1c | Overall | rs34872471 | C | T | 0.291 | 0.485 | 0.017 | 3.5E-177 | 806.4 |
| HbA1c | Overall | rs10736269 | C | T | 0.471 | -0.098 | 0.016 | 4.5E-10 | 38.9 |
| HbA1c | Overall | rs11257655 | T | C | 0.208 | 0.245 | 0.019 | 1.5E-37 | 164.1 |
| HbA1c | Overall | rs7904973 | T | G | 0.576 | 0.093 | 0.016 | 3.0E-09 | 35.2 |
| HbA1c | Overall | rs11598987 | G | C | 0.527 | 0.088 | 0.016 | 1.8E-08 | 31.7 |
| HbA1c | Overall | rs3847378 | G | A | 0.264 | -0.116 | 0.018 | 5.3E-11 | 43.1 |
| HbA1c | Overall | rs61850681 | A | G | 0.239 | -0.227 | 0.018 | 3.2E-35 | 153.4 |
| HbA1c | Overall | rs561279660 | T | C | 0.015 | -0.690 | 0.068 | 3.7E-24 | 102.8 |
| HbA1c | Overall | rs72814244 | G | C | 0.052 | 0.330 | 0.035 | 3.5E-21 | 89.2 |
| HbA1c | Overall | rs150025254 | T | C | 0.030 | -1.049 | 0.046 | 4.5E-114 | 515.7 |
| HbA1c | Overall | rs75048187 | G | A | 0.013 | 0.610 | 0.070 | 3.6E-18 | 75.6 |
| HbA1c | Overall | rs6480398 | G | A | 0.906 | 1.273 | 0.027 | 0.0E+00 | 2171.4 |
| HbA1c | Overall | rs555721560 | G | T | 0.022 | 0.310 | 0.056 | 2.5E-08 | 31.1 |
| HbA1c | Overall | rs138691176 | A | G | 0.033 | 0.267 | 0.045 | 4.0E-09 | 34.6 |
| HbA1c | Overall | rs1248673 | C | T | 0.293 | 0.125 | 0.017 | 2.3E-13 | 53.7 |
| HbA1c | Overall | rs701848 | C | T | 0.390 | -0.121 | 0.016 | 3.3E-14 | 57.5 |
| HbA1c | Overall | rs7923837 | A | G | 0.380 | -0.196 | 0.016 | 1.7E-34 | 150.0 |
| HbA1c | Overall | rs2068888 | A | G | 0.449 | -0.116 | 0.016 | 1.2E-13 | 55.0 |
| HbA1c | Overall | rs12365580 | A | G | 0.151 | -0.218 | 0.022 | 1.3E-23 | 100.3 |
| HbA1c | Overall | rs11042852 | G | A | 0.087 | 0.269 | 0.028 | 3.1E-22 | 94.0 |
| HbA1c | Overall | rs3802885 | C | A | 0.075 | -0.206 | 0.030 | 3.0E-12 | 48.7 |
| HbA1c | Overall | rs6589939 | G | A | 0.379 | 0.112 | 0.016 | 2.9E-12 | 48.7 |
| HbA1c | Overall | rs11512059 | T | C | 0.083 | 0.184 | 0.028 | 6.5E-11 | 42.7 |

| | | | | | | | | | |
|---|---|---|---|---|---|---|---|---|---|
| HbA1c | Overall | rs7124355 | G | A | 0.675 | -0.125 | 0.017 | 5.0E-14 | 56.7 |
| HbA1c | Overall | rs689 | T | A | 0.710 | -0.148 | 0.017 | 9.1E-18 | 73.7 |
| HbA1c | Overall | rs12419995 | T | A | 0.656 | -0.205 | 0.017 | 7.2E-33 | 142.6 |
| HbA1c | Overall | rs2237895 | C | A | 0.415 | 0.185 | 0.016 | 5.3E-32 | 138.7 |
| HbA1c | Overall | rs10047488 | C | G | 0.083 | 0.177 | 0.029 | 6.4E-10 | 38.2 |
| HbA1c | Overall | rs4755309 | T | G | 0.114 | -0.171 | 0.024 | 2.9E-12 | 48.8 |
| HbA1c | Overall | rs10501320 | C | G | 0.264 | -0.182 | 0.018 | 4.9E-25 | 106.8 |
| HbA1c | Overall | rs174549 | A | G | 0.310 | -0.167 | 0.017 | 2.1E-23 | 99.4 |
| HbA1c | Overall | rs12294913 | G | C | 0.050 | 0.281 | 0.037 | 3.9E-14 | 57.2 |
| HbA1c | Overall | rs1152620 | G | A | 0.262 | 0.120 | 0.018 | 1.1E-11 | 46.2 |
| HbA1c | Overall | rs11602873 | T | A | 0.157 | -0.236 | 0.021 | 1.2E-28 | 123.4 |
| HbA1c | Overall | rs689419 | G | T | 0.782 | 0.114 | 0.019 | 1.6E-09 | 36.4 |
| HbA1c | Overall | rs10830963 | G | C | 0.275 | 0.435 | 0.017 | 7.8E-139 | 629.7 |
| HbA1c | Overall | rs3020069 | A | G | 0.683 | 0.106 | 0.017 | 1.7E-10 | 40.8 |
| HbA1c | Overall | rs4910498 | T | A | 0.643 | -0.224 | 0.016 | 2.4E-43 | 190.6 |
| HbA1c | Overall | rs7310615 | G | C | 0.517 | 0.193 | 0.016 | 5.9E-35 | 152.2 |
| HbA1c | Overall | rs2936839 | C | T | 0.610 | -0.105 | 0.016 | 4.9E-11 | 43.2 |
| HbA1c | Overall | rs73226260 | A | G | 0.034 | -0.286 | 0.043 | 3.2E-11 | 44.1 |
| HbA1c | Overall | rs11043299 | C | T | 0.621 | 0.104 | 0.017 | 1.5E-09 | 36.6 |
| HbA1c | Overall | rs7306544 | C | T | 0.104 | 0.146 | 0.025 | 9.6E-09 | 32.9 |
| HbA1c | Overall | rs704168 | G | A | 0.184 | -0.123 | 0.020 | 9.7E-10 | 37.4 |
| HbA1c | Overall | rs10492373 | A | G | 0.196 | -0.177 | 0.019 | 1.3E-19 | 82.1 |
| HbA1c | Overall | rs4238013 | T | C | 0.796 | -0.122 | 0.019 | 3.6E-10 | 39.3 |
| HbA1c | Overall | rs76895963 | G | T | 0.020 | -1.275 | 0.060 | 2.1E-101 | 457.4 |
| HbA1c | Overall | rs12819124 | A | C | 0.472 | -0.417 | 0.016 | 2.3E-158 | 719.7 |
| HbA1c | Overall | rs6580661 | T | A | 0.224 | 0.227 | 0.019 | 7.3E-34 | 147.2 |
| HbA1c | Overall | rs11169605 | C | T | 0.358 | 0.125 | 0.016 | 1.3E-14 | 59.3 |
| HbA1c | Overall | rs765634 | G | A | 0.367 | -0.090 | 0.016 | 3.5E-08 | 30.4 |
| HbA1c | Overall | rs79755767 | A | G | 0.100 | -0.243 | 0.026 | 1.2E-20 | 86.7 |
| HbA1c | Overall | rs4760278 | A | C | 0.226 | -0.128 | 0.019 | 4.1E-12 | 48.1 |
| HbA1c | Overall | rs2258238 | T | A | 0.105 | 0.218 | 0.025 | 8.2E-18 | 73.9 |
| HbA1c | Overall | rs2137537 | C | T | 0.546 | 0.090 | 0.016 | 1.0E-08 | 32.8 |
| HbA1c | Overall | rs12813389 | T | A | 0.539 | 0.091 | 0.016 | 5.5E-09 | 34.0 |
| HbA1c | Overall | rs79621919 | A | G | 0.059 | -0.261 | 0.033 | 2.9E-15 | 62.3 |
| HbA1c | Overall | rs1044364 | C | T | 0.056 | -0.191 | 0.034 | 2.0E-08 | 31.5 |
| HbA1c | Overall | rs12876143 | C | T | 0.089 | 0.554 | 0.027 | 1.3E-91 | 412.4 |
| HbA1c | Overall | rs368865 | G | A | 0.725 | 0.189 | 0.017 | 1.1E-27 | 119.0 |
| HbA1c | Overall | rs7329468 | T | C | 0.217 | -0.256 | 0.019 | 1.9E-41 | 181.9 |
| HbA1c | Overall | rs4769580 | A | T | 0.219 | 0.224 | 0.019 | 1.9E-32 | 140.6 |
| HbA1c | Overall | rs75629141 | A | T | 0.829 | -0.209 | 0.021 | 5.8E-24 | 101.9 |
| HbA1c | Overall | rs1327315 | T | C | 0.289 | -0.134 | 0.017 | 5.7E-15 | 61.0 |
| HbA1c | Overall | rs61990662 | T | C | 0.173 | -0.127 | 0.021 | 9.5E-10 | 37.4 |
| HbA1c | Overall | rs9324022 | C | T | 0.206 | 0.163 | 0.020 | 6.4E-17 | 69.9 |
| HbA1c | Overall | rs12147688 | T | G | 0.258 | 0.145 | 0.018 | 4.5E-16 | 66.0 |
| HbA1c | Overall | rs8013143 | G | A | 0.278 | 0.139 | 0.017 | 1.3E-15 | 64.0 |
| HbA1c | Overall | rs7151822 | C | T | 0.247 | -0.164 | 0.018 | 9.6E-20 | 82.7 |
| HbA1c | Overall | rs72681698 | C | T | 0.011 | -0.497 | 0.074 | 2.3E-11 | 44.7 |
| HbA1c | Overall | rs11621337 | A | C | 0.063 | -0.206 | 0.035 | 3.2E-09 | 35.0 |

| | | | | | | | | | |
|---|---|---|---|---|---|---|---|---|---|
| HbA1c | Overall | rs2285005 | A | G | 0.340 | 0.182 | 0.016 | 1.5E-28 | 122.8 |
| HbA1c | Overall | rs35889227 | T | G | 0.631 | -0.150 | 0.016 | 9.5E-21 | 87.3 |
| HbA1c | Overall | rs34715063 | C | T | 0.130 | 0.145 | 0.023 | 4.1E-10 | 39.1 |
| HbA1c | Overall | rs28526689 | A | G | 0.344 | 0.101 | 0.016 | 6.3E-10 | 38.2 |
| HbA1c | Overall | rs4516170 | C | T | 0.050 | -0.325 | 0.036 | 6.6E-20 | 83.4 |
| HbA1c | Overall | rs1550026 | T | A | 0.408 | 0.145 | 0.016 | 6.4E-20 | 83.5 |
| HbA1c | Overall | rs59153558 | G | A | 0.720 | 0.200 | 0.017 | 1.0E-30 | 132.8 |
| HbA1c | Overall | rs11638511 | G | A | 0.561 | 0.091 | 0.016 | 5.9E-09 | 33.9 |
| HbA1c | Overall | rs7166540 | G | A | 0.238 | -0.127 | 0.018 | 5.0E-12 | 47.7 |
| HbA1c | Overall | rs1866476 | C | T | 0.732 | -0.141 | 0.018 | 7.9E-16 | 64.9 |
| HbA1c | Overall | rs61215894 | C | T | 0.558 | -0.096 | 0.016 | 2.0E-09 | 36.0 |
| HbA1c | Overall | rs4451969 | C | T | 0.671 | -0.139 | 0.017 | 3.6E-17 | 71.0 |
| HbA1c | Overall | rs35591 | G | C | 0.233 | 0.136 | 0.018 | 1.1E-13 | 55.3 |
| HbA1c | Overall | rs112414002 | T | C | 0.026 | 0.281 | 0.049 | 1.0E-08 | 32.8 |
| HbA1c | Overall | rs181207 | T | C | 0.336 | 0.164 | 0.016 | 2.5E-23 | 99.1 |
| HbA1c | Overall | rs2858010 | C | T | 0.340 | -0.169 | 0.016 | 1.1E-24 | 105.2 |
| HbA1c | Overall | rs4889490 | T | G | 0.390 | -0.108 | 0.016 | 1.0E-11 | 46.2 |
| HbA1c | Overall | rs8050500 | C | T | 0.446 | -0.180 | 0.016 | 1.1E-30 | 132.6 |
| HbA1c | Overall | rs1121980 | A | G | 0.422 | 0.146 | 0.016 | 1.6E-20 | 86.3 |
| HbA1c | Overall | rs35158985 | G | A | 0.308 | 0.159 | 0.017 | 2.9E-21 | 89.6 |
| HbA1c | Overall | rs72802365 | C | G | 0.079 | -0.193 | 0.029 | 2.3E-11 | 44.7 |
| HbA1c | Overall | rs247826 | T | C | 0.223 | 0.185 | 0.019 | 4.6E-23 | 97.8 |
| HbA1c | Overall | rs72811487 | A | G | 0.035 | -0.386 | 0.042 | 4.3E-20 | 84.3 |
| HbA1c | Overall | rs7195055 | T | C | 0.015 | -0.392 | 0.065 | 1.8E-09 | 36.1 |
| HbA1c | Overall | rs551118 | G | C | 0.578 | 0.382 | 0.016 | 9.0E-127 | 574.1 |
| HbA1c | Overall | rs11655029 | C | T | 0.695 | -0.223 | 0.017 | 1.0E-39 | 174.0 |
| HbA1c | Overall | rs12943904 | C | G | 0.306 | -0.097 | 0.017 | 7.6E-09 | 33.4 |
| HbA1c | Overall | rs4985997 | T | C | 0.350 | -0.136 | 0.016 | 7.0E-17 | 69.7 |
| HbA1c | Overall | rs216195 | G | T | 0.303 | 0.137 | 0.017 | 4.8E-16 | 65.9 |
| HbA1c | Overall | rs12450826 | T | A | 0.293 | -0.186 | 0.017 | 1.7E-26 | 113.5 |
| HbA1c | Overall | rs9894558 | A | T | 0.270 | -0.115 | 0.018 | 4.2E-10 | 39.0 |
| HbA1c | Overall | rs10908278 | A | T | 0.515 | -0.123 | 0.016 | 3.5E-15 | 62.0 |
| HbA1c | Overall | rs150325706 | A | T | 0.262 | 0.131 | 0.019 | 1.1E-11 | 46.2 |
| HbA1c | Overall | rs7222851 | C | T | 0.512 | -0.105 | 0.016 | 1.4E-11 | 45.7 |
| HbA1c | Overall | rs35895680 | A | C | 0.327 | -0.096 | 0.017 | 9.1E-09 | 33.0 |
| HbA1c | Overall | rs12600858 | A | G | 0.224 | -0.203 | 0.019 | 1.4E-27 | 118.4 |
| HbA1c | Overall | rs72838857 | G | A | 0.213 | 0.144 | 0.019 | 3.4E-14 | 57.5 |
| HbA1c | Overall | rs2854215 | T | C | 0.280 | -0.113 | 0.018 | 1.4E-10 | 41.2 |
| HbA1c | Overall | rs1030097 | G | C | 0.495 | 0.106 | 0.016 | 1.2E-11 | 45.9 |
| HbA1c | Overall | rs1641523 | T | C | 0.567 | -0.186 | 0.016 | 3.0E-32 | 139.8 |
| HbA1c | Overall | rs2748427 | G | A | 0.218 | 0.540 | 0.019 | 2.9E-182 | 829.8 |
| HbA1c | Overall | rs7208422 | T | A | 0.475 | 0.117 | 0.015 | 3.5E-14 | 57.4 |
| HbA1c | Overall | rs113373052 | T | C | 0.313 | 0.467 | 0.017 | 3.5E-171 | 778.8 |
| HbA1c | Overall | rs4791641 | T | C | 0.505 | -0.148 | 0.016 | 1.3E-21 | 91.2 |
| HbA1c | Overall | rs8088001 | G | T | 0.071 | -0.168 | 0.030 | 2.8E-08 | 30.9 |
| HbA1c | Overall | rs12607898 | G | A | 0.747 | 0.164 | 0.018 | 3.9E-20 | 84.5 |
| HbA1c | Overall | rs12605978 | T | C | 0.322 | 0.093 | 0.017 | 2.5E-08 | 31.1 |
| HbA1c | Overall | rs3810027 | G | C | 0.335 | 0.096 | 0.016 | 5.9E-09 | 33.9 |

| | | | | | | | | | |
|---|---|---|---|---|---|---|---|---|---|
| HbA1c | Overall | rs2658747 | G | A | 0.379 | 0.110 | 0.016 | 7.0E-12 | 47.0 |
| HbA1c | Overall | rs11671760 | A | G | 0.206 | 0.117 | 0.019 | 1.6E-09 | 36.4 |
| HbA1c | Overall | rs56397034 | C | G | 0.391 | -0.156 | 0.016 | 8.0E-23 | 96.7 |
| HbA1c | Overall | rs4808579 | C | T | 0.392 | 0.315 | 0.016 | 2.0E-87 | 393.1 |
| HbA1c | Overall | rs4808072 | C | T | 0.581 | 0.101 | 0.016 | 1.6E-10 | 40.9 |
| HbA1c | Overall | rs75372982 | G | A | 0.289 | -0.195 | 0.017 | 2.7E-29 | 126.3 |
| HbA1c | Overall | rs72973028 | C | A | 0.172 | 0.135 | 0.021 | 7.0E-11 | 42.5 |
| HbA1c | Overall | rs17561351 | G | A | 0.064 | 0.210 | 0.032 | 3.8E-11 | 43.7 |
| HbA1c | Overall | rs10407429 | A | G | 0.426 | -0.167 | 0.016 | 1.7E-26 | 113.5 |
| HbA1c | Overall | rs62136856 | G | A | 0.717 | 0.127 | 0.017 | 1.8E-13 | 54.2 |
| HbA1c | Overall | rs12459419 | T | C | 0.326 | -0.172 | 0.017 | 2.1E-25 | 108.5 |
| HbA1c | Overall | rs2278832 | G | A | 0.205 | 0.120 | 0.020 | 2.0E-09 | 36.0 |
| HbA1c | Overall | rs45607834 | T | C | 0.103 | 0.154 | 0.026 | 2.0E-09 | 36.0 |
| HbA1c | Overall | rs351978 | G | A | 0.581 | -0.103 | 0.016 | 1.1E-10 | 41.7 |
| HbA1c | Overall | rs11166447 | C | T | 0.359 | -0.101 | 0.016 | 5.2E-10 | 38.6 |
| HbA1c | Overall | rs76757826 | C | T | 0.055 | 0.206 | 0.034 | 1.6E-09 | 36.4 |
| HbA1c | Overall | rs1975283 | C | A | 0.685 | -0.166 | 0.017 | 4.1E-23 | 98.1 |
| HbA1c | Overall | rs2487569 | T | A | 0.111 | 0.152 | 0.025 | 1.2E-09 | 36.9 |
| HbA1c | Overall | rs198325 | T | C | 0.222 | -0.184 | 0.019 | 5.6E-23 | 97.4 |
| HbA1c | Overall | rs12748814 | T | C | 0.085 | 0.174 | 0.028 | 3.5E-10 | 39.4 |
| HbA1c | Overall | rs857725 | G | T | 0.267 | 0.543 | 0.017 | 1.9E-211 | 964.4 |
| HbA1c | Overall | rs7528296 | C | T | 0.386 | -0.112 | 0.016 | 2.7E-12 | 48.9 |
| HbA1c | Overall | rs523647 | T | C | 0.463 | 0.099 | 0.017 | 3.0E-09 | 35.2 |
| HbA1c | Overall | rs2808454 | T | A | 0.542 | 0.145 | 0.016 | 1.8E-20 | 86.1 |
| HbA1c | Overall | rs1078308 | C | A | 0.727 | -0.124 | 0.017 | 1.4E-12 | 50.2 |
| HbA1c | Overall | rs340882 | G | C | 0.621 | 0.239 | 0.016 | 5.7E-50 | 221.0 |
| HbA1c | Overall | rs348330 | A | G | 0.635 | -0.147 | 0.016 | 1.1E-19 | 82.4 |
| HbA1c | Overall | rs3811444 | T | C | 0.334 | 0.166 | 0.016 | 5.7E-24 | 101.9 |
| HbA1c | Overall | rs28695210 | A | G | 0.083 | 0.267 | 0.029 | 1.5E-20 | 86.4 |
| HbA1c | Overall | rs563437995 | C | G | 0.008 | -1.076 | 0.092 | 8.8E-32 | 137.7 |
| HbA1c | Overall | rs797399 | G | A | 0.729 | -0.097 | 0.018 | 4.0E-08 | 30.2 |
| HbA1c | Overall | rs1175550 | G | A | 0.230 | -0.285 | 0.019 | 1.6E-52 | 232.7 |
| HbA1c | Overall | rs3768321 | T | G | 0.197 | 0.169 | 0.020 | 4.3E-18 | 75.2 |
| HbA1c | Overall | rs557458132 | T | C | 0.282 | -0.137 | 0.017 | 3.7E-15 | 61.9 |
| HbA1c | Overall | rs72900998 | C | T | 0.089 | -0.250 | 0.027 | 3.3E-20 | 84.8 |
| HbA1c | Overall | rs4926930 | G | A | 0.426 | -0.089 | 0.016 | 1.4E-08 | 32.2 |
| HbA1c | Overall | rs7513688 | A | G | 0.361 | 0.126 | 0.016 | 6.1E-15 | 60.9 |
| HbA1c | Overall | rs1166698 | A | G | 0.212 | -0.122 | 0.019 | 1.2E-10 | 41.5 |
| HbA1c | Overall | rs13042195 | G | A | 0.213 | -0.108 | 0.019 | 1.6E-08 | 32.0 |
| HbA1c | Overall | rs149142833 | T | C | 0.156 | 0.172 | 0.022 | 1.7E-15 | 63.3 |
| HbA1c | Overall | rs4431018 | G | A | 0.379 | 0.088 | 0.016 | 4.3E-08 | 30.0 |
| HbA1c | Overall | rs67897819 | A | G | 0.212 | 0.129 | 0.020 | 5.0E-11 | 43.2 |
| HbA1c | Overall | rs55966194 | G | C | 0.284 | -0.149 | 0.017 | 6.0E-18 | 74.5 |
| HbA1c | Overall | rs6014993 | G | A | 0.487 | 0.176 | 0.016 | 1.6E-29 | 127.3 |
| HbA1c | Overall | rs4809556 | A | G | 0.596 | -0.106 | 0.016 | 2.3E-11 | 44.7 |
| HbA1c | Overall | rs1056441 | C | T | 0.673 | 0.099 | 0.017 | 2.0E-09 | 35.9 |
| HbA1c | Overall | rs6077396 | A | G | 0.507 | -0.142 | 0.016 | 7.9E-20 | 83.1 |
| HbA1c | Overall | rs3827181 | T | C | 0.346 | 0.127 | 0.016 | 6.2E-15 | 60.8 |

| | | | | | | | | | |
|---|---|---|---|---|---|---|---|---|---|
| HbA1c | Overall | rs61138219 | T | C | 0.153 | -0.155 | 0.022 | 7.6E-13 | 51.4 |
| HbA1c | Overall | rs17850433 | C | T | 0.012 | 0.633 | 0.071 | 6.4E-19 | 79.0 |
| HbA1c | Overall | rs35963032 | T | C | 0.152 | -0.140 | 0.022 | 1.4E-10 | 41.1 |
| HbA1c | Overall | rs12628032 | T | C | 0.305 | 0.103 | 0.017 | 8.1E-10 | 37.7 |
| HbA1c | Overall | rs6518681 | G | A | 0.907 | 0.220 | 0.027 | 2.5E-16 | 67.2 |
| HbA1c | Overall | rs117721418 | T | C | 0.081 | -0.226 | 0.028 | 1.9E-15 | 63.1 |
| HbA1c | Overall | rs855791 | G | A | 0.561 | -0.422 | 0.016 | 4.1E-159 | 723.2 |
| HbA1c | Overall | rs2143918 | C | A | 0.471 | -0.186 | 0.016 | 5.3E-33 | 143.3 |
| HbA1c | Overall | rs2009581 | A | G | 0.275 | 0.096 | 0.017 | 4.5E-08 | 29.9 |
| HbA1c | Overall | rs650588 | G | A | 0.581 | -0.106 | 0.016 | 1.7E-11 | 45.3 |
| HbA1c | Overall | rs111631066 | A | G | 0.015 | -0.479 | 0.065 | 1.6E-13 | 54.5 |
| HbA1c | Overall | rs4667575 | G | C | 0.592 | 0.088 | 0.016 | 2.6E-08 | 31.0 |
| HbA1c | Overall | rs3843328 | A | G | 0.711 | -0.161 | 0.017 | 1.2E-20 | 86.9 |
| HbA1c | Overall | rs853787 | T | G | 0.651 | 0.636 | 0.016 | 0.0E+00 | 1517.2 |
| HbA1c | Overall | rs4972439 | C | T | 0.220 | -0.193 | 0.019 | 8.9E-25 | 105.7 |
| HbA1c | Overall | rs6731171 | A | T | 0.626 | 0.104 | 0.016 | 1.9E-10 | 40.5 |
| HbA1c | Overall | rs6736362 | T | C | 0.562 | 0.114 | 0.016 | 4.3E-13 | 52.5 |
| HbA1c | Overall | rs1399627 | G | A | 0.653 | 0.153 | 0.016 | 6.3E-21 | 88.1 |
| HbA1c | Overall | rs838717 | A | G | 0.566 | 0.114 | 0.016 | 3.0E-13 | 53.2 |
| HbA1c | Overall | rs72781680 | T | C | 0.135 | 0.348 | 0.023 | 1.2E-52 | 233.2 |
| HbA1c | Overall | rs13020237 | G | A | 0.182 | -0.114 | 0.020 | 1.2E-08 | 32.5 |
| HbA1c | Overall | rs780093 | C | T | 0.618 | 0.177 | 0.016 | 1.4E-28 | 123.0 |
| HbA1c | Overall | rs11124556 | T | G | 0.514 | -0.090 | 0.016 | 6.7E-09 | 33.6 |
| HbA1c | Overall | rs77981966 | T | C | 0.076 | -0.490 | 0.029 | 6.3E-63 | 280.5 |
| HbA1c | Overall | rs2164701 | C | G | 0.140 | 0.199 | 0.022 | 7.1E-19 | 78.8 |
| HbA1c | Overall | rs78583604 | T | G | 0.132 | 0.180 | 0.024 | 2.6E-14 | 58.0 |
| HbA1c | Overall | rs7606173 | C | G | 0.431 | -0.164 | 0.016 | 1.2E-25 | 109.6 |
| HbA1c | Overall | rs7572278 | A | T | 0.204 | 0.194 | 0.019 | 8.2E-24 | 101.2 |
| HbA1c | Overall | rs4435501 | C | A | 0.108 | 0.141 | 0.026 | 4.3E-08 | 30.0 |
| HbA1c | Overall | rs9826367 | G | A | 0.449 | -0.251 | 0.016 | 2.7E-58 | 259.2 |
| HbA1c | Overall | rs11708067 | G | A | 0.245 | -0.353 | 0.018 | 1.6E-85 | 384.3 |
| HbA1c | Overall | rs12497133 | A | G | 0.533 | 0.120 | 0.016 | 2.1E-14 | 58.4 |
| HbA1c | Overall | rs13059110 | T | G | 0.133 | 0.144 | 0.023 | 3.6E-10 | 39.3 |
| HbA1c | Overall | rs75367758 | A | G | 0.048 | -0.296 | 0.036 | 3.2E-16 | 66.7 |
| HbA1c | Overall | rs12493136 | A | G | 0.164 | 0.169 | 0.021 | 1.0E-15 | 64.4 |
| HbA1c | Overall | rs11926854 | C | T | 0.037 | -0.240 | 0.042 | 7.9E-09 | 33.3 |
| HbA1c | Overall | rs6785881 | T | C | 0.481 | -0.140 | 0.016 | 2.2E-19 | 81.1 |
| HbA1c | Overall | rs4955744 | C | A | 0.146 | 0.134 | 0.022 | 1.0E-09 | 37.3 |
| HbA1c | Overall | rs1905505 | A | G | 0.284 | -0.283 | 0.017 | 1.2E-60 | 270.0 |
| HbA1c | Overall | rs360408 | A | T | 0.565 | -0.124 | 0.016 | 9.1E-15 | 60.1 |
| HbA1c | Overall | rs4894797 | A | G | 0.576 | 0.188 | 0.016 | 4.5E-33 | 143.6 |
| HbA1c | Overall | rs4859137 | T | C | 0.772 | 0.117 | 0.019 | 3.4E-10 | 39.4 |
| HbA1c | Overall | rs73061095 | G | T | 0.314 | 0.147 | 0.017 | 1.3E-18 | 77.6 |
| HbA1c | Overall | rs6777684 | G | A | 0.610 | 0.175 | 0.016 | 7.2E-28 | 119.8 |
| HbA1c | Overall | rs41297495 | T | C | 0.382 | -0.099 | 0.016 | 4.9E-10 | 38.7 |
| HbA1c | Overall | rs2015708 | T | C | 0.137 | 0.167 | 0.023 | 1.5E-13 | 54.5 |
| HbA1c | Overall | rs2686588 | G | C | 0.565 | -0.135 | 0.016 | 6.0E-18 | 74.5 |
| HbA1c | Overall | rs1496653 | G | A | 0.204 | -0.138 | 0.019 | 6.8E-13 | 51.6 |

| | | | | | | | | | |
|---|---|---|---|---|---|---|---|---|---|
| HbA1c | Overall | rs4678917 | C | T | 0.105 | -0.146 | 0.026 | 1.3E-08 | 32.4 |
| HbA1c | Overall | rs62246446 | G | C | 0.176 | -0.138 | 0.020 | 1.3E-11 | 45.9 |
| HbA1c | Overall | rs2230929 | A | G | 0.171 | 0.258 | 0.021 | 8.8E-36 | 156.0 |
| HbA1c | Overall | rs2336664 | G | A | 0.363 | 0.097 | 0.016 | 2.0E-09 | 36.0 |
| HbA1c | Overall | rs11130982 | G | T | 0.711 | 0.110 | 0.017 | 1.6E-10 | 40.9 |
| HbA1c | Overall | rs9840088 | C | A | 0.201 | -0.113 | 0.020 | 7.9E-09 | 33.3 |
| HbA1c | Overall | rs397834892 | G | A | 0.471 | -0.105 | 0.016 | 1.6E-11 | 45.4 |
| HbA1c | Overall | rs17508261 | C | T | 0.122 | 0.188 | 0.024 | 2.5E-15 | 62.6 |
| HbA1c | Overall | rs9994509 | G | A | 0.393 | -0.135 | 0.016 | 2.8E-17 | 71.5 |
| HbA1c | Overall | rs2044341 | C | T | 0.684 | -0.356 | 0.017 | 5.9E-100 | 450.7 |
| HbA1c | Overall | rs76505278 | C | T | 0.103 | -0.187 | 0.026 | 2.7E-13 | 53.5 |
| HbA1c | Overall | rs13125518 | G | A | 0.459 | -0.090 | 0.016 | 1.1E-08 | 32.7 |
| HbA1c | Overall | rs75636043 | A | G | 0.078 | -0.172 | 0.029 | 3.9E-09 | 34.7 |
| HbA1c | Overall | rs6840504 | T | C | 0.443 | 0.113 | 0.016 | 4.9E-13 | 52.3 |
| HbA1c | Overall | rs55881843 | A | G | 0.139 | -0.226 | 0.023 | 1.5E-23 | 100.0 |
| HbA1c | Overall | rs34811474 | A | G | 0.232 | -0.107 | 0.018 | 5.0E-09 | 34.2 |
| HbA1c | Overall | rs6844176 | C | T | 0.455 | 0.087 | 0.016 | 2.8E-08 | 30.9 |
| HbA1c | Overall | rs9990955 | T | C | 0.284 | 0.126 | 0.017 | 2.1E-13 | 53.9 |
| HbA1c | Overall | rs4689394 | G | C | 0.599 | 0.121 | 0.016 | 2.9E-14 | 57.8 |
| HbA1c | Overall | rs1481012 | G | A | 0.113 | 0.198 | 0.025 | 6.9E-16 | 65.2 |
| HbA1c | Overall | rs35188965 | T | C | 0.582 | 0.120 | 0.016 | 2.6E-14 | 58.0 |
| HbA1c | Overall | rs469753 | T | G | 0.475 | 0.091 | 0.016 | 5.3E-09 | 34.1 |
| HbA1c | Overall | rs10053046 | G | A | 0.329 | 0.118 | 0.017 | 1.5E-12 | 50.1 |
| HbA1c | Overall | rs6885132 | G | C | 0.099 | -0.233 | 0.026 | 8.5E-19 | 78.4 |
| HbA1c | Overall | rs35018839 | G | A | 0.095 | -0.151 | 0.027 | 1.9E-08 | 31.6 |
| HbA1c | Overall | rs13167071 | C | G | 0.640 | 0.135 | 0.016 | 5.9E-17 | 70.0 |
| HbA1c | Overall | rs7720275 | C | T | 0.170 | 0.161 | 0.021 | 6.2E-15 | 60.8 |
| HbA1c | Overall | rs56395926 | A | G | 0.229 | -0.108 | 0.019 | 5.2E-09 | 34.1 |
| HbA1c | Overall | rs3857286 | T | C | 0.695 | -0.104 | 0.017 | 9.4E-10 | 37.5 |
| HbA1c | Overall | rs6893283 | T | C | 0.015 | -0.377 | 0.065 | 6.0E-09 | 33.8 |
| HbA1c | Overall | rs145381182 | A | G | 0.090 | 0.157 | 0.027 | 1.0E-08 | 32.8 |
| HbA1c | Overall | rs2270927 | G | C | 0.094 | 0.193 | 0.027 | 4.1E-13 | 52.6 |
| HbA1c | Overall | rs6878122 | A | G | 0.680 | -0.181 | 0.017 | 1.5E-27 | 118.3 |
| HbA1c | Overall | rs17419291 | C | T | 0.087 | -0.165 | 0.028 | 1.9E-09 | 36.1 |
| HbA1c | Overall | rs2546197 | G | T | 0.078 | 0.160 | 0.029 | 3.6E-08 | 30.3 |
| HbA1c | Overall | rs144489757 | G | C | 0.284 | -0.120 | 0.017 | 3.0E-12 | 48.7 |
| HbA1c | Overall | rs148570357 | T | C | 0.295 | -0.155 | 0.017 | 1.6E-19 | 81.7 |
| HbA1c | Overall | rs72939920 | T | A | 0.235 | -0.194 | 0.018 | 4.6E-26 | 111.5 |
| HbA1c | Overall | rs9398642 | T | C | 0.145 | -0.132 | 0.022 | 2.2E-09 | 35.8 |
| HbA1c | Overall | rs1490384 | T | C | 0.498 | 0.122 | 0.016 | 4.8E-15 | 61.3 |
| HbA1c | Overall | rs9402645 | T | G | 0.200 | 0.119 | 0.020 | 1.1E-09 | 37.1 |
| HbA1c | Overall | rs9376091 | T | C | 0.262 | -0.288 | 0.018 | 4.3E-59 | 262.9 |
| HbA1c | Overall | rs592423 | C | A | 0.554 | -0.131 | 0.016 | 3.6E-17 | 71.0 |
| HbA1c | Overall | rs3910736 | T | C | 0.308 | 0.110 | 0.017 | 7.1E-11 | 42.5 |
| HbA1c | Overall | rs262813 | C | T | 0.478 | 0.090 | 0.016 | 8.0E-09 | 33.3 |
| HbA1c | Overall | rs2665357 | C | A | 0.517 | 0.120 | 0.016 | 1.5E-14 | 59.2 |
| HbA1c | Overall | rs67131976 | T | C | 0.172 | 0.315 | 0.021 | 3.2E-53 | 235.9 |
| HbA1c | Overall | rs142513102 | G | A | 0.026 | -0.484 | 0.049 | 1.9E-23 | 99.6 |

| | | | | | | | | | |
|---|---|---|---|---|---|---|---|---|---|
| HbA1c | Overall | rs1800562 | A | G | 0.079 | -0.842 | 0.029 | 6.8E-188 | 855.8 |
| HbA1c | Overall | rs2442725 | C | T | 0.694 | 0.159 | 0.017 | 4.3E-21 | 88.8 |
| HbA1c | Overall | rs9268469 | A | G | 0.155 | 0.301 | 0.021 | 1.2E-44 | 196.5 |
| HbA1c | Overall | rs833058 | T | C | 0.365 | 0.111 | 0.016 | 5.2E-12 | 47.6 |
| HbA1c | Overall | rs998584 | A | C | 0.482 | 0.156 | 0.016 | 1.2E-23 | 100.6 |
| HbA1c | Overall | rs2294856 | C | G | 0.355 | 0.114 | 0.016 | 1.9E-12 | 49.6 |
| HbA1c | Overall | rs3748059 | G | A | 0.799 | -0.149 | 0.019 | 1.6E-14 | 58.9 |
| HbA1c | Overall | rs13192435 | C | T | 0.025 | 0.353 | 0.049 | 8.5E-13 | 51.2 |
| HbA1c | Overall | rs78588343 | A | G | 0.179 | -0.226 | 0.020 | 8.3E-29 | 124.0 |
| HbA1c | Overall | rs1171105 | C | T | 0.695 | 0.095 | 0.017 | 4.1E-08 | 30.1 |
| HbA1c | Overall | rs12667932 | A | G | 0.080 | -0.314 | 0.029 | 5.5E-28 | 120.3 |
| HbA1c | Overall | rs35722851 | C | T | 0.495 | -0.088 | 0.016 | 1.5E-08 | 32.1 |
| HbA1c | Overall | rs8176059 | A | G | 0.011 | -0.439 | 0.073 | 2.2E-09 | 35.8 |
| HbA1c | Overall | rs17168486 | T | C | 0.173 | 0.198 | 0.021 | 7.4E-22 | 92.3 |
| HbA1c | Overall | rs10487796 | A | T | 0.450 | -0.183 | 0.016 | 9.0E-32 | 137.6 |
| HbA1c | Overall | rs6459737 | A | G | 0.347 | -0.117 | 0.016 | 6.4E-13 | 51.7 |
| HbA1c | Overall | rs60238952 | G | A | 0.219 | -0.113 | 0.019 | 2.3E-09 | 35.7 |
| HbA1c | Overall | rs1708302 | T | C | 0.501 | -0.178 | 0.016 | 1.4E-30 | 132.2 |
| HbA1c | Overall | rs76323047 | G | A | 0.118 | 0.335 | 0.024 | 5.4E-44 | 193.6 |
| HbA1c | Overall | rs1004558 | T | C | 0.179 | 0.727 | 0.020 | 4.1E-283 | 1295.3 |
| HbA1c | Overall | rs836472 | A | G | 0.266 | -0.109 | 0.018 | 4.7E-10 | 38.8 |
| HbA1c | Overall | rs2460421 | T | A | 0.502 | -0.112 | 0.015 | 5.8E-13 | 51.9 |
| HbA1c | Overall | rs6953344 | G | A | 0.246 | 0.127 | 0.018 | 2.2E-12 | 49.3 |
| HbA1c | Overall | rs3211821 | G | A | 0.562 | -0.106 | 0.016 | 1.7E-11 | 45.3 |
| HbA1c | Overall | rs6973494 | A | C | 0.516 | 0.116 | 0.016 | 1.4E-13 | 54.7 |
| HbA1c | Overall | rs10488532 | T | C | 0.108 | -0.143 | 0.025 | 1.1E-08 | 32.6 |
| HbA1c | Overall | rs117370443 | C | T | 0.039 | 0.373 | 0.040 | 1.4E-20 | 86.6 |
| HbA1c | Overall | rs636089 | C | T | 0.386 | 0.095 | 0.016 | 2.9E-09 | 35.3 |
| HbA1c | Overall | rs7820334 | T | C | 0.311 | -0.146 | 0.017 | 4.8E-18 | 75.0 |
| HbA1c | Overall | rs2737263 | T | G | 0.284 | -0.156 | 0.017 | 1.1E-19 | 82.4 |
| HbA1c | Overall | rs13266634 | T | C | 0.311 | -0.379 | 0.017 | 2.4E-113 | 512.3 |
| HbA1c | Overall | rs58253018 | T | A | 0.635 | 0.105 | 0.017 | 2.3E-10 | 40.2 |
| HbA1c | Overall | rs2060985 | G | A | 0.537 | -0.122 | 0.016 | 4.8E-15 | 61.3 |
| HbA1c | Overall | rs3757971 | C | T | 0.380 | 0.130 | 0.016 | 3.2E-16 | 66.7 |
| HbA1c | Overall | rs187055391 | A | G | 0.172 | -0.134 | 0.021 | 1.5E-10 | 41.0 |
| HbA1c | Overall | rs66593272 | T | A | 0.038 | -0.915 | 0.041 | 2.4E-111 | 503.0 |
| HbA1c | Overall | rs4737010 | A | G | 0.228 | 0.534 | 0.019 | 1.0E-182 | 831.9 |
| HbA1c | Overall | rs55840085 | A | G | 0.355 | 0.169 | 0.016 | 1.7E-25 | 108.9 |
| HbA1c | Overall | rs28708563 | A | G | 0.042 | 0.220 | 0.039 | 2.3E-08 | 31.3 |
| HbA1c | Overall | rs62523081 | A | G | 0.362 | 0.094 | 0.016 | 7.1E-09 | 33.5 |
| HbA1c | Overall | rs1967604 | G | A | 0.712 | -0.208 | 0.017 | 1.1E-33 | 146.3 |
| HbA1c | Overall | rs1560980 | C | G | 0.041 | 0.316 | 0.039 | 5.1E-16 | 65.8 |
| HbA1c | Overall | rs584048 | T | C | 0.861 | 0.134 | 0.022 | 2.1E-09 | 35.9 |
| HbA1c | Overall | rs5900831 | G | T | 0.653 | 0.135 | 0.017 | 3.2E-16 | 66.7 |
| HbA1c | Overall | rs550057 | T | C | 0.255 | 0.325 | 0.018 | 5.9E-74 | 331.1 |
| HbA1c | Overall | rs28641468 | C | T | 0.756 | 0.234 | 0.018 | 1.9E-38 | 168.2 |
| HbA1c | Overall | rs10811660 | A | G | 0.173 | -0.359 | 0.020 | 1.5E-68 | 306.3 |
| HbA1c | Overall | rs10758593 | A | G | 0.397 | 0.107 | 0.016 | 1.1E-11 | 46.1 |

| | | | | | | | | | |
|---|---|---|---|---|---|---|---|---|---|
| HbA1c | Overall | rs2236496 | C | T | 0.208 | -0.182 | 0.019 | 2.5E-21 | 89.9 |
| HbA1c | Overall | rs11145347 | C | T | 0.197 | 0.239 | 0.020 | 1.7E-34 | 150.1 |
| HbA1c | Overall | rs61750929 | T | C | 0.055 | -0.692 | 0.034 | 8.6E-92 | 413.1 |
| HbA1c | Overall | rs7047279 | C | T | 0.568 | -0.124 | 0.016 | 4.2E-15 | 61.6 |
| HbA1c | Women | rs7903146 | T | C | 0.290 | 0.423 | 0.023 | 5.3E-76 | 340.7 |
| HbA1c | Women | rs11257655 | T | C | 0.208 | 0.234 | 0.026 | 7.3E-20 | 83.2 |
| HbA1c | Women | rs7923442 | G | A | 0.178 | -0.156 | 0.027 | 9.8E-09 | 32.9 |
| HbA1c | Women | rs61850681 | A | G | 0.239 | -0.218 | 0.025 | 5.9E-19 | 79.1 |
| HbA1c | Women | rs10998678 | A | G | 0.053 | 0.341 | 0.047 | 2.6E-13 | 53.5 |
| HbA1c | Women | rs2015803 | T | C | 0.247 | 0.318 | 0.024 | 1.4E-39 | 173.3 |
| HbA1c | Women | rs17476364 | C | T | 0.110 | -2.079 | 0.033 | 0.0E+00 | 3990.7 |
| HbA1c | Women | rs2305196 | A | G | 0.265 | -0.221 | 0.024 | 1.1E-20 | 86.9 |
| HbA1c | Women | rs7077479 | A | C | 0.337 | 0.140 | 0.022 | 3.9E-10 | 39.2 |
| HbA1c | Women | rs701848 | C | T | 0.390 | -0.118 | 0.021 | 3.2E-08 | 30.6 |
| HbA1c | Women | rs11187149 | A | G | 0.381 | -0.189 | 0.021 | 9.1E-19 | 78.3 |
| HbA1c | Women | rs4418728 | T | G | 0.449 | -0.152 | 0.021 | 3.5E-13 | 52.9 |
| HbA1c | Women | rs12365580 | A | G | 0.151 | -0.209 | 0.029 | 7.6E-13 | 51.4 |
| HbA1c | Women | rs11042850 | A | G | 0.087 | 0.292 | 0.037 | 2.0E-15 | 63.1 |
| HbA1c | Women | rs7944748 | G | T | 0.378 | 0.119 | 0.022 | 3.2E-08 | 30.6 |
| HbA1c | Women | rs2351960 | G | A | 0.893 | -0.219 | 0.034 | 7.8E-11 | 42.3 |
| HbA1c | Women | rs3782123 | A | C | 0.733 | -0.203 | 0.024 | 6.9E-18 | 74.3 |
| HbA1c | Women | rs11564725 | T | C | 0.236 | 0.202 | 0.025 | 3.1E-16 | 66.7 |
| HbA1c | Women | rs4755950 | T | C | 0.113 | -0.187 | 0.033 | 1.2E-08 | 32.5 |
| HbA1c | Women | rs10501320 | C | G | 0.264 | -0.194 | 0.024 | 1.8E-16 | 67.8 |
| HbA1c | Women | rs174598 | A | G | 0.329 | -0.148 | 0.022 | 2.4E-11 | 44.6 |
| HbA1c | Women | rs12294913 | G | C | 0.050 | 0.299 | 0.050 | 1.6E-09 | 36.4 |
| HbA1c | Women | rs11602873 | T | A | 0.157 | -0.224 | 0.029 | 4.3E-15 | 61.6 |
| HbA1c | Women | rs10830963 | G | C | 0.275 | 0.434 | 0.023 | 1.7E-77 | 347.5 |
| HbA1c | Women | rs1392026 | T | C | 0.635 | -0.175 | 0.022 | 7.3E-16 | 65.1 |
| HbA1c | Women | rs7310615 | G | C | 0.517 | 0.188 | 0.021 | 2.9E-19 | 80.5 |
| HbA1c | Women | rs2936839 | C | T | 0.610 | -0.129 | 0.021 | 1.6E-09 | 36.4 |
| HbA1c | Women | rs147730268 | T | G | 0.091 | -0.229 | 0.037 | 9.2E-10 | 37.5 |
| HbA1c | Women | rs10842994 | T | C | 0.196 | -0.156 | 0.026 | 2.4E-09 | 35.7 |
| HbA1c | Women | rs76895963 | G | T | 0.020 | -1.082 | 0.080 | 1.1E-41 | 183.1 |
| HbA1c | Women | rs10492081 | G | A | 0.180 | 0.198 | 0.027 | 2.4E-13 | 53.7 |
| HbA1c | Women | rs4760682 | A | C | 0.813 | 0.501 | 0.027 | 5.8E-79 | 354.3 |
| HbA1c | Women | rs35979828 | T | C | 0.069 | -0.236 | 0.041 | 8.9E-09 | 33.1 |
| HbA1c | Women | rs2261181 | T | C | 0.096 | 0.244 | 0.035 | 5.8E-12 | 47.4 |
| HbA1c | Women | rs1579238 | G | A | 0.241 | -0.157 | 0.024 | 1.1E-10 | 41.6 |
| HbA1c | Women | rs34011672 | G | A | 0.089 | 0.562 | 0.037 | 4.0E-53 | 235.5 |
| HbA1c | Women | rs12861645 | G | T | 0.272 | 0.241 | 0.023 | 8.1E-25 | 105.8 |
| HbA1c | Women | rs11616945 | C | T | 0.337 | 0.140 | 0.022 | 4.0E-10 | 39.1 |
| HbA1c | Women | rs12020979 | G | T | 0.185 | -0.268 | 0.028 | 6.3E-22 | 92.7 |
| HbA1c | Women | rs4325403 | A | T | 0.220 | 0.231 | 0.025 | 1.0E-19 | 82.6 |
| HbA1c | Women | rs75629141 | A | T | 0.829 | -0.164 | 0.028 | 3.0E-09 | 35.2 |
| HbA1c | Women | rs941899 | C | T | 0.529 | -0.135 | 0.021 | 8.7E-11 | 42.1 |
| HbA1c | Women | rs9324022 | C | T | 0.206 | 0.163 | 0.026 | 5.6E-10 | 38.5 |
| HbA1c | Women | rs1760940 | C | A | 0.248 | 0.176 | 0.024 | 2.4E-13 | 53.6 |

| | | | | | | | | | |
|---|---|---|---|---|---|---|---|---|---|
| HbA1c | Women | rs941718 | C | T | 0.278 | 0.151 | 0.023 | 7.3E-11 | 42.4 |
| HbA1c | Women | rs8018574 | A | T | 0.243 | -0.154 | 0.024 | 2.4E-10 | 40.1 |
| HbA1c | Women | rs12878001 | G | T | 0.160 | 0.160 | 0.029 | 2.1E-08 | 31.4 |
| HbA1c | Women | rs229572 | A | T | 0.235 | 0.190 | 0.025 | 8.5E-15 | 60.2 |
| HbA1c | Women | rs35889227 | T | G | 0.631 | -0.143 | 0.022 | 3.3E-11 | 44.0 |
| HbA1c | Women | rs4516170 | C | T | 0.050 | -0.313 | 0.048 | 5.8E-11 | 42.9 |
| HbA1c | Women | rs1550026 | T | A | 0.408 | 0.137 | 0.021 | 1.1E-10 | 41.6 |
| HbA1c | Women | rs869301 | C | T | 0.720 | 0.192 | 0.023 | 1.2E-16 | 68.6 |
| HbA1c | Women | rs11638511 | G | A | 0.561 | 0.126 | 0.021 | 2.2E-09 | 35.8 |
| HbA1c | Women | rs4451969 | C | T | 0.671 | -0.150 | 0.022 | 1.5E-11 | 45.6 |
| HbA1c | Women | rs9302377 | T | C | 0.493 | 0.123 | 0.022 | 2.5E-08 | 31.1 |
| HbA1c | Women | rs6600191 | C | T | 0.181 | -0.218 | 0.027 | 9.5E-16 | 64.5 |
| HbA1c | Women | rs35661204 | T | C | 0.226 | 0.189 | 0.025 | 3.2E-14 | 57.6 |
| HbA1c | Women | rs8050500 | C | T | 0.446 | -0.162 | 0.021 | 1.1E-14 | 59.8 |
| HbA1c | Women | rs11075985 | A | C | 0.423 | 0.124 | 0.021 | 3.8E-09 | 34.7 |
| HbA1c | Women | rs9929239 | T | C | 0.292 | 0.159 | 0.023 | 3.0E-12 | 48.7 |
| HbA1c | Women | rs247828 | C | A | 0.305 | 0.138 | 0.023 | 9.8E-10 | 37.4 |
| HbA1c | Women | rs72811487 | A | G | 0.035 | -0.413 | 0.056 | 2.0E-13 | 54.1 |
| HbA1c | Women | rs551118 | G | C | 0.578 | 0.372 | 0.021 | 1.1E-67 | 302.4 |
| HbA1c | Women | rs11655029 | C | T | 0.695 | -0.211 | 0.023 | 1.1E-20 | 87.0 |
| HbA1c | Women | rs6902 | C | A | 0.386 | -0.118 | 0.021 | 3.4E-08 | 30.5 |
| HbA1c | Women | rs12450826 | T | A | 0.293 | -0.172 | 0.023 | 2.4E-13 | 53.7 |
| HbA1c | Women | rs11651755 | T | C | 0.515 | -0.133 | 0.021 | 1.7E-10 | 40.8 |
| HbA1c | Women | rs2060779 | A | G | 0.222 | -0.176 | 0.025 | 2.3E-12 | 49.3 |
| HbA1c | Women | rs9899611 | G | A | 0.213 | 0.163 | 0.025 | 1.3E-10 | 41.3 |
| HbA1c | Women | rs858518 | A | G | 0.573 | -0.144 | 0.021 | 7.6E-12 | 46.9 |
| HbA1c | Women | rs2748427 | G | A | 0.218 | 0.637 | 0.025 | 5.2E-142 | 644.8 |
| HbA1c | Women | rs112281307 | C | A | 0.094 | 0.229 | 0.036 | 1.2E-10 | 41.5 |
| HbA1c | Women | rs9909940 | T | C | 0.312 | 0.474 | 0.022 | 3.2E-99 | 447.6 |
| HbA1c | Women | rs4792590 | G | A | 0.877 | -0.215 | 0.032 | 1.2E-11 | 45.9 |
| HbA1c | Women | rs4542747 | T | C | 0.746 | 0.153 | 0.024 | 1.7E-10 | 40.8 |
| HbA1c | Women | rs8110787 | T | C | 0.391 | -0.212 | 0.021 | 2.0E-23 | 99.4 |
| HbA1c | Women | rs4808579 | C | T | 0.392 | 0.300 | 0.021 | 7.7E-45 | 197.5 |
| HbA1c | Women | rs4499344 | A | G | 0.293 | -0.211 | 0.023 | 2.7E-20 | 85.2 |
| HbA1c | Women | rs10410896 | A | C | 0.890 | 0.199 | 0.034 | 3.7E-09 | 34.8 |
| HbA1c | Women | rs55914970 | T | G | 0.441 | -0.154 | 0.021 | 2.2E-13 | 53.8 |
| HbA1c | Women | rs12459419 | T | C | 0.326 | -0.163 | 0.022 | 2.0E-13 | 54.1 |
| HbA1c | Women | rs7250849 | T | G | 0.106 | 0.235 | 0.034 | 3.8E-12 | 48.2 |
| HbA1c | Women | rs351988 | G | A | 0.565 | -0.117 | 0.021 | 3.3E-08 | 30.5 |
| HbA1c | Women | rs1975283 | C | A | 0.685 | -0.169 | 0.023 | 6.6E-14 | 56.2 |
| HbA1c | Women | rs267738 | G | T | 0.221 | -0.201 | 0.025 | 9.0E-16 | 64.7 |
| HbA1c | Women | rs4133213 | A | C | 0.450 | -0.121 | 0.021 | 1.5E-08 | 32.1 |
| HbA1c | Women | rs857721 | A | T | 0.267 | 0.504 | 0.023 | 1.8E-102 | 462.6 |
| HbA1c | Women | rs7528296 | C | T | 0.386 | -0.130 | 0.021 | 1.1E-09 | 37.1 |
| HbA1c | Women | rs1044145 | C | T | 0.540 | 0.155 | 0.021 | 1.3E-13 | 54.8 |
| HbA1c | Women | rs340882 | G | C | 0.621 | 0.254 | 0.022 | 4.8E-32 | 138.9 |
| HbA1c | Women | rs348330 | A | G | 0.635 | -0.131 | 0.022 | 1.5E-09 | 36.5 |
| HbA1c | Women | rs1339847 | A | G | 0.110 | -0.224 | 0.033 | 1.7E-11 | 45.3 |

| | | | | | | | | | |
|---|---|---|---|---|---|---|---|---|---|
| HbA1c | Women | rs2301153 | C | G | 0.799 | -0.171 | 0.028 | 4.9E-10 | 38.7 |
| HbA1c | Women | rs563437995 | C | G | 0.008 | -1.019 | 0.123 | 1.1E-16 | 68.8 |
| HbA1c | Women | rs1175550 | G | A | 0.230 | -0.268 | 0.025 | 1.1E-26 | 114.4 |
| HbA1c | Women | rs12141885 | T | C | 0.700 | -0.126 | 0.023 | 2.6E-08 | 31.0 |
| HbA1c | Women | rs1768809 | C | A | 0.556 | 0.132 | 0.021 | 3.0E-10 | 39.7 |
| HbA1c | Women | rs12121576 | G | A | 0.183 | -0.163 | 0.028 | 4.9E-09 | 34.2 |
| HbA1c | Women | rs149142833 | T | C | 0.156 | 0.165 | 0.029 | 1.2E-08 | 32.5 |
| HbA1c | Women | rs6066148 | C | G | 0.262 | -0.144 | 0.024 | 1.3E-09 | 36.8 |
| HbA1c | Women | rs6014993 | G | A | 0.487 | 0.204 | 0.021 | 1.5E-22 | 95.6 |
| HbA1c | Women | rs73129526 | C | T | 0.144 | 0.168 | 0.030 | 1.4E-08 | 32.2 |
| HbA1c | Women | rs6077396 | A | G | 0.507 | -0.114 | 0.021 | 4.4E-08 | 30.0 |
| HbA1c | Women | rs3827181 | T | C | 0.346 | 0.131 | 0.022 | 2.3E-09 | 35.7 |
| HbA1c | Women | rs61138219 | T | C | 0.153 | -0.160 | 0.029 | 3.3E-08 | 30.5 |
| HbA1c | Women | rs17850433 | C | T | 0.012 | 0.624 | 0.095 | 4.7E-11 | 43.3 |
| HbA1c | Women | rs62221525 | T | C | 0.179 | -0.168 | 0.027 | 7.1E-10 | 38.0 |
| HbA1c | Women | rs4680 | A | G | 0.516 | 0.119 | 0.021 | 9.4E-09 | 33.0 |
| HbA1c | Women | rs6518681 | G | A | 0.907 | 0.206 | 0.036 | 1.0E-08 | 32.8 |
| HbA1c | Women | rs855791 | G | A | 0.561 | -0.417 | 0.021 | 1.3E-87 | 394.1 |
| HbA1c | Women | rs2143918 | C | A | 0.471 | -0.166 | 0.021 | 1.9E-15 | 63.1 |
| HbA1c | Women | rs111631066 | A | G | 0.015 | -0.550 | 0.087 | 2.8E-10 | 39.8 |
| HbA1c | Women | rs180935712 | A | G | 0.019 | 0.533 | 0.078 | 9.8E-12 | 46.4 |
| HbA1c | Women | rs143869345 | G | A | 0.017 | 0.681 | 0.082 | 1.1E-16 | 68.9 |
| HbA1c | Women | rs560887 | C | T | 0.701 | 0.738 | 0.023 | 1.2E-232 | 1063.7 |
| HbA1c | Women | rs4972439 | C | T | 0.220 | -0.192 | 0.025 | 2.6E-14 | 58.0 |
| HbA1c | Women | rs1047891 | A | C | 0.316 | -0.154 | 0.022 | 5.6E-12 | 47.5 |
| HbA1c | Women | rs13427681 | C | G | 0.562 | 0.129 | 0.021 | 7.0E-10 | 38.0 |
| HbA1c | Women | rs2943640 | C | A | 0.652 | 0.121 | 0.022 | 2.7E-08 | 30.9 |
| HbA1c | Women | rs7346 | A | G | 0.191 | -0.163 | 0.027 | 7.7E-10 | 37.8 |
| HbA1c | Women | rs72780125 | C | T | 0.131 | 0.429 | 0.031 | 8.3E-44 | 192.8 |
| HbA1c | Women | rs7595986 | A | G | 0.781 | -0.156 | 0.025 | 5.1E-10 | 38.7 |
| HbA1c | Women | rs1504 | G | T | 0.444 | -0.116 | 0.021 | 2.9E-08 | 30.8 |
| HbA1c | Women | rs112694524 | A | G | 0.072 | -0.469 | 0.041 | 2.0E-30 | 131.4 |
| HbA1c | Women | rs2121564 | G | C | 0.129 | 0.169 | 0.031 | 4.8E-08 | 29.8 |
| HbA1c | Women | rs78583604 | T | G | 0.132 | 0.175 | 0.032 | 3.1E-08 | 30.6 |
| HbA1c | Women | rs13019832 | A | G | 0.418 | -0.149 | 0.021 | 2.2E-12 | 49.3 |
| HbA1c | Women | rs7572278 | A | T | 0.204 | 0.175 | 0.026 | 1.2E-11 | 45.9 |
| HbA1c | Women | rs9826367 | G | A | 0.449 | -0.258 | 0.021 | 5.2E-35 | 152.4 |
| HbA1c | Women | rs11708067 | G | A | 0.245 | -0.392 | 0.024 | 2.8E-59 | 263.8 |
| HbA1c | Women | rs6807945 | T | C | 0.837 | -0.186 | 0.028 | 5.7E-11 | 42.9 |
| HbA1c | Women | rs77630894 | T | G | 0.049 | -0.266 | 0.048 | 3.8E-08 | 30.3 |
| HbA1c | Women | rs12493136 | A | G | 0.164 | 0.200 | 0.028 | 1.3E-12 | 50.3 |
| HbA1c | Women | rs16831139 | G | A | 0.400 | -0.137 | 0.022 | 2.5E-10 | 40.0 |
| HbA1c | Women | rs5398 | A | G | 0.285 | -0.284 | 0.023 | 4.6E-35 | 152.7 |
| HbA1c | Women | rs360408 | A | T | 0.565 | -0.123 | 0.021 | 9.8E-09 | 32.9 |
| HbA1c | Women | rs16856859 | G | A | 0.583 | 0.199 | 0.021 | 5.0E-21 | 88.5 |
| HbA1c | Women | rs10222374 | T | C | 0.785 | 0.152 | 0.026 | 2.6E-09 | 35.4 |
| HbA1c | Women | rs6777684 | G | A | 0.610 | 0.173 | 0.021 | 5.4E-16 | 65.6 |
| HbA1c | Women | rs2686588 | G | C | 0.565 | -0.133 | 0.021 | 2.2E-10 | 40.3 |

| | | | | | | | | | |
|---|---|---|---|---|---|---|---|---|---|
| HbA1c | Women | rs62246446 | G | C | 0.176 | -0.166 | 0.027 | 1.1E-09 | 37.1 |
| HbA1c | Women | rs67882627 | T | A | 0.171 | 0.251 | 0.028 | 1.1E-19 | 82.4 |
| HbA1c | Women | rs11130982 | G | T | 0.711 | 0.178 | 0.023 | 1.3E-14 | 59.5 |
| HbA1c | Women | rs223376 | A | T | 0.485 | -0.122 | 0.021 | 5.6E-09 | 34.0 |
| HbA1c | Women | rs17430251 | G | A | 0.122 | 0.178 | 0.032 | 2.1E-08 | 31.4 |
| HbA1c | Women | rs2298978 | A | G | 0.403 | -0.142 | 0.021 | 2.5E-11 | 44.6 |
| HbA1c | Women | rs6814613 | C | G | 0.665 | -0.364 | 0.023 | 1.1E-58 | 261.0 |
| HbA1c | Women | rs34835465 | C | T | 0.070 | -0.295 | 0.041 | 5.8E-13 | 51.9 |
| HbA1c | Women | rs6840504 | T | C | 0.443 | 0.127 | 0.021 | 1.5E-09 | 36.5 |
| HbA1c | Women | rs11132255 | A | G | 0.161 | -0.237 | 0.028 | 5.9E-17 | 70.0 |
| HbA1c | Women | rs11731874 | C | T | 0.575 | 0.123 | 0.021 | 6.5E-09 | 33.7 |
| HbA1c | Women | rs2622622 | G | C | 0.314 | -0.158 | 0.023 | 2.9E-12 | 48.8 |
| HbA1c | Women | rs112034864 | G | C | 0.503 | 0.141 | 0.021 | 1.9E-11 | 45.1 |
| HbA1c | Women | rs6885132 | G | C | 0.099 | -0.252 | 0.035 | 9.5E-13 | 51.0 |
| HbA1c | Women | rs4704834 | G | A | 0.646 | 0.181 | 0.022 | 1.1E-16 | 68.9 |
| HbA1c | Women | rs4496693 | C | T | 0.094 | 0.219 | 0.036 | 9.1E-10 | 37.5 |
| HbA1c | Women | rs17419291 | C | T | 0.087 | -0.211 | 0.037 | 1.0E-08 | 32.8 |
| HbA1c | Women | rs35247507 | G | A | 0.286 | -0.164 | 0.023 | 1.3E-12 | 50.4 |
| HbA1c | Women | rs13214717 | A | C | 0.299 | -0.165 | 0.023 | 3.7E-13 | 52.8 |
| HbA1c | Women | rs72939920 | T | A | 0.235 | -0.205 | 0.025 | 7.1E-17 | 69.7 |
| HbA1c | Women | rs9385400 | G | T | 0.455 | 0.160 | 0.021 | 1.9E-14 | 58.6 |
| HbA1c | Women | rs9376091 | T | C | 0.262 | -0.274 | 0.024 | 1.0E-30 | 132.8 |
| HbA1c | Women | rs592423 | C | A | 0.554 | -0.114 | 0.021 | 4.6E-08 | 29.9 |
| HbA1c | Women | rs6920313 | C | T | 0.306 | 0.131 | 0.023 | 6.4E-09 | 33.7 |
| HbA1c | Women | rs9368219 | T | C | 0.173 | 0.282 | 0.028 | 1.6E-24 | 104.6 |
| HbA1c | Women | rs1800562 | A | G | 0.079 | -0.871 | 0.039 | 7.0E-113 | 510.5 |
| HbA1c | Women | rs2071596 | A | G | 0.163 | -0.232 | 0.028 | 1.4E-16 | 68.2 |
| HbA1c | Women | rs11967891 | T | A | 0.181 | 0.320 | 0.028 | 3.7E-30 | 130.3 |
| HbA1c | Women | rs9272606 | T | C | 0.660 | 0.127 | 0.022 | 8.6E-09 | 33.1 |
| HbA1c | Women | rs998584 | A | C | 0.482 | 0.191 | 0.021 | 4.9E-20 | 84.0 |
| HbA1c | Women | rs34101263 | A | G | 0.299 | 0.148 | 0.023 | 6.6E-11 | 42.6 |
| HbA1c | Women | rs3748059 | G | A | 0.799 | -0.168 | 0.026 | 9.6E-11 | 41.9 |
| HbA1c | Women | rs78588343 | A | G | 0.179 | -0.268 | 0.027 | 5.1E-23 | 97.6 |
| HbA1c | Women | rs75759008 | G | A | 0.080 | -0.284 | 0.038 | 1.2E-13 | 55.1 |
| HbA1c | Women | rs12154627 | C | T | 0.498 | -0.120 | 0.021 | 1.3E-08 | 32.3 |
| HbA1c | Women | rs17168486 | T | C | 0.173 | 0.190 | 0.028 | 6.7E-12 | 47.1 |
| HbA1c | Women | rs1974620 | T | C | 0.549 | 0.153 | 0.021 | 3.4E-13 | 53.0 |
| HbA1c | Women | rs62491945 | A | G | 0.347 | -0.125 | 0.022 | 8.7E-09 | 33.1 |
| HbA1c | Women | rs864745 | C | T | 0.506 | -0.170 | 0.021 | 2.6E-16 | 67.1 |
| HbA1c | Women | rs2908277 | A | G | 0.120 | 0.333 | 0.032 | 2.2E-25 | 108.4 |
| HbA1c | Women | rs1799884 | T | C | 0.178 | 0.769 | 0.027 | 2.4E-177 | 807.9 |
| HbA1c | Women | rs868961 | T | G | 0.340 | 0.132 | 0.022 | 1.8E-09 | 36.2 |
| HbA1c | Women | rs6973494 | A | C | 0.516 | 0.120 | 0.021 | 1.3E-08 | 32.3 |
| HbA1c | Women | rs117370443 | C | T | 0.039 | 0.335 | 0.054 | 4.7E-10 | 38.8 |
| HbA1c | Women | rs7820334 | T | C | 0.311 | -0.143 | 0.023 | 2.6E-10 | 40.0 |
| HbA1c | Women | rs55780214 | A | T | 0.290 | -0.192 | 0.023 | 7.1E-17 | 69.7 |
| HbA1c | Women | rs13266634 | T | C | 0.311 | -0.369 | 0.022 | 1.3E-60 | 269.9 |
| HbA1c | Women | rs2060984 | C | T | 0.450 | -0.119 | 0.021 | 1.2E-08 | 32.5 |

| | | | | | | | | | |
|---|---|---|---|---|---|---|---|---|---|
| HbA1c | Women | rs6474359 | C | T | 0.038 | -0.854 | 0.054 | 2.3E-55 | 245.8 |
| HbA1c | Women | rs4737010 | A | G | 0.228 | 0.471 | 0.025 | 1.8E-80 | 361.3 |
| HbA1c | Women | rs55840085 | A | G | 0.355 | 0.157 | 0.022 | 4.8E-13 | 52.3 |
| HbA1c | Women | rs7042939 | G | A | 0.620 | -0.199 | 0.022 | 3.8E-20 | 84.5 |
| HbA1c | Women | rs41273438 | G | A | 0.041 | 0.322 | 0.052 | 7.9E-10 | 37.8 |
| HbA1c | Women | rs5900831 | G | T | 0.653 | 0.142 | 0.022 | 1.5E-10 | 41.0 |
| HbA1c | Women | rs550057 | T | C | 0.255 | 0.287 | 0.024 | 3.8E-33 | 143.9 |
| HbA1c | Women | rs28641468 | C | T | 0.756 | 0.256 | 0.024 | 2.7E-26 | 112.6 |
| HbA1c | Women | rs10811660 | A | G | 0.173 | -0.334 | 0.027 | 4.7E-34 | 148.1 |
| HbA1c | Women | rs2236495 | C | T | 0.208 | -0.192 | 0.026 | 7.9E-14 | 55.8 |
| HbA1c | Women | rs11145347 | C | T | 0.197 | 0.221 | 0.026 | 2.7E-17 | 71.6 |
| HbA1c | Women | rs61750929 | T | C | 0.055 | -0.716 | 0.046 | 2.5E-55 | 245.7 |
| HbA1c | Men | rs114322470 | G | T | 0.019 | -0.565 | 0.086 | 5.1E-11 | 43.1 |
| HbA1c | Men | rs34872471 | C | T | 0.291 | 0.555 | 0.026 | 1.2E-104 | 472.6 |
| HbA1c | Men | rs11257655 | T | C | 0.208 | 0.256 | 0.029 | 3.0E-19 | 80.5 |
| HbA1c | Men | rs7904973 | T | G | 0.576 | 0.135 | 0.023 | 1.0E-08 | 32.8 |
| HbA1c | Men | rs3780962 | G | A | 0.532 | 0.135 | 0.023 | 7.4E-09 | 33.4 |
| HbA1c | Men | rs56197478 | G | A | 0.223 | -0.256 | 0.028 | 9.5E-20 | 82.7 |
| HbA1c | Men | rs550048289 | G | T | 0.015 | -0.830 | 0.102 | 3.9E-16 | 66.3 |
| HbA1c | Men | rs1106676 | G | T | 0.656 | 0.290 | 0.024 | 2.0E-32 | 140.6 |
| HbA1c | Men | rs11812380 | C | T | 0.012 | 0.757 | 0.111 | 7.9E-12 | 46.8 |
| HbA1c | Men | rs10159477 | A | G | 0.149 | -1.770 | 0.032 | 0.0E+00 | 2999.4 |
| HbA1c | Men | rs749108 | T | C | 0.696 | 0.158 | 0.026 | 6.2E-10 | 38.3 |
| HbA1c | Men | rs12219514 | G | A | 0.434 | -0.207 | 0.023 | 1.1E-18 | 78.0 |
| HbA1c | Men | rs1502276 | A | G | 0.184 | -0.176 | 0.030 | 5.4E-09 | 34.0 |
| HbA1c | Men | rs7127313 | T | C | 0.346 | 0.198 | 0.024 | 6.2E-16 | 65.4 |
| HbA1c | Men | rs6484302 | T | C | 0.176 | 0.223 | 0.031 | 4.3E-13 | 52.5 |
| HbA1c | Men | rs7124355 | G | A | 0.675 | -0.138 | 0.025 | 3.1E-08 | 30.7 |
| HbA1c | Men | rs12419995 | T | A | 0.656 | -0.235 | 0.026 | 5.3E-20 | 83.9 |
| HbA1c | Men | rs2237895 | C | A | 0.415 | 0.211 | 0.024 | 3.0E-19 | 80.5 |
| HbA1c | Men | rs11039165 | G | A | 0.271 | -0.167 | 0.026 | 1.6E-10 | 40.9 |
| HbA1c | Men | rs174533 | A | G | 0.347 | -0.192 | 0.024 | 3.5E-15 | 62.0 |
| HbA1c | Men | rs9667947 | C | T | 0.161 | -0.252 | 0.032 | 1.6E-15 | 63.5 |
| HbA1c | Men | rs10830963 | G | C | 0.275 | 0.436 | 0.026 | 1.1E-63 | 284.1 |
| HbA1c | Men | rs360140 | A | C | 0.643 | -0.283 | 0.024 | 2.1E-31 | 135.9 |
| HbA1c | Men | rs7137828 | T | C | 0.517 | 0.200 | 0.023 | 9.5E-18 | 73.6 |
| HbA1c | Men | rs112066678 | C | A | 0.054 | 0.294 | 0.051 | 9.3E-09 | 33.0 |
| HbA1c | Men | rs9668691 | C | T | 0.232 | -0.202 | 0.028 | 2.5E-13 | 53.5 |
| HbA1c | Men | rs4238013 | T | C | 0.796 | -0.165 | 0.029 | 1.5E-08 | 32.1 |
| HbA1c | Men | rs76895963 | G | T | 0.020 | -1.488 | 0.089 | 2.3E-62 | 278.0 |
| HbA1c | Men | rs12819124 | A | C | 0.472 | -0.451 | 0.023 | 2.6E-83 | 374.4 |
| HbA1c | Men | rs6580661 | T | A | 0.224 | 0.215 | 0.028 | 1.4E-14 | 59.3 |
| HbA1c | Men | rs143343646 | G | A | 0.063 | -0.266 | 0.049 | 4.5E-08 | 29.9 |
| HbA1c | Men | rs79755767 | A | G | 0.100 | -0.243 | 0.039 | 5.6E-10 | 38.5 |
| HbA1c | Men | rs343087 | G | A | 0.861 | -0.191 | 0.034 | 1.3E-08 | 32.3 |
| HbA1c | Men | rs79621919 | A | G | 0.059 | -0.299 | 0.049 | 1.5E-09 | 36.6 |
| HbA1c | Men | rs420431 | G | C | 0.122 | 0.482 | 0.035 | 4.0E-42 | 185.1 |
| HbA1c | Men | rs7329468 | T | C | 0.217 | -0.270 | 0.028 | 2.6E-21 | 89.8 |

| | | | | | | | | | |
|---|---|---|---|---|---|---|---|---|---|
| HbA1c | Men | rs11149501 | C | T | 0.223 | 0.203 | 0.028 | 5.2E-13 | 52.1 |
| HbA1c | Men | rs2858980 | A | G | 0.833 | -0.264 | 0.031 | 2.2E-17 | 72.0 |
| HbA1c | Men | rs1359790 | A | G | 0.289 | -0.199 | 0.026 | 6.9E-15 | 60.6 |
| HbA1c | Men | rs561278583 | C | G | 0.206 | 0.168 | 0.029 | 5.3E-09 | 34.1 |
| HbA1c | Men | rs12886807 | T | C | 0.242 | -0.153 | 0.028 | 3.5E-08 | 30.4 |
| HbA1c | Men | rs12434326 | C | G | 0.249 | -0.177 | 0.027 | 4.8E-11 | 43.3 |
| HbA1c | Men | rs2285005 | A | G | 0.340 | 0.203 | 0.025 | 1.6E-16 | 68.1 |
| HbA1c | Men | rs10873398 | A | G | 0.646 | -0.162 | 0.024 | 2.6E-11 | 44.5 |
| HbA1c | Men | rs113204516 | G | A | 0.049 | -0.350 | 0.054 | 1.1E-10 | 41.6 |
| HbA1c | Men | rs7167567 | A | T | 0.423 | 0.155 | 0.024 | 4.6E-11 | 43.3 |
| HbA1c | Men | rs11633054 | G | A | 0.713 | 0.210 | 0.026 | 3.3E-16 | 66.6 |
| HbA1c | Men | rs62022948 | T | C | 0.329 | -0.138 | 0.025 | 2.5E-08 | 31.0 |
| HbA1c | Men | rs1866476 | C | T | 0.732 | -0.170 | 0.026 | 1.0E-10 | 41.8 |
| HbA1c | Men | rs396263 | G | A | 0.211 | 0.161 | 0.029 | 1.5E-08 | 32.1 |
| HbA1c | Men | rs165975 | C | T | 0.234 | 0.173 | 0.028 | 3.2E-10 | 39.6 |
| HbA1c | Men | rs184499898 | G | A | 0.011 | 0.715 | 0.116 | 6.4E-10 | 38.2 |
| HbA1c | Men | rs181205 | C | T | 0.343 | 0.208 | 0.025 | 3.9E-17 | 70.8 |
| HbA1c | Men | rs2858010 | C | T | 0.340 | -0.185 | 0.025 | 5.8E-14 | 56.4 |
| HbA1c | Men | rs8050500 | C | T | 0.446 | -0.199 | 0.023 | 2.0E-17 | 72.1 |
| HbA1c | Men | rs7188250 | C | T | 0.414 | 0.175 | 0.024 | 1.3E-13 | 54.9 |
| HbA1c | Men | rs35158985 | G | A | 0.308 | 0.162 | 0.025 | 1.3E-10 | 41.3 |
| HbA1c | Men | rs247826 | T | C | 0.223 | 0.229 | 0.028 | 3.0E-16 | 66.8 |
| HbA1c | Men | rs72811487 | A | G | 0.035 | -0.357 | 0.063 | 1.8E-08 | 31.7 |
| HbA1c | Men | rs551118 | G | C | 0.578 | 0.394 | 0.024 | 2.9E-61 | 272.9 |
| HbA1c | Men | rs1108646 | G | A | 0.682 | -0.243 | 0.025 | 4.8E-22 | 93.2 |
| HbA1c | Men | rs71373617 | T | C | 0.320 | -0.162 | 0.025 | 9.4E-11 | 42.0 |
| HbA1c | Men | rs216195 | G | T | 0.303 | 0.154 | 0.025 | 9.9E-10 | 37.3 |
| HbA1c | Men | rs9303620 | C | T | 0.278 | -0.201 | 0.026 | 9.3E-15 | 60.1 |
| HbA1c | Men | rs17698176 | G | T | 0.229 | 0.173 | 0.028 | 6.0E-10 | 38.3 |
| HbA1c | Men | rs12600858 | A | G | 0.224 | -0.237 | 0.028 | 2.2E-17 | 72.0 |
| HbA1c | Men | rs8064261 | A | G | 0.275 | -0.157 | 0.026 | 1.8E-09 | 36.2 |
| HbA1c | Men | rs1030097 | G | C | 0.495 | 0.128 | 0.023 | 4.1E-08 | 30.1 |
| HbA1c | Men | rs1641523 | T | C | 0.567 | -0.245 | 0.024 | 3.8E-25 | 107.4 |
| HbA1c | Men | rs2748427 | G | A | 0.218 | 0.436 | 0.028 | 3.6E-54 | 240.4 |
| HbA1c | Men | rs113373052 | T | C | 0.313 | 0.463 | 0.025 | 7.7E-76 | 340.0 |
| HbA1c | Men | rs8078338 | G | A | 0.506 | -0.169 | 0.023 | 4.0E-13 | 52.6 |
| HbA1c | Men | rs6507691 | C | T | 0.749 | 0.184 | 0.027 | 7.7E-12 | 46.8 |
| HbA1c | Men | rs17533945 | C | T | 0.392 | 0.337 | 0.024 | 6.9E-45 | 197.7 |
| HbA1c | Men | rs10401969 | C | T | 0.077 | 0.263 | 0.044 | 2.0E-09 | 36.0 |
| HbA1c | Men | rs75372982 | G | A | 0.289 | -0.175 | 0.026 | 1.6E-11 | 45.4 |
| HbA1c | Men | rs584007 | G | A | 0.641 | -0.184 | 0.024 | 3.6E-14 | 57.4 |
| HbA1c | Men | rs35255921 | A | G | 0.013 | -0.581 | 0.101 | 9.2E-09 | 33.0 |
| HbA1c | Men | rs62111724 | C | T | 0.366 | -0.207 | 0.024 | 2.4E-17 | 71.8 |
| HbA1c | Men | rs12985029 | A | G | 0.340 | -0.157 | 0.025 | 1.6E-10 | 40.9 |
| HbA1c | Men | rs1975283 | C | A | 0.685 | -0.164 | 0.025 | 6.6E-11 | 42.6 |
| HbA1c | Men | rs587739738 | T | C | 0.083 | -0.284 | 0.042 | 1.8E-11 | 45.2 |
| HbA1c | Men | rs857720 | C | T | 0.267 | 0.591 | 0.026 | 4.1E-112 | 507.1 |
| HbA1c | Men | rs2228445 | C | T | 0.902 | 0.283 | 0.039 | 4.5E-13 | 52.4 |

| | | | | | | | | | |
|---|---|---|---|---|---|---|---|---|---|
| HbA1c | Men | rs79687284 | C | G | 0.035 | 0.603 | 0.063 | 1.5E-21 | 90.9 |
| HbA1c | Men | rs238763 | A | T | 0.592 | -0.169 | 0.024 | 1.1E-12 | 50.6 |
| HbA1c | Men | rs3811444 | T | C | 0.334 | 0.192 | 0.025 | 5.8E-15 | 61.0 |
| HbA1c | Men | rs4518840 | T | A | 0.878 | -0.279 | 0.036 | 9.2E-15 | 60.1 |
| HbA1c | Men | rs114247628 | A | G | 0.006 | -1.373 | 0.159 | 5.1E-18 | 74.9 |
| HbA1c | Men | rs1175550 | G | A | 0.230 | -0.304 | 0.028 | 2.4E-27 | 117.4 |
| HbA1c | Men | rs3768321 | T | G | 0.197 | 0.212 | 0.029 | 4.6E-13 | 52.4 |
| HbA1c | Men | rs3789586 | T | C | 0.153 | -0.248 | 0.032 | 1.7E-14 | 58.9 |
| HbA1c | Men | rs7513688 | A | G | 0.361 | 0.149 | 0.024 | 9.3E-10 | 37.5 |
| HbA1c | Men | rs2065703 | T | C | 0.159 | 0.179 | 0.032 | 1.9E-08 | 31.6 |
| HbA1c | Men | rs6063050 | C | T | 0.283 | -0.162 | 0.026 | 4.8E-10 | 38.8 |
| HbA1c | Men | rs6014993 | G | A | 0.487 | 0.146 | 0.023 | 4.0E-10 | 39.1 |
| HbA1c | Men | rs2427361 | A | C | 0.620 | -0.151 | 0.024 | 4.9E-10 | 38.7 |
| HbA1c | Men | rs6086540 | C | G | 0.507 | -0.173 | 0.023 | 1.2E-13 | 55.0 |
| HbA1c | Men | rs17850433 | C | T | 0.012 | 0.647 | 0.108 | 1.8E-09 | 36.2 |
| HbA1c | Men | rs59923178 | G | T | 0.096 | -0.251 | 0.039 | 2.0E-10 | 40.5 |
| HbA1c | Men | rs77696994 | T | C | 0.076 | -0.301 | 0.044 | 1.0E-11 | 46.3 |
| HbA1c | Men | rs855791 | G | A | 0.561 | -0.428 | 0.023 | 3.3E-74 | 332.5 |
| HbA1c | Men | rs8138197 | A | G | 0.471 | -0.210 | 0.023 | 1.7E-19 | 81.6 |
| HbA1c | Men | rs830021 | C | A | 0.697 | -0.153 | 0.025 | 1.7E-09 | 36.3 |
| HbA1c | Men | rs560887 | C | T | 0.701 | 0.685 | 0.025 | 1.1E-161 | 735.8 |
| HbA1c | Men | rs13031216 | T | C | 0.220 | -0.195 | 0.028 | 3.4E-12 | 48.4 |
| HbA1c | Men | rs1399623 | C | A | 0.623 | 0.186 | 0.024 | 1.7E-14 | 58.9 |
| HbA1c | Men | rs838717 | A | G | 0.566 | 0.133 | 0.023 | 1.3E-08 | 32.3 |
| HbA1c | Men | rs113362373 | G | A | 0.135 | 0.269 | 0.034 | 2.9E-15 | 62.3 |
| HbA1c | Men | rs780093 | C | T | 0.618 | 0.237 | 0.024 | 3.2E-23 | 98.6 |
| HbA1c | Men | rs10173636 | G | C | 0.130 | -0.456 | 0.035 | 8.9E-40 | 174.3 |
| HbA1c | Men | rs2164701 | C | G | 0.140 | 0.235 | 0.034 | 2.6E-12 | 49.0 |
| HbA1c | Men | rs77000664 | G | A | 0.177 | 0.169 | 0.031 | 3.5E-08 | 30.4 |
| HbA1c | Men | rs7606173 | C | G | 0.431 | -0.182 | 0.023 | 8.1E-15 | 60.3 |
| HbA1c | Men | rs7572278 | A | T | 0.204 | 0.214 | 0.029 | 1.1E-13 | 55.2 |
| HbA1c | Men | rs7638232 | A | G | 0.418 | -0.263 | 0.024 | 6.3E-29 | 124.6 |
| HbA1c | Men | rs11708067 | G | A | 0.245 | -0.313 | 0.027 | 6.0E-31 | 133.9 |
| HbA1c | Men | rs12497133 | A | G | 0.533 | 0.148 | 0.023 | 2.6E-10 | 39.9 |
| HbA1c | Men | rs6785881 | T | C | 0.481 | -0.153 | 0.023 | 5.8E-11 | 42.9 |
| HbA1c | Men | rs1905505 | A | G | 0.284 | -0.283 | 0.026 | 5.2E-28 | 120.4 |
| HbA1c | Men | rs12633493 | T | C | 0.586 | 0.184 | 0.024 | 7.6E-15 | 60.5 |
| HbA1c | Men | rs6780171 | A | T | 0.313 | 0.210 | 0.025 | 6.9E-17 | 69.7 |
| HbA1c | Men | rs2041965 | T | C | 0.344 | -0.144 | 0.024 | 3.9E-09 | 34.7 |
| HbA1c | Men | rs6777684 | G | A | 0.610 | 0.178 | 0.024 | 1.2E-13 | 55.0 |
| HbA1c | Men | rs1496653 | G | A | 0.204 | -0.159 | 0.029 | 3.5E-08 | 30.4 |
| HbA1c | Men | rs12715435 | A | G | 0.171 | 0.267 | 0.031 | 7.8E-18 | 74.0 |
| HbA1c | Men | rs11728350 | G | A | 0.125 | 0.198 | 0.035 | 1.8E-08 | 31.7 |
| HbA1c | Men | rs9994509 | G | A | 0.393 | -0.134 | 0.024 | 1.9E-08 | 31.6 |
| HbA1c | Men | rs2044341 | C | T | 0.684 | -0.358 | 0.025 | 2.5E-46 | 204.4 |
| HbA1c | Men | rs17462188 | T | C | 0.690 | 0.152 | 0.025 | 2.2E-09 | 35.8 |
| HbA1c | Men | rs111537480 | G | A | 0.138 | -0.216 | 0.034 | 2.4E-10 | 40.1 |
| HbA1c | Men | rs225176 | G | C | 0.671 | -0.146 | 0.026 | 1.3E-08 | 32.3 |

| | | | | | | | | | |
|---|---|---|---|---|---|---|---|---|---|
| HbA1c | Men | rs59816691 | G | C | 0.669 | 0.137 | 0.025 | 3.5E-08 | 30.4 |
| HbA1c | Men | rs258217 | G | A | 0.464 | -0.157 | 0.024 | 5.1E-11 | 43.2 |
| HbA1c | Men | rs7720275 | C | T | 0.170 | 0.180 | 0.031 | 6.9E-09 | 33.6 |
| HbA1c | Men | rs7732130 | A | G | 0.681 | -0.249 | 0.025 | 1.4E-23 | 100.1 |
| HbA1c | Men | rs34533653 | A | T | 0.266 | -0.149 | 0.026 | 1.5E-08 | 32.0 |
| HbA1c | Men | rs12210538 | G | A | 0.236 | -0.183 | 0.027 | 2.1E-11 | 44.8 |
| HbA1c | Men | rs56293029 | A | C | 0.259 | -0.308 | 0.027 | 2.0E-30 | 131.4 |
| HbA1c | Men | rs62440928 | A | G | 0.607 | -0.158 | 0.024 | 5.1E-11 | 43.2 |
| HbA1c | Men | rs12201779 | T | C | 0.395 | -0.143 | 0.024 | 2.2E-09 | 35.8 |
| HbA1c | Men | rs2665357 | C | A | 0.517 | 0.142 | 0.023 | 1.1E-09 | 37.1 |
| HbA1c | Men | rs67131976 | T | C | 0.172 | 0.355 | 0.031 | 9.1E-31 | 133.0 |
| HbA1c | Men | rs1800562 | A | G | 0.079 | -0.815 | 0.043 | 2.7E-79 | 355.9 |
| HbA1c | Men | rs204991 | C | T | 0.221 | 0.161 | 0.028 | 9.5E-09 | 32.9 |
| HbA1c | Men | rs3135365 | A | C | 0.769 | 0.257 | 0.028 | 1.2E-20 | 86.9 |
| HbA1c | Men | rs2308891 | A | C | 0.459 | 0.164 | 0.024 | 1.2E-11 | 46.1 |
| HbA1c | Men | rs3778321 | A | G | 0.180 | -0.180 | 0.030 | 2.7E-09 | 35.4 |
| HbA1c | Men | rs12667932 | A | G | 0.080 | -0.347 | 0.043 | 7.0E-16 | 65.2 |
| HbA1c | Men | rs11763864 | T | A | 0.157 | 0.218 | 0.032 | 8.8E-12 | 46.6 |
| HbA1c | Men | rs11514706 | C | A | 0.524 | 0.223 | 0.023 | 1.3E-21 | 91.2 |
| HbA1c | Men | rs11982272 | C | T | 0.202 | -0.160 | 0.029 | 4.2E-08 | 30.0 |
| HbA1c | Men | rs1708302 | T | C | 0.501 | -0.192 | 0.023 | 1.6E-16 | 68.0 |
| HbA1c | Men | rs76323047 | G | A | 0.118 | 0.336 | 0.036 | 1.1E-20 | 86.9 |
| HbA1c | Men | rs1004558 | T | C | 0.179 | 0.685 | 0.030 | 1.0E-112 | 509.9 |
| HbA1c | Men | rs55803206 | A | C | 0.464 | 0.133 | 0.024 | 2.0E-08 | 31.5 |
| HbA1c | Men | rs117370443 | C | T | 0.039 | 0.419 | 0.060 | 3.1E-12 | 48.6 |
| HbA1c | Men | rs4300038 | A | G | 0.311 | -0.394 | 0.025 | 3.9E-55 | 244.8 |
| HbA1c | Men | rs2954021 | G | A | 0.506 | 0.211 | 0.023 | 1.0E-19 | 82.5 |
| HbA1c | Men | rs2060985 | G | A | 0.537 | -0.130 | 0.023 | 2.2E-08 | 31.3 |
| HbA1c | Men | rs3757974 | G | A | 0.380 | 0.148 | 0.024 | 5.4E-10 | 38.5 |
| HbA1c | Men | rs66593272 | T | A | 0.038 | -0.982 | 0.061 | 2.9E-58 | 259.2 |
| HbA1c | Men | rs4737010 | A | G | 0.228 | 0.603 | 0.028 | 1.5E-104 | 472.3 |
| HbA1c | Men | rs55840085 | A | G | 0.355 | 0.184 | 0.024 | 3.7E-14 | 57.3 |
| HbA1c | Men | rs7829826 | A | T | 0.534 | -0.145 | 0.023 | 4.9E-10 | 38.7 |
| HbA1c | Men | rs1967604 | G | A | 0.712 | -0.210 | 0.026 | 3.4E-16 | 66.6 |
| HbA1c | Men | rs550057 | T | C | 0.255 | 0.368 | 0.027 | 3.3E-43 | 190.0 |
| HbA1c | Men | rs28641468 | C | T | 0.756 | 0.211 | 0.027 | 5.8E-15 | 61.0 |
| HbA1c | Men | rs10811660 | A | G | 0.173 | -0.387 | 0.031 | 2.7E-36 | 158.4 |
| HbA1c | Men | rs10758593 | A | G | 0.397 | 0.141 | 0.024 | 2.8E-09 | 35.3 |
| HbA1c | Men | rs17803780 | C | T | 0.211 | -0.169 | 0.028 | 2.7E-09 | 35.4 |
| HbA1c | Men | rs7025532 | C | T | 0.199 | 0.261 | 0.029 | 2.4E-19 | 80.9 |
| HbA1c | Men | rs61750929 | T | C | 0.055 | -0.668 | 0.051 | 2.8E-39 | 172.0 |
| HbA1c | Men | rs7047279 | C | T | 0.568 | -0.161 | 0.024 | 9.1E-12 | 46.5 |

BMI, body mass index; HbA1c, glycated hemoglobin.

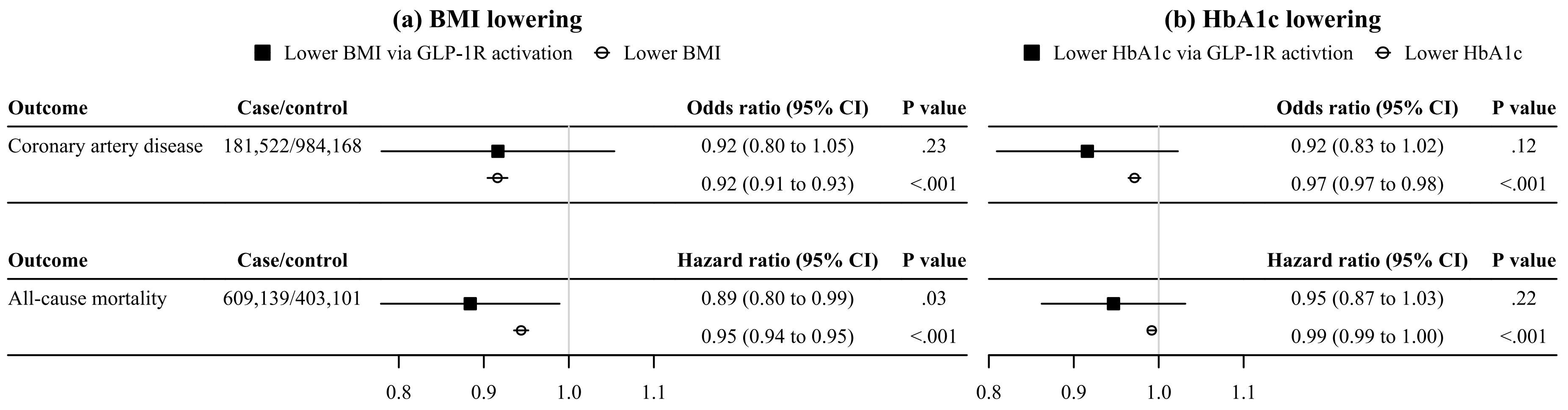

**eFigure 1. IVW MR estimates for the associations of GLP-1R activation with the risk of coronary artery disease and all-cause mortality in comparison with those for lower BMI and lower HbA1c.**

BMI, body mass index; GLP-1R, glucagon-like peptide-1 receptor; HbA1c, glycated hemoglobin; IVW, inverse variance weighted; MR, Mendelian randomization. Black squares denote genetically predicted (a) lower BMI or (b) lower HbA1c via GLP-1R activation based on *GLP1R* variants. White circles denote genetically predicted (a) lower BMI or (b) lower HbA1c based on genome-wide variants. Estimates are presented as odds ratio for coronary artery disease or hazard ratio for all-cause mortality per 1-kg/m$^2$ decrease in BMI or per 1-mmol/mol decrease in HbA1c.

**eTable 5. Sensitivity MR analyses for the associations of GLP-1R activation, lower BMI, and lower HbA1c with the risk of coronary artery disease and all-cause mortality.**

| Exposure | Outcome | SNPs | Method | Estimate (95% CI) | Measure | *P* value | *P* value (intercept) | *P* value (Q) |
|---|---|---|---|---|---|---|---|---|
| Lower BMI via GLP-1R activation | Coronary artery disease | 3 | IVW | 0.94 (0.84 to 1.07) | Odds ratio | 0.36 | | 0.64 |
| Lower BMI via GLP-1R activation | Coronary artery disease | 3 | Weighted median | 0.94 (0.81 to 1.08) | Odds ratio | 0.38 | | |
| Lower BMI via GLP-1R activation | Coronary artery disease | 3 | MR Egger | 0.70 (0.35 to 1.37) | Odds ratio | 0.29 | 0.37 | |
| Lower BMI via GLP-1R activation | Coronary artery disease | 3 | MR-RAPS | 0.94 (0.83 to 1.08) | Odds ratio | 0.39 | | |
| Lower BMI via GLP-1R activation | All-cause mortality | 3 | IVW | 0.91 (0.83 to 1.00) | Hazard ratio | 0.05 | | 0.11 |
| Lower BMI via GLP-1R activation | All-cause mortality | 3 | Weighted median | 0.94 (0.83 to 1.07) | Hazard ratio | 0.34 | | |
| Lower BMI via GLP-1R activation | All-cause mortality | 3 | MR Egger | 1.12 (0.41 to 3.02) | Hazard ratio | 0.83 | 0.68 | |
| Lower BMI via GLP-1R activation | All-cause mortality | 3 | MR-RAPS | 0.92 (0.83 to 1.01) | Hazard ratio | 0.09 | | |
| Lower BMI | Coronary artery disease | 505 | IVW | 0.92 (0.91 to 0.93) | Odds ratio | <0.001 | | <0.001 |
| Lower BMI | Coronary artery disease | 505 | Weighted median | 0.92 (0.91 to 0.93) | Odds ratio | <0.001 | | |
| Lower BMI | Coronary artery disease | 505 | MR Egger | 0.92 (0.90 to 0.95) | Odds ratio | <0.001 | 0.74 | |
| Lower BMI | Coronary artery disease | 505 | MR-RAPS | 0.91 (0.91 to 0.92) | Odds ratio | <0.001 | | |
| Lower BMI | All-cause mortality | 490 | IVW | 0.95 (0.94 to 0.95) | Hazard ratio | <0.001 | | <0.001 |
| Lower BMI | All-cause mortality | 490 | Weighted median | 0.94 (0.93 to 0.95) | Hazard ratio | <0.001 | | |
| Lower BMI | All-cause mortality | 490 | MR Egger | 0.95 (0.93 to 0.97) | Hazard ratio | <0.001 | 0.51 | |
| Lower BMI | All-cause mortality | 490 | MR-RAPS | 0.94 (0.93 to 0.94) | Hazard ratio | <0.001 | | |
| Lower BMI | All-cause mortality | 489 | IVW (Steiger filtering) | 0.94 (0.94 to 0.95) | Hazard ratio | <0.001 | | <0.001 |
| Lower BMI | All-cause mortality | 489 | Weighted median (Steiger filtering) | 0.94 (0.93 to 0.95) | Hazard ratio | <0.001 | | |
| Lower BMI | All-cause mortality | 489 | MR Egger (Steiger filtering) | 0.94 (0.93 to 0.96) | Hazard ratio | <0.001 | 0.86 | |
| Lower BMI | All-cause mortality | 489 | MR-RAPS (Steiger filtering) | 0.94 (0.93 to 0.94) | Hazard ratio | <0.001 | | |

| | | | | | | | | |
|---|---|---|---|---|---|---|---|---|
| Lower HbA1c via GLP-1R activation | Coronary artery disease | 2 | IVW | 0.88 (0.80 to 0.96) | Odds ratio | 0.003 | | 0.13 |
| Lower HbA1c via GLP-1R activation | All-cause mortality | 2 | IVW | 0.93 (0.87 to 1.00) | Hazard ratio | 0.04 | | 0.59 |
| Lower HbA1c | Coronary artery disease | 303 | IVW | 0.97 (0.97 to 0.98) | Odds ratio | <0.001 | | <0.001 |
| Lower HbA1c | Coronary artery disease | 303 | Weighted median | 0.98 (0.97 to 0.98) | Odds ratio | <0.001 | | |
| Lower HbA1c | Coronary artery disease | 303 | MR Egger | 0.99 (0.97 to 1.00) | Odds ratio | 0.04 | 0.006 | |
| Lower HbA1c | Coronary artery disease | 303 | MR-RAPS | 0.97 (0.97 to 0.98) | Odds ratio | <0.001 | | |
| Lower HbA1c | All-cause mortality | 286 | IVW | 0.99 (0.99 to 1.00) | Hazard ratio | <0.001 | | <0.001 |
| Lower HbA1c | All-cause mortality | 286 | Weighted median | 1.00 (0.99 to 1.00) | Hazard ratio | 0.05 | | |
| Lower HbA1c | All-cause mortality | 286 | MR Egger | 1.00 (1.00 to 1.01) | Hazard ratio | 0.14 | <0.001 | |
| Lower HbA1c | All-cause mortality | 286 | MR-RAPS | 0.99 (0.99 to 0.99) | Hazard ratio | <0.001 | | |

BMI, body mass index; GLP-1R, glucagon-like peptide-1 receptor; HbA1c, glycated hemoglobin; IVW, inverse variance weighted; MR, Mendelian randomization; RAPS, robust adjusted profile score. Estimates are presented per 1-kg/$m^2$ decrease in BMI or per 1-mmol/mol decrease in HbA1c.

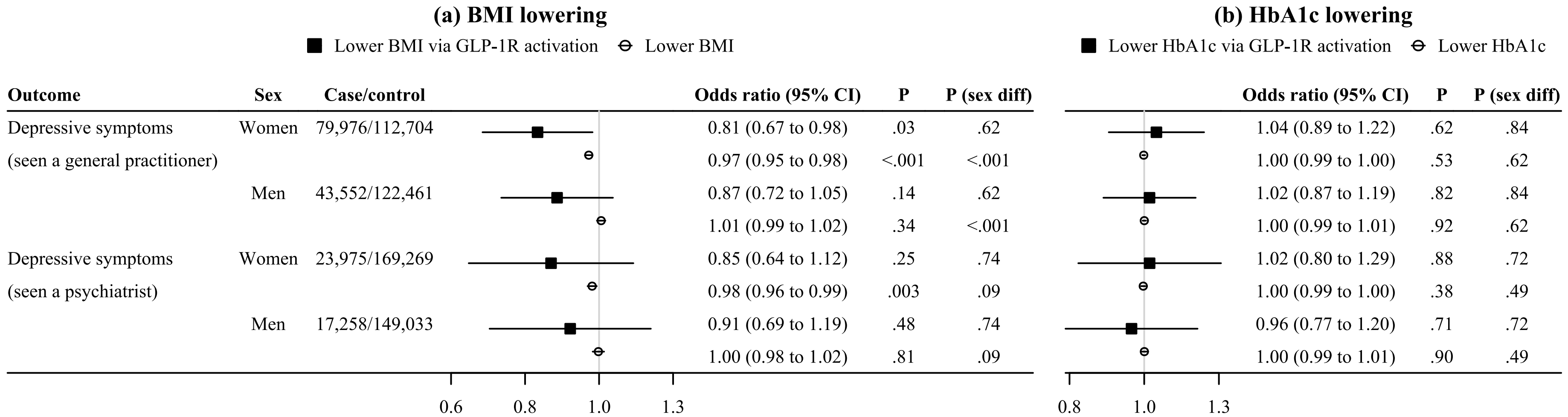

| Outcome | Sex | Case/control | (a) Odds ratio (95% CI) | (a) P | (a) P (sex diff) | (b) Odds ratio (95% CI) | (b) P | (b) P (sex diff) |
|---|---|---|---|---|---|---|---|---|
| Depressive symptoms | Women | 79,976/112,704 | 0.81 (0.67 to 0.98) | .03 | .62 | 1.04 (0.89 to 1.22) | .62 | .84 |
| (seen a general practitioner) | | | 0.97 (0.95 to 0.98) | <.001 | <.001 | 1.00 (0.99 to 1.00) | .53 | .62 |
| | Men | 43,552/122,461 | 0.87 (0.72 to 1.05) | .14 | .62 | 1.02 (0.87 to 1.19) | .82 | .84 |
| | | | 1.01 (0.99 to 1.02) | .34 | <.001 | 1.00 (0.99 to 1.01) | .92 | .62 |
| Depressive symptoms | Women | 23,975/169,269 | 0.85 (0.64 to 1.12) | .25 | .74 | 1.02 (0.80 to 1.29) | .88 | .72 |
| (seen a psychiatrist) | | | 0.98 (0.96 to 0.99) | .003 | .09 | 1.00 (0.99 to 1.00) | .38 | .49 |
| | Men | 17,258/149,033 | 0.91 (0.69 to 1.19) | .48 | .74 | 0.96 (0.77 to 1.20) | .71 | .72 |
| | | | 1.00 (0.98 to 1.02) | .81 | .09 | 1.00 (0.99 to 1.01) | .90 | .49 |

**eFigure 2. IVW MR estimates for sex-specific associations of GLP-1R activation with the risk of depressive symptoms in comparison with those for lower BMI and lower HbA1c.**

BMI, body mass index; GLP-1R, glucagon-like peptide-1 receptor; HbA1c, glycated hemoglobin; IVW, inverse variance weighted; MR, Mendelian randomization. Black squares denote genetically predicted (a) lower BMI or (b) lower HbA1c via GLP-1R activation based on *GLP1R* variants. White circles denote genetically predicted (a) lower BMI or (b) lower HbA1c based on genome-wide variants. Estimates are presented as odds ratio per 1-kg/$m^2$ decrease in BMI or per 1-mmol/mol decrease in HbA1c.

**eTable 6. Sensitivity MR analyses for the associations of GLP-1R activation, lower BMI, and lower HbA1c with mental health well-being.**

| Exposure | Outcome | SNPs | Method | Beta (95% CI) | *P* value | *P* value (intercept) | *P* value (Q) |
|---|---|---|---|---|---|---|---|
| Lower BMI via GLP-1R activation | Well-being spectrum | 3 | IVW | 0.04 (0.02 to 0.07) | <0.001 | | 0.01 |
| Lower BMI via GLP-1R activation | Well-being spectrum | 3 | Weighted median | 0.04 (0.01 to 0.07) | 0.01 | | |
| Lower BMI via GLP-1R activation | Well-being spectrum | 3 | MR Egger | 0.04 (-0.35 to 0.43) | 0.84 | 0.99 | |
| Lower BMI via GLP-1R activation | Well-being spectrum | 3 | MR-RAPS | 0.04 (0.02 to 0.07) | <0.001 | | |
| Lower BMI via GLP-1R activation | Life satisfaction | 3 | IVW | 0.06 (0.03 to 0.09) | <0.001 | | 0.01 |
| Lower BMI via GLP-1R activation | Life satisfaction | 3 | Weighted median | 0.06 (0.01 to 0.10) | 0.03 | | |
| Lower BMI via GLP-1R activation | Life satisfaction | 3 | MR Egger | 0.06 (-0.51 to 0.64) | 0.83 | 0.99 | |
| Lower BMI via GLP-1R activation | Life satisfaction | 3 | MR-RAPS | 0.06 (0.02 to 0.09) | 0.00 | | |
| Lower BMI via GLP-1R activation | Positive affect | 3 | IVW | 0.06 (0.03 to 0.08) | <0.001 | | 0.01 |
| Lower BMI via GLP-1R activation | Positive affect | 3 | Weighted median | 0.05 (0.01 to 0.09) | 0.01 | | |
| Lower BMI via GLP-1R activation | Positive affect | 3 | MR Egger | 0.05 (-0.44 to 0.54) | 0.84 | 0.98 | |
| Lower BMI via GLP-1R activation | Positive affect | 3 | MR-RAPS | 0.05 (0.02 to 0.08) | 0.00 | | |
| Lower BMI via GLP-1R activation | Neuroticism | 3 | IVW | -0.07 (-0.10 to -0.03) | <0.001 | | 0.01 |
| Lower BMI via GLP-1R activation | Neuroticism | 3 | Weighted median | -0.06 (-0.12 to -0.01) | 0.02 | | |
| Lower BMI via GLP-1R activation | Neuroticism | 3 | MR Egger | -0.06 (-0.67 to 0.54) | 0.83 | 0.99 | |
| Lower BMI via GLP-1R activation | Neuroticism | 3 | MR-RAPS | -0.07 (-0.11 to -0.03) | 0.00 | | |
| Lower BMI via GLP-1R activation | Depressive symptoms | 3 | IVW | -0.05 (-0.07 to -0.02) | <0.001 | | 0.01 |
| Lower BMI via GLP-1R activation | Depressive symptoms | 3 | Weighted median | -0.04 (-0.08 to -0.01) | 0.01 | | |
| Lower BMI via GLP-1R activation | Depressive symptoms | 3 | MR Egger | -0.04 (-0.44 to 0.36) | 0.84 | 0.98 | |
| Lower BMI via GLP-1R activation | Depressive symptoms | 3 | MR-RAPS | -0.04 (-0.07 to -0.02) | <0.001 | | |
| Lower BMI | Well-being spectrum | 464 | IVW | 0.003 (0.000 to 0.01) | 0.04 | | <0.001 |
| Lower BMI | Well-being spectrum | 464 | Weighted median | 0.002 (-0.000 to 0.005) | 0.08 | | |
| Lower BMI | Well-being spectrum | 464 | MR Egger | -0.001 (-0.01 to 0.01) | 0.81 | 0.30 | |
| Lower BMI | Well-being spectrum | 464 | MR-RAPS | 0.003 (0.002 to 0.005) | <0.001 | | |

| | | | | | | | |
|---|---|---|---|---|---|---|---|
| Lower BMI | Life satisfaction | 464 | IVW | 0.003 (-0.000 to 0.01) | 0.05 | | <0.001 |
| Lower BMI | Life satisfaction | 464 | Weighted median | 0.002 (-0.001 to 0.01) | 0.24 | | |
| Lower BMI | Life satisfaction | 464 | MR Egger | -0.001 (-0.01 to 0.01) | 0.80 | 0.33 | |
| Lower BMI | Life satisfaction | 464 | MR-RAPS | 0.004 (0.002 to 0.01) | <0.001 | | |
| Lower BMI | Positive affect | 464 | IVW | 0.002 (-0.001 to 0.01) | 0.16 | | <0.001 |
| Lower BMI | Positive affect | 464 | Weighted median | 0.002 (-0.002 to 0.01) | 0.29 | | |
| Lower BMI | Positive affect | 464 | MR Egger | -0.001 (-0.01 to 0.01) | 0.76 | 0.41 | |
| Lower BMI | Positive affect | 464 | MR-RAPS | 0.004 (0.002 to 0.01) | <0.001 | | |
| Lower BMI | Neuroticism | 464 | IVW | -0.004 (-0.01 to 0.000) | 0.05 | | <0.001 |
| Lower BMI | Neuroticism | 464 | Weighted median | -0.005 (-0.01 to -0.001) | 0.02 | | |
| Lower BMI | Neuroticism | 464 | MR Egger | 0.002 (-0.01 to 0.01) | 0.71 | 0.27 | |
| Lower BMI | Neuroticism | 464 | MR-RAPS | -0.01 (-0.01 to -0.003) | <0.001 | | |
| Lower BMI | Depressive symptoms | 464 | IVW | -0.004 (-0.01 to -0.001) | 0.00 | | <0.001 |
| Lower BMI | Depressive symptoms | 464 | Weighted median | -0.003 (-0.01 to -0.001) | 0.02 | | |
| Lower BMI | Depressive symptoms | 464 | MR Egger | -0.001 (-0.01 to 0.01) | 0.85 | 0.35 | |
| Lower BMI | Depressive symptoms | 464 | MR-RAPS | -0.005 (-0.01 to -0.003) | <0.001 | | |
| Lower HbA1c via GLP-1R activation | Well-being spectrum | 1 | IVW | 0.003 (-0.02 to 0.02) | 0.81 | | |
| Lower HbA1c via GLP-1R activation | Life satisfaction | 1 | IVW | 0.002 (-0.03 to 0.03) | 0.90 | | |
| Lower HbA1c via GLP-1R activation | Positive affect | 1 | IVW | 0.002 (-0.02 to 0.03) | 0.88 | | |
| Lower HbA1c via GLP-1R activation | Neuroticism | 1 | IVW | -0.003 (-0.04 to 0.03) | 0.88 | | |
| Lower HbA1c via GLP-1R activation | Depressive symptoms | 1 | IVW | -0.001 (-0.02 to 0.02) | 0.90 | | |
| Lower HbA1c | Well-being spectrum | 242 | IVW | -0.001 (-0.002 to 0.001) | 0.35 | | <0.001 |
| Lower HbA1c | Well-being spectrum | 242 | Weighted median | 0.000 (-0.001 to 0.001) | 0.83 | | |
| Lower HbA1c | Well-being spectrum | 242 | MR Egger | -0.000 (-0.002 to 0.002) | 0.78 | 0.81 | |
| Lower HbA1c | Well-being spectrum | 242 | MR-RAPS | -0.000 (-0.001 to 0.000) | 0.42 | | |
| Lower HbA1c | Life satisfaction | 242 | IVW | -0.001 (-0.002 to 0.001) | 0.42 | | <0.001 |
| Lower HbA1c | Life satisfaction | 242 | Weighted median | -0.000 (-0.002 to 0.002) | 0.96 | | |
| Lower HbA1c | Life satisfaction | 242 | MR Egger | -0.000 (-0.003 to 0.002) | 0.73 | 0.93 | |

| | | | | | | | |
|---|---|---|---|---|---|---|---|
| Lower HbA1c | Life satisfaction | 242 | MR-RAPS | -0.000 (-0.001 to 0.001) | 0.49 | | |
| Lower HbA1c | Positive affect | 242 | IVW | -0.001 (-0.002 to 0.001) | 0.38 | | <0.001 |
| Lower HbA1c | Positive affect | 242 | Weighted median | -0.000 (-0.002 to 0.002) | 0.88 | | |
| Lower HbA1c | Positive affect | 242 | MR Egger | -0.000 (-0.003 to 0.002) | 0.74 | 0.88 | |
| Lower HbA1c | Positive affect | 242 | MR-RAPS | -0.000 (-0.001 to 0.000) | 0.34 | | |
| Lower HbA1c | Neuroticism | 242 | IVW | 0.001 (-0.001 to 0.002) | 0.54 | | <0.001 |
| Lower HbA1c | Neuroticism | 242 | Weighted median | -0.000 (-0.002 to 0.002) | 0.74 | | |
| Lower HbA1c | Neuroticism | 242 | MR Egger | 0.000 (-0.003 to 0.004) | 0.81 | 0.93 | |
| Lower HbA1c | Neuroticism | 242 | MR-RAPS | 0.000 (-0.001 to 0.001) | 0.63 | | |
| Lower HbA1c | Depressive symptoms | 242 | IVW | 0.000 (-0.001 to 0.001) | 0.63 | | <0.001 |
| Lower HbA1c | Depressive symptoms | 242 | Weighted median | -0.000 (-0.001 to 0.001) | 0.77 | | |
| Lower HbA1c | Depressive symptoms | 242 | MR Egger | 0.000 (-0.002 to 0.003) | 0.68 | 0.85 | |
| Lower HbA1c | Depressive symptoms | 242 | MR-RAPS | 0.000 (-0.001 to 0.001) | 0.70 | | |

BMI, body mass index; GLP-1R, glucagon-like peptide-1 receptor; HbA1c, glycated hemoglobin; IVW, inverse variance weighting; MR, Mendelian randomization; RAPS, robust adjusted profile score. Positive associations with the well-being spectrum, life satisfaction, and positive affect and negative associations with neuroticism and depressive symptoms indicate better mental health well-being. Estimates are presented in standard deviation units per 1-kg/$m^2$ decrease in BMI or per 1-mmol/mol decrease in HbA1c. Only one variant (rs10305518) was used to predict lower HbA1c via GLP-1R activation because rs112385083 and its proxy were not available in the outcome data.

**eTable 7. Sensitivity MR analyses for sex-specific associations of GLP-1R activation, lower BMI, and HbA1c with the risk of depression symptoms.**

| Exposure | Outcome | Sex | SNPs | Method | Odds ratio (95% CI) | *P* value | *P* value (intercept) | *P* value (Q) |
|---|---|---|---|---|---|---|---|---|
| Lower BMI via GLP-1R activation | Depressive symptoms (seen a general practitioner) | Women | 3 | IVW | 0.81 (0.68 to 0.97) | 0.02 | | 0.65 |
| Lower BMI via GLP-1R activation | Depressive symptoms (seen a general practitioner) | Women | 3 | Weighted median | 0.81 (0.62 to 1.07) | 0.14 | | |
| Lower BMI via GLP-1R activation | Depressive symptoms (seen a general practitioner) | Women | 3 | MR Egger | 0.86 (0.47 to 1.59) | 0.64 | 0.84 | |
| Lower BMI via GLP-1R activation | Depressive symptoms (seen a general practitioner) | Women | 3 | MR-RAPS | 0.81 (0.65 to 1.00) | 0.05 | | |
| Lower BMI via GLP-1R activation | Depressive symptoms (seen a general practitioner) | Men | 3 | IVW | 0.94 (0.81 to 1.11) | 0.48 | | 0.21 |
| Lower BMI via GLP-1R activation | Depressive symptoms (seen a general practitioner) | Men | 3 | Weighted median | 0.91 (0.74 to 1.13) | 0.40 | | |
| Lower BMI via GLP-1R activation | Depressive symptoms (seen a general practitioner) | Men | 3 | MR Egger | 0.61 (0.16 to 2.30) | 0.47 | 0.51 | |
| Lower BMI via GLP-1R activation | Depressive symptoms (seen a general practitioner) | Men | 3 | MR-RAPS | 0.94 (0.79 to 1.12) | 0.47 | | |
| Lower BMI via GLP-1R activation | Depressive symptoms (seen a psychiatrist) | Women | 3 | IVW | 0.78 (0.60 to 1.02) | 0.07 | | 0.03 |
| Lower BMI via GLP-1R activation | Depressive symptoms (seen a psychiatrist) | Women | 3 | Weighted median | 0.81 (0.55 to 1.17) | 0.26 | | |
| Lower BMI via GLP-1R activation | Depressive symptoms (seen a psychiatrist) | Women | 3 | MR Egger | 2.16 (0.66 to 7.00) | 0.20 | 0.08 | |
| Lower BMI via GLP-1R activation | Depressive symptoms (seen a psychiatrist) | Women | 3 | MR-RAPS | 0.70 (0.52 to 0.96) | 0.03 | | |

| | | | | | | | | |
|---|---|---|---|---|---|---|---|---|
| Lower BMI via GLP-1R activation | Depressive symptoms (seen a psychiatrist) | Men | 3 | IVW | 1.04 (0.83 to 1.31) | 0.73 | | 0.15 |
| Lower BMI via GLP-1R activation | Depressive symptoms (seen a psychiatrist) | Men | 3 | Weighted median | 0.97 (0.71 to 1.31) | 0.84 | | |
| Lower BMI via GLP-1R activation | Depressive symptoms (seen a psychiatrist) | Men | 3 | MR Egger | 0.57 (0.06 to 5.32) | 0.62 | 0.59 | |
| Lower BMI via GLP-1R activation | Depressive symptoms (seen a psychiatrist) | Men | 3 | MR-RAPS | 1.02 (0.80 to 1.31) | 0.86 | | |
| Lower BMI | Depressive symptoms (seen a general practitioner) | Women | 300 | IVW | 0.97 (0.95 to 0.98) | <0.001 | | <0.001 |
| Lower BMI | Depressive symptoms (seen a general practitioner) | Women | 300 | Weighted median | 0.98 (0.96 to 0.99) | 0.01 | | |
| Lower BMI | Depressive symptoms (seen a general practitioner) | Women | 300 | MR Egger | 0.99 (0.96 to 1.03) | 0.75 | 0.10 | |
| Lower BMI | Depressive symptoms (seen a general practitioner) | Women | 300 | MR-RAPS | 0.97 (0.96 to 0.98) | <0.001 | | |
| Lower BMI | Depressive symptoms (seen a general practitioner) | Men | 251 | IVW | 1.01 (0.99 to 1.02) | 0.34 | | <0.001 |
| Lower BMI | Depressive symptoms (seen a general practitioner) | Men | 251 | Weighted median | 1.01 (0.99 to 1.03) | 0.37 | | |
| Lower BMI | Depressive symptoms (seen a general practitioner) | Men | 251 | MR Egger | 1.00 (0.96 to 1.04) | 0.998 | 0.70 | |
| Lower BMI | Depressive symptoms (seen a general practitioner) | Men | 251 | MR-RAPS | 1.01 (1.00 to 1.02) | 0.11 | | |
| Lower BMI | Depressive symptoms (seen a psychiatrist) | Women | 300 | IVW | 0.98 (0.96 to 0.99) | 0.003 | | <0.001 |
| Lower BMI | Depressive symptoms (seen a psychiatrist) | Women | 300 | Weighted median | 0.99 (0.97 to 1.01) | 0.25 | | |

| | | | | | | | | |
|---|---|---|---|---|---|---|---|---|
| Lower BMI | Depressive symptoms (seen a psychiatrist) | Women | 300 | MR Egger | 0.99 (0.95 to 1.04) | 0.75 | 0.43 | |
| Lower BMI | Depressive symptoms (seen a psychiatrist) | Women | 300 | MR-RAPS | 0.98 (0.96 to 0.99) | <0.001 | | |
| Lower BMI | Depressive symptoms (seen a psychiatrist) | Men | 251 | IVW | 1.00 (0.98 to 1.02) | 0.81 | | <0.001 |
| Lower BMI | Depressive symptoms (seen a psychiatrist) | Men | 251 | Weighted median | 1.01 (0.98 to 1.03) | 0.61 | | |
| Lower BMI | Depressive symptoms (seen a psychiatrist) | Men | 251 | MR Egger | 1.01 (0.95 to 1.06) | 0.85 | 0.76 | |
| Lower BMI | Depressive symptoms (seen a psychiatrist) | Men | 251 | MR-RAPS | 1.00 (0.98 to 1.02) | 0.94 | | |
| Lower HbA1c via GLP-1R activation | Depressive symptoms (seen a general practitioner) | Women | 2 | IVW | 1.07 (0.97 to 1.20) | 0.19 | | 0.61 |
| Lower HbA1c via GLP-1R activation | Depressive symptoms (seen a general practitioner) | Men | 2 | IVW | 1.03 (0.90 to 1.18) | 0.68 | | 0.76 |
| Lower HbA1c via GLP-1R activation | Depressive symptoms (seen a psychiatrist) | Women | 2 | IVW | 1.00 (0.86 to 1.18) | 0.97 | | 0.87 |
| Lower HbA1c via GLP-1R activation | Depressive symptoms (seen a psychiatrist) | Men | 2 | IVW | 0.99 (0.81 to 1.20) | 0.92 | | 0.54 |
| Lower HbA1c | Depressive symptoms (seen a general practitioner) | Women | 193 | IVW | 1.00 (0.99 to 1.00) | 0.53 | | <0.001 |
| Lower HbA1c | Depressive symptoms (seen a general practitioner) | Women | 193 | Weighted median | 1.00 (0.99 to 1.01) | 0.68 | | |
| Lower HbA1c | Depressive symptoms (seen a general practitioner) | Women | 193 | MR Egger | 1.00 (0.99 to 1.01) | 0.85 | 0.49 | |
| Lower HbA1c | Depressive symptoms (seen a general practitioner) | Women | 193 | MR-RAPS | 1.00 (0.99 to 1.00) | 0.43 | | |

| | | | | | | | | |
|---|---|---|---|---|---|---|---|---|
| Lower HbA1c | Depressive symptoms (seen a general practitioner) | Men | 168 | IVW | 1.00 (0.99 to 1.01) | 0.92 | | 0.01 |
| Lower HbA1c | Depressive symptoms (seen a general practitioner) | Men | 168 | Weighted median | 1.00 (0.99 to 1.01) | 0.50 | | |
| Lower HbA1c | Depressive symptoms (seen a general practitioner) | Men | 168 | MR Egger | 1.00 (0.99 to 1.01) | 0.90 | 0.93 | |
| Lower HbA1c | Depressive symptoms (seen a general practitioner) | Men | 168 | MR-RAPS | 1.00 (1.00 to 1.01) | 0.60 | | |
| Lower HbA1c | Depressive symptoms (seen a psychiatrist) | Women | 193 | IVW | 1.00 (0.99 to 1.00) | 0.38 | | 0.003 |
| Lower HbA1c | Depressive symptoms (seen a psychiatrist) | Women | 193 | Weighted median | 0.99 (0.98 to 1.00) | 0.21 | | |
| Lower HbA1c | Depressive symptoms (seen a psychiatrist) | Women | 193 | MR Egger | 1.00 (0.99 to 1.01) | 0.90 | 0.43 | |
| Lower HbA1c | Depressive symptoms (seen a psychiatrist) | Women | 193 | MR-RAPS | 1.00 (0.99 to 1.00) | 0.15 | | |
| Lower HbA1c | Depressive symptoms (seen a psychiatrist) | Men | 168 | IVW | 1.00 (0.99 to 1.01) | 0.90 | | 0.11 |
| Lower HbA1c | Depressive symptoms (seen a psychiatrist) | Men | 168 | Weighted median | 1.00 (0.98 to 1.01) | 0.45 | | |
| Lower HbA1c | Depressive symptoms (seen a psychiatrist) | Men | 168 | MR Egger | 1.00 (0.98 to 1.01) | 0.61 | 0.49 | |
| Lower HbA1c | Depressive symptoms (seen a psychiatrist) | Men | 168 | MR-RAPS | 1.00 (0.99 to 1.01) | 0.85 | | |

BMI, body mass index; GLP-1R, glucagon-like peptide-1 receptor; HbA1c, glycated hemoglobin; IVW, inverse variance weighting; MR, Mendelian randomization; RAPS, robust adjusted profile score. Estimates are presented as odds ratio per 1-kg/$m^2$ decrease in BMI or per 1-mmol/mol decrease in HbA1c.

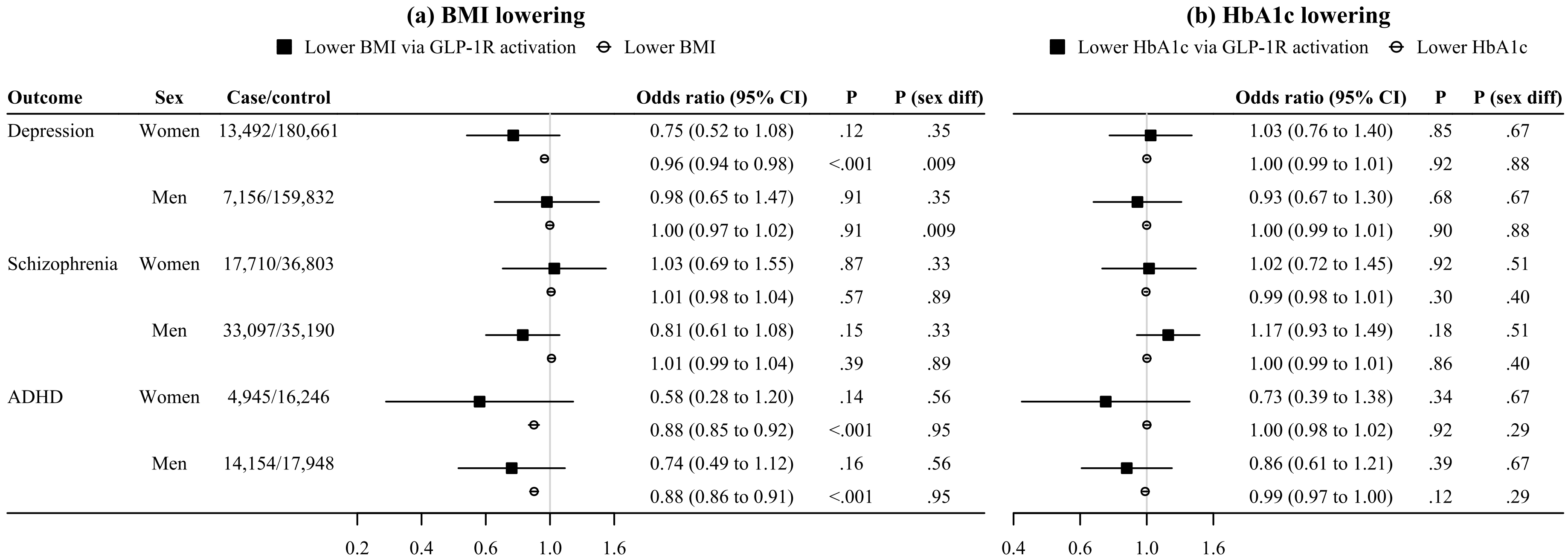

| Outcome | Sex | Case/control | (a) Odds ratio (95% CI) | (a) P | (a) P (sex diff) | (b) Odds ratio (95% CI) | (b) P | (b) P (sex diff) |
|---|---|---|---|---|---|---|---|---|
| Depression | Women | 13,492/180,661 | 0.75 (0.52 to 1.08) | .12 | .35 | 1.03 (0.76 to 1.40) | .85 | .67 |
| | | | 0.96 (0.94 to 0.98) | <.001 | .009 | 1.00 (0.99 to 1.01) | .92 | .88 |
| | Men | 7,156/159,832 | 0.98 (0.65 to 1.47) | .91 | .35 | 0.93 (0.67 to 1.30) | .68 | .67 |
| | | | 1.00 (0.97 to 1.02) | .91 | .009 | 1.00 (0.99 to 1.01) | .90 | .88 |
| Schizophrenia | Women | 17,710/36,803 | 1.03 (0.69 to 1.55) | .87 | .33 | 1.02 (0.72 to 1.45) | .92 | .51 |
| | | | 1.01 (0.98 to 1.04) | .57 | .89 | 0.99 (0.98 to 1.01) | .30 | .40 |
| | Men | 33,097/35,190 | 0.81 (0.61 to 1.08) | .15 | .33 | 1.17 (0.93 to 1.49) | .18 | .51 |
| | | | 1.01 (0.99 to 1.04) | .39 | .89 | 1.00 (0.99 to 1.01) | .86 | .40 |
| ADHD | Women | 4,945/16,246 | 0.58 (0.28 to 1.20) | .14 | .56 | 0.73 (0.39 to 1.38) | .34 | .67 |
| | | | 0.88 (0.85 to 0.92) | <.001 | .95 | 1.00 (0.98 to 1.02) | .92 | .29 |
| | Men | 14,154/17,948 | 0.74 (0.49 to 1.12) | .16 | .56 | 0.86 (0.61 to 1.21) | .39 | .67 |
| | | | 0.88 (0.86 to 0.91) | <.001 | .95 | 0.99 (0.97 to 1.00) | .12 | .29 |

**eFigure 3. IVW MR estimates for sex-specific associations of GLP-1R activation with the risk of depression, schizophrenia, and ADHD in comparison with those for lower BMI and lower HbA1c.**

ADHD, attention deficit hyperactivity disorder; BMI, body mass index; GLP-1R, glucagon-like peptide-1 receptor; HbA1c, glycated hemoglobin; IVW, inverse variance weighted; MR, Mendelian randomization. Black squares denote genetically predicted (a) lower BMI or (b) lower HbA1c via GLP-1R activation based on *GLP1R* variants. White circles denote genetically predicted (a) lower BMI or (b) lower HbA1c based on genome-wide variants. Estimates are presented as odds ratio per 1-kg/$m^2$ decrease in BMI or per 1-mmol/mol decrease in HbA1c.

**eTable 8. Sensitivity MR analyses for the associations of GLP-1R activation, lower BMI, and lower HbA1c with the risk of mental health disorders.**

| Exposure | Outcome | Sex | SNPs | Method | Odds ratio (95% CI) | *P* value | *P* value (intercept) | *P* value (Q) |
|---|---|---|---|---|---|---|---|---|
| Lower BMI via GLP-1R activation | Depression | Overall | 3 | IVW | 0.88 (0.79 to 0.98) | 0.02 | | 0.05 |
| Lower BMI via GLP-1R activation | Depression | Overall | 3 | Weighted median | 0.88 (0.76 to 1.01) | 0.07 | | |
| Lower BMI via GLP-1R activation | Depression | Overall | 3 | MR Egger | 0.82 (0.20 to 3.36) | 0.78 | 0.91 | |
| Lower BMI via GLP-1R activation | Depression | Overall | 3 | MR-RAPS | 0.87 (0.78 to 0.98) | 0.02 | | |
| Lower BMI via GLP-1R activation | Depression | Women | 3 | IVW | 0.67 (0.47 to 0.94) | 0.02 | | 0.11 |
| Lower BMI via GLP-1R activation | Depression | Women | 3 | Weighted median | 0.75 (0.42 to 1.33) | 0.32 | | |
| Lower BMI via GLP-1R activation | Depression | Women | 3 | MR Egger | 2.14 (0.67 to 6.86) | 0.20 | 0.04 | |
| Lower BMI via GLP-1R activation | Depression | Women | 3 | MR-RAPS | 0.67 (0.45 to 0.99) | 0.04 | | |
| Lower BMI via GLP-1R activation | Depression | Men | 3 | IVW | 1.02 (0.72 to 1.44) | 0.90 | | 0.64 |
| Lower BMI via GLP-1R activation | Depression | Men | 3 | Weighted median | 1.13 (0.74 to 1.72) | 0.57 | | |
| Lower BMI via GLP-1R activation | Depression | Men | 3 | MR Egger | 2.14 (0.31 to 14.80) | 0.44 | 0.45 | |
| Lower BMI via GLP-1R activation | Depression | Men | 3 | MR-RAPS | 1.02 (0.70 to 1.49) | 0.91 | | |
| Lower BMI via GLP-1R activation | Post-partum depression | Women | 3 | IVW | 0.64 (0.45 to 0.91) | 0.01 | | 0.76 |
| Lower BMI via GLP-1R activation | Post-partum depression | Women | 3 | Weighted median | 0.63 (0.40 to 1.00) | 0.05 | | |
| Lower BMI via GLP-1R activation | Post-partum depression | Women | 3 | MR Egger | 0.48 (0.14 to 1.57) | 0.22 | 0.60 | |
| Lower BMI via GLP-1R activation | Post-partum depression | Women | 3 | MR-RAPS | 0.64 (0.42 to 0.98) | 0.04 | | |
| Lower BMI via GLP-1R activation | Bipolar disorder | Overall | 3 | IVW | 0.70 (0.55 to 0.88) | 0.003 | | 0.06 |
| Lower BMI via GLP-1R activation | Bipolar disorder | Overall | 3 | Weighted median | 0.64 (0.48 to 0.87) | 0.01 | | |
| Lower BMI via GLP-1R activation | Bipolar disorder | Overall | 3 | MR Egger | 0.18 (0.04 to 0.79) | 0.02 | 0.07 | |
| Lower BMI via GLP-1R activation | Bipolar disorder | Overall | 3 | MR-RAPS | 0.66 (0.51 to 0.86) | 0.002 | | |
| Lower BMI via GLP-1R activation | Bipolar disorder I | Overall | 3 | IVW | 0.65 (0.49 to 0.86) | 0.003 | | 0.27 |
| Lower BMI via GLP-1R activation | Bipolar disorder I | Overall | 3 | Weighted median | 0.63 (0.44 to 0.91) | 0.01 | | |
| Lower BMI via GLP-1R activation | Bipolar disorder I | Overall | 3 | MR Egger | 0.19 (0.04 to 0.91) | 0.04 | 0.12 | |
| Lower BMI via GLP-1R activation | Bipolar disorder I | Overall | 3 | MR-RAPS | 0.64 (0.47 to 0.88) | 0.01 | | |

| | | | | | | | | |
|---|---|---|---|---|---|---|---|---|
| Lower BMI via GLP-1R activation | Bipolar disorder II | Overall | 3 | IVW | 0.72 (0.44 to 1.17) | 0.18 | | 0.12 |
| Lower BMI via GLP-1R activation | Bipolar disorder II | Overall | 3 | Weighted median | 0.65 (0.35 to 1.23) | 0.19 | | |
| Lower BMI via GLP-1R activation | Bipolar disorder II | Overall | 3 | MR Egger | 0.32 (0.002 to 66.81) | 0.68 | 0.76 | |
| Lower BMI via GLP-1R activation | Bipolar disorder II | Overall | 3 | MR-RAPS | 0.68 (0.40 to 1.13) | 0.14 | | |
| Lower BMI via GLP-1R activation | PTSD | Overall | 3 | IVW | 0.69 (0.48 to 0.99) | 0.04 | | 0.91 |
| Lower BMI via GLP-1R activation | PTSD | Overall | 3 | Weighted median | 0.69 (0.45 to 1.05) | 0.09 | | |
| Lower BMI via GLP-1R activation | PTSD | Overall | 3 | MR Egger | 1.05 (0.14 to 7.70) | 0.96 | 0.67 | |
| Lower BMI via GLP-1R activation | PTSD | Overall | 3 | MR-RAPS | 0.69 (0.46 to 1.02) | 0.06 | | |
| Lower BMI via GLP-1R activation | Schizophrenia | Overall | 3 | IVW | 0.93 (0.76 to 1.15) | 0.52 | | 0.18 |
| Lower BMI via GLP-1R activation | Schizophrenia | Overall | 3 | Weighted median | 0.97 (0.74 to 1.27) | 0.82 | | |
| Lower BMI via GLP-1R activation | Schizophrenia | Overall | 3 | MR Egger | 1.13 (0.14 to 9.28) | 0.91 | 0.86 | |
| Lower BMI via GLP-1R activation | Schizophrenia | Overall | 3 | MR-RAPS | 0.94 (0.75 to 1.17) | 0.57 | | |
| Lower BMI via GLP-1R activation | Schizophrenia | Women | 3 | IVW | 1.02 (0.70 to 1.49) | 0.93 | | 0.51 |
| Lower BMI via GLP-1R activation | Schizophrenia | Women | 3 | Weighted median | 0.88 (0.55 to 1.41) | 0.59 | | |
| Lower BMI via GLP-1R activation | Schizophrenia | Women | 3 | MR Egger | 1.40 (0.36 to 5.39) | 0.63 | 0.63 | |
| Lower BMI via GLP-1R activation | Schizophrenia | Women | 3 | MR-RAPS | 1.02 (0.66 to 1.57) | 0.93 | | |
| Lower BMI via GLP-1R activation | Schizophrenia | Men | 3 | IVW | 0.93 (0.73 to 1.18) | 0.57 | | 0.10 |
| Lower BMI via GLP-1R activation | Schizophrenia | Men | 3 | Weighted median | 0.95 (0.67 to 1.33) | 0.75 | | |
| Lower BMI via GLP-1R activation | Schizophrenia | Men | 3 | MR Egger | 1.69 (0.12 to 24.58) | 0.70 | 0.66 | |
| Lower BMI via GLP-1R activation | Schizophrenia | Men | 3 | MR-RAPS | 0.93 (0.72 to 1.21) | 0.60 | | |
| Lower BMI via GLP-1R activation | Anorexia nervosa | Overall | 3 | IVW | 1.00 (0.71 to 1.39) | 0.99 | | 0.84 |
| Lower BMI via GLP-1R activation | Anorexia nervosa | Overall | 3 | Weighted median | 0.98 (0.66 to 1.43) | 0.90 | | |
| Lower BMI via GLP-1R activation | Anorexia nervosa | Overall | 3 | MR Egger | 0.84 (0.13 to 5.37) | 0.85 | 0.85 | |
| Lower BMI via GLP-1R activation | Anorexia nervosa | Overall | 3 | MR-RAPS | 1.00 (0.70 to 1.43) | 0.99 | | |
| Lower BMI via GLP-1R activation | ADHD | Overall | 3 | IVW | 0.78 (0.61 to 0.98) | 0.03 | | 0.04 |
| Lower BMI via GLP-1R activation | ADHD | Overall | 3 | Weighted median | 0.86 (0.62 to 1.20) | 0.39 | | |
| Lower BMI via GLP-1R activation | ADHD | Overall | 3 | MR Egger | 1.33 (0.06 to 30.98) | 0.86 | 0.73 | |

| | | | | | | | | |
|---|---|---|---|---|---|---|---|---|
| Lower BMI via GLP-1R activation | ADHD | Overall | 3 | MR-RAPS | 0.83 (0.65 to 1.05) | 0.12 | | |
| Lower BMI via GLP-1R activation | ADHD | Women | 3 | IVW | 0.68 (0.34 to 1.36) | 0.28 | | 0.38 |
| Lower BMI via GLP-1R activation | ADHD | Women | 3 | Weighted median | 0.58 (0.25 to 1.35) | 0.21 | | |
| Lower BMI via GLP-1R activation | ADHD | Women | 3 | MR Egger | 0.14 (0.01 to 1.50) | 0.11 | 0.17 | |
| Lower BMI via GLP-1R activation | ADHD | Women | 3 | MR-RAPS | 0.67 (0.30 to 1.47) | 0.31 | | |
| Lower BMI via GLP-1R activation | ADHD | Men | 3 | IVW | 0.85 (0.60 to 1.20) | 0.36 | | 0.03 |
| Lower BMI via GLP-1R activation | ADHD | Men | 3 | Weighted median | 1.15 (0.71 to 1.86) | 0.57 | | |
| Lower BMI via GLP-1R activation | ADHD | Men | 3 | MR Egger | 7.28 (0.32 to 166.88) | 0.21 | 0.17 | |
| Lower BMI via GLP-1R activation | ADHD | Men | 3 | MR-RAPS | 0.96 (0.67 to 1.37) | 0.81 | | |
| Lower BMI via GLP-1R activation | Autism spectrum disorder | Overall | 3 | IVW | 0.82 (0.58 to 1.15) | 0.24 | | 0.23 |
| Lower BMI via GLP-1R activation | Autism spectrum disorder | Overall | 3 | Weighted median | 0.69 (0.45 to 1.06) | 0.09 | | |
| Lower BMI via GLP-1R activation | Autism spectrum disorder | Overall | 3 | MR Egger | 0.26 (0.02 to 2.90) | 0.28 | 0.35 | |
| Lower BMI via GLP-1R activation | Autism spectrum disorder | Overall | 3 | MR-RAPS | 0.79 (0.55 to 1.15) | 0.22 | | |
| Lower BMI via GLP-1R activation | Tourette syndrome | Overall | 3 | IVW | 1.08 (0.58 to 2.03) | 0.81 | | 0.56 |
| Lower BMI via GLP-1R activation | Tourette syndrome | Overall | 3 | Weighted median | 1.13 (0.54 to 2.38) | 0.74 | | |
| Lower BMI via GLP-1R activation | Tourette syndrome | Overall | 3 | MR Egger | 6.57 (0.20 to 214.68) | 0.29 | 0.30 | |
| Lower BMI via GLP-1R activation | Tourette syndrome | Overall | 3 | MR-RAPS | 1.08 (0.55 to 2.13) | 0.82 | | |
| Lower BMI | Depression | Overall | 502 | IVW | 0.98 (0.97 to 0.99) | <0.001 | | <0.001 |
| Lower BMI | Depression | Overall | 502 | Weighted median | 0.98 (0.97 to 0.99) | <0.001 | | |
| Lower BMI | Depression | Overall | 502 | MR Egger | 0.99 (0.97 to 1.02) | 0.55 | 0.24 | |
| Lower BMI | Depression | Overall | 502 | MR-RAPS | 0.98 (0.97 to 0.98) | <0.001 | | |
| Lower BMI | Depression | Women | 300 | IVW | 0.96 (0.94 to 0.98) | <0.001 | | 0.04 |
| Lower BMI | Depression | Women | 300 | Weighted median | 0.95 (0.92 to 0.98) | <0.001 | | |
| Lower BMI | Depression | Women | 300 | MR Egger | 0.96 (0.91 to 1.01) | 0.12 | 0.94 | |
| Lower BMI | Depression | Women | 300 | MR-RAPS | 0.96 (0.94 to 0.97) | <0.001 | | |
| Lower BMI | Depression | Men | 251 | IVW | 1.00 (0.97 to 1.02) | 0.91 | | 0.04 |

| | | | | | | | | |
|---|---|---|---|---|---|---|---|---|
| Lower BMI | Depression | Men | 251 | Weighted median | 1.01 (0.97 to 1.05) | 0.67 | | |
| Lower BMI | Depression | Men | 251 | MR Egger | 1.05 (0.98 to 1.12) | 0.21 | 0.16 | |
| Lower BMI | Depression | Men | 251 | MR-RAPS | 1.00 (0.98 to 1.02) | 0.98 | | |
| Lower BMI | Post-partum depression | Women | 300 | IVW | 0.98 (0.96 to 1.00) | 0.03 | | <0.001 |
| Lower BMI | Post-partum depression | Women | 300 | Weighted median | 0.99 (0.96 to 1.02) | 0.45 | | |
| Lower BMI | Post-partum depression | Women | 300 | MR Egger | 0.98 (0.93 to 1.04) | 0.57 | 0.81 | |
| Lower BMI | Post-partum depression | Women | 300 | MR-RAPS | 0.98 (0.96 to 0.99) | 0.01 | | |
| Lower BMI | Bipolar disorder | Overall | 506 | IVW | 1.00 (0.98 to 1.02) | 0.77 | | <0.001 |
| Lower BMI | Bipolar disorder | Overall | 506 | Weighted median | 1.00 (0.98 to 1.03) | 0.87 | | |
| Lower BMI | Bipolar disorder | Overall | 506 | MR Egger | 0.97 (0.92 to 1.02) | 0.27 | 0.19 | |
| Lower BMI | Bipolar disorder | Overall | 506 | MR-RAPS | 1.00 (0.99 to 1.01) | 0.88 | | |
| Lower BMI | Bipolar disorder I | Overall | 499 | IVW | 1.01 (0.98 to 1.03) | 0.57 | | <0.001 |
| Lower BMI | Bipolar disorder I | Overall | 499 | Weighted median | 1.00 (0.97 to 1.03) | 0.93 | | |
| Lower BMI | Bipolar disorder I | Overall | 499 | MR Egger | 0.96 (0.90 to 1.02) | 0.20 | 0.11 | |
| Lower BMI | Bipolar disorder I | Overall | 499 | MR-RAPS | 1.00 (0.98 to 1.01) | 0.95 | | |
| Lower BMI | Bipolar disorder II | Overall | 499 | IVW | 1.00 (0.97 to 1.03) | 0.94 | | <0.001 |
| Lower BMI | Bipolar disorder II | Overall | 499 | Weighted median | 0.99 (0.95 to 1.04) | 0.82 | | |
| Lower BMI | Bipolar disorder II | Overall | 499 | MR Egger | 0.98 (0.90 to 1.05) | 0.52 | 0.51 | |
| Lower BMI | Bipolar disorder II | Overall | 499 | MR-RAPS | 1.00 (0.97 to 1.02) | 0.79 | | |
| Lower BMI | PTSD | Overall | 506 | IVW | 0.94 (0.92 to 0.96) | <0.001 | | <0.001 |
| Lower BMI | PTSD | Overall | 506 | Weighted median | 0.94 (0.91 to 0.97) | <0.001 | | |
| Lower BMI | PTSD | Overall | 506 | MR Egger | 0.96 (0.91 to 1.01) | 0.13 | 0.36 | |
| Lower BMI | PTSD | Overall | 506 | MR-RAPS | 0.94 (0.92 to 0.95) | <0.001 | | |
| Lower BMI | Schizophrenia | Overall | 505 | IVW | 1.03 (1.01 to 1.05) | 0.01 | | <0.001 |
| Lower BMI | Schizophrenia | Overall | 505 | Weighted median | 1.00 (0.98 to 1.03) | 0.72 | | |
| Lower BMI | Schizophrenia | Overall | 505 | MR Egger | 0.93 (0.88 to 0.99) | 0.02 | <0.001 | |
| Lower BMI | Schizophrenia | Overall | 505 | MR-RAPS | 1.02 (1.01 to 1.03) | <0.001 | | |

| | | | | | | | | |
|---|---|---|---|---|---|---|---|---|
| Lower BMI | Schizophrenia | Overall | 498 | IVW (Steiger filtering) | 1.03 (1.01 to 1.05) | 0.004 | | <0.001 |
| Lower BMI | Schizophrenia | Overall | 498 | Weighted median (Steiger filtering) | 1.00 (0.98 to 1.03) | 0.72 | | |
| Lower BMI | Schizophrenia | Overall | 498 | MR Egger (Steiger filtering) | 0.95 (0.90 to 1.01) | 0.09 | 0.003 | |
| Lower BMI | Schizophrenia | Overall | 498 | MR-RAPS (Steiger filtering) | 1.02 (1.01 to 1.03) | 0.001 | | |
| Lower BMI | Schizophrenia | Women | 300 | IVW | 1.01 (0.98 to 1.04) | 0.57 | | <0.001 |
| Lower BMI | Schizophrenia | Women | 300 | Weighted median | 0.98 (0.94 to 1.02) | 0.25 | | |
| Lower BMI | Schizophrenia | Women | 300 | MR Egger | 0.90 (0.83 to 0.98) | 0.01 | 0.003 | |
| Lower BMI | Schizophrenia | Women | 300 | MR-RAPS | 1.01 (0.99 to 1.03) | 0.22 | | |
| Lower BMI | Schizophrenia | Women | 297 | IVW (Steiger filtering) | 1.01 (0.98 to 1.04) | 0.56 | | <0.001 |
| Lower BMI | Schizophrenia | Women | 297 | Weighted median (Steiger filtering) | 0.98 (0.94 to 1.02) | 0.26 | | |
| Lower BMI | Schizophrenia | Women | 297 | MR Egger (Steiger filtering) | 0.92 (0.85 to 0.99) | 0.03 | 0.009 | |
| Lower BMI | Schizophrenia | Women | 297 | MR-RAPS (Steiger filtering) | 1.01 (0.99 to 1.03) | 0.25 | | |
| Lower BMI | Schizophrenia | Men | 252 | IVW | 1.01 (0.99 to 1.04) | 0.39 | | <0.001 |
| Lower BMI | Schizophrenia | Men | 252 | Weighted median | 0.99 (0.96 to 1.02) | 0.69 | | |
| Lower BMI | Schizophrenia | Men | 252 | MR Egger | 0.97 (0.90 to 1.04) | 0.32 | 0.16 | |
| Lower BMI | Schizophrenia | Men | 252 | MR-RAPS | 1.01 (0.99 to 1.02) | 0.44 | | |
| Lower BMI | Schizophrenia | Men | 251 | IVW (Steiger filtering) | 1.01 (0.98 to 1.03) | 0.50 | | <0.001 |
| Lower BMI | Schizophrenia | Men | 251 | Weighted median (Steiger filtering) | 0.99 (0.96 to 1.02) | 0.69 | | |

| | | | | | | | | |
|---|---|---|---|---|---|---|---|---|
| Lower BMI | Schizophrenia | Men | 251 | MR Egger (Steiger filtering) | 0.97 (0.91 to 1.04) | 0.40 | 0.24 | |
| Lower BMI | Schizophrenia | Men | 251 | MR-RAPS (Steiger filtering) | 1.01 (0.99 to 1.02) | 0.50 | | |
| Lower BMI | Anorexia nervosa | Overall | 449 | IVW | 1.10 (1.07 to 1.12) | <0.001 | | <0.001 |
| Lower BMI | Anorexia nervosa | Overall | 449 | Weighted median | 1.07 (1.03 to 1.11) | <0.001 | | |
| Lower BMI | Anorexia nervosa | Overall | 449 | MR Egger | 1.02 (0.96 to 1.09) | 0.54 | 0.03 | |
| Lower BMI | Anorexia nervosa | Overall | 449 | MR-RAPS | 1.10 (1.08 to 1.12) | <0.001 | | |
| Lower BMI | Anorexia nervosa | Overall | 448 | IVW (Steiger filtering) | 1.09 (1.06 to 1.12) | <0.001 | | <0.001 |
| Lower BMI | Anorexia nervosa | Overall | 448 | Weighted median (Steiger filtering) | 1.07 (1.03 to 1.10) | <0.001 | | |
| Lower BMI | Anorexia nervosa | Overall | 448 | MR Egger (Steiger filtering) | 1.02 (0.95 to 1.09) | 0.60 | 0.02 | |
| Lower BMI | Anorexia nervosa | Overall | 448 | MR-RAPS (Steiger filtering) | 1.10 (1.08 to 1.12) | <0.001 | | |
| Lower BMI | ADHD | Overall | 496 | IVW | 0.89 (0.87 to 0.90) | <0.001 | | <0.001 |
| Lower BMI | ADHD | Overall | 496 | Weighted median | 0.90 (0.88 to 0.92) | <0.001 | | |
| Lower BMI | ADHD | Overall | 496 | MR Egger | 0.95 (0.91 to 1.00) | 0.04 | 0.001 | |
| Lower BMI | ADHD | Overall | 496 | MR-RAPS | 0.89 (0.88 to 0.90) | <0.001 | | |
| Lower BMI | ADHD | Women | 257 | IVW | 0.88 (0.85 to 0.92) | <0.001 | | 0.03 |
| Lower BMI | ADHD | Women | 257 | Weighted median | 0.91 (0.85 to 0.96) | 0.001 | | |
| Lower BMI | ADHD | Women | 257 | MR Egger | 0.92 (0.81 to 1.04) | 0.20 | 0.49 | |
| Lower BMI | ADHD | Women | 257 | MR-RAPS | 0.89 (0.86 to 0.92) | <0.001 | | |
| Lower BMI | ADHD | Women | 256 | IVW (Steiger filtering) | 0.89 (0.85 to 0.92) | <0.001 | | 0.12 |
| Lower BMI | ADHD | Women | 256 | Weighted median (Steiger filtering) | 0.91 (0.85 to 0.96) | 0.001 | | |

| | | | | | | | | |
|---|---|---|---|---|---|---|---|---|
| Lower BMI | ADHD | Women | 256 | MR Egger (Steiger filtering) | 0.92 (0.81 to 1.04) | 0.16 | 0.58 | |
| Lower BMI | ADHD | Women | 256 | MR-RAPS (Steiger filtering) | 0.89 (0.86 to 0.93) | <0.001 | | |
| Lower BMI | ADHD | Men | 215 | IVW | 0.88 (0.86 to 0.91) | <0.001 | | <0.001 |
| Lower BMI | ADHD | Men | 215 | Weighted median | 0.85 (0.82 to 0.89) | <0.001 | | |
| Lower BMI | ADHD | Men | 215 | MR Egger | 0.89 (0.81 to 0.99) | 0.03 | 0.84 | |
| Lower BMI | ADHD | Men | 215 | MR-RAPS | 0.88 (0.85 to 0.90) | <0.001 | | |
| Lower BMI | Autism spectrum disorder | Overall | 453 | IVW | 0.98 (0.95 to 1.00) | 0.05 | | <0.001 |
| Lower BMI | Autism spectrum disorder | Overall | 453 | Weighted median | 0.98 (0.94 to 1.01) | 0.17 | | |
| Lower BMI | Autism spectrum disorder | Overall | 453 | MR Egger | 0.98 (0.92 to 1.04) | 0.51 | 0.90 | |
| Lower BMI | Autism spectrum disorder | Overall | 453 | MR-RAPS | 0.97 (0.95 to 0.99) | 0.002 | | |
| Lower BMI | Autism spectrum disorder | Overall | 450 | IVW (Steiger filtering) | 0.97 (0.95 to 0.99) | 0.01 | | <0.001 |
| Lower BMI | Autism spectrum disorder | Overall | 450 | Weighted median (Steiger filtering) | 0.97 (0.94 to 1.01) | 0.11 | | |
| Lower BMI | Autism spectrum disorder | Overall | 450 | MR Egger (Steiger filtering) | 0.98 (0.93 to 1.04) | 0.54 | 0.68 | |
| Lower BMI | Autism spectrum disorder | Overall | 450 | MR-RAPS (Steiger filtering) | 0.97 (0.95 to 0.99) | <0.001 | | |
| Lower BMI | Tourette syndrome | Overall | 451 | IVW | 0.97 (0.93 to 1.01) | 0.14 | | <0.001 |
| Lower BMI | Tourette syndrome | Overall | 451 | Weighted median | 0.96 (0.90 to 1.03) | 0.22 | | |
| Lower BMI | Tourette syndrome | Overall | 451 | MR Egger | 1.03 (0.93 to 1.15) | 0.56 | 0.21 | |
| Lower BMI | Tourette syndrome | Overall | 451 | MR-RAPS | 0.97 (0.93 to 1.00) | 0.09 | | |
| Lower BMI | Tourette syndrome | Overall | 444 | IVW (Steiger filtering) | 0.98 (0.94 to 1.02) | 0.28 | | <0.001 |
| Lower BMI | Tourette syndrome | Overall | 444 | Weighted median (Steiger filtering) | 0.96 (0.90 to 1.03) | 0.24 | | |

| | | | | | | | | |
|---|---|---|---|---|---|---|---|---|
| Lower BMI | Tourette syndrome | Overall | 444 | MR Egger (Steiger filtering) | 1.03 (0.93 to 1.14) | 0.63 | 0.33 | |
| Lower BMI | Tourette syndrome | Overall | 444 | MR-RAPS (Steiger filtering) | 0.97 (0.94 to 1.01) | 0.14 | | |
| Lower HbA1c via GLP-1R activation | Depression | Overall | 2 | IVW | 1.03 (0.96 to 1.11) | 0.45 | | 0.97 |
| Lower HbA1c via GLP-1R activation | Depression | Women | 2 | IVW | 1.13 (0.92 to 1.40) | 0.25 | | 0.40 |
| Lower HbA1c via GLP-1R activation | Depression | Men | 2 | IVW | 0.94 (0.70 to 1.26) | 0.69 | | 0.90 |
| Lower HbA1c via GLP-1R activation | Post-partum depression | Women | 2 | IVW | 0.90 (0.73 to 1.11) | 0.32 | | 0.51 |
| Lower HbA1c via GLP-1R activation | Bipolar disorder | Overall | 2 | IVW | 1.10 (0.93 to 1.29) | 0.26 | | 0.20 |
| Lower HbA1c via GLP-1R activation | Bipolar disorder I | Overall | 2 | IVW | 1.14 (0.93 to 1.40) | 0.21 | | 0.62 |
| Lower HbA1c via GLP-1R activation | Bipolar disorder II | Overall | 2 | IVW | 1.33 (0.91 to 1.95) | 0.14 | | 0.46 |
| Lower HbA1c via GLP-1R activation | PTSD | Overall | 2 | IVW | 1.09 (0.84 to 1.41) | 0.52 | | 0.25 |
| Lower HbA1c via GLP-1R activation | Schizophrenia | Overall | 2 | IVW | 1.09 (0.94 to 1.27) | 0.26 | | 0.54 |
| Lower HbA1c via GLP-1R activation | Schizophrenia | Women | 2 | IVW | 0.91 (0.72 to 1.15) | 0.42 | | 0.41 |
| Lower HbA1c via GLP-1R activation | Schizophrenia | Men | 2 | IVW | 1.15 (0.94 to 1.42) | 0.18 | | 0.76 |
| Lower HbA1c via GLP-1R activation | Anorexia nervosa | Overall | 2 | IVW | 1.18 (0.93 to 1.50) | 0.17 | | 0.01 |
| Lower HbA1c via GLP-1R activation | ADHD | Overall | 2 | IVW | 0.97 (0.83 to 1.14) | 0.71 | | 0.18 |
| Lower HbA1c via GLP-1R activation | ADHD | Women | 2 | IVW | 0.89 (0.57 to 1.38) | 0.61 | | 0.40 |
| Lower HbA1c via GLP-1R activation | ADHD | Men | 2 | IVW | 0.86 (0.64 to 1.16) | 0.33 | | 0.99 |
| Lower HbA1c via GLP-1R activation | Autism spectrum disorder | Overall | 2 | IVW | 0.85 (0.66 to 1.08) | 0.18 | | 0.23 |
| Lower HbA1c via GLP-1R activation | Tourette syndrome | Overall | 2 | IVW | 0.57 (0.37 to 0.89) | 0.01 | | 0.73 |
| Lower HbA1c | Depression | Overall | 286 | IVW | 1.00 (1.00 to 1.00) | 0.90 | | <0.001 |
| Lower HbA1c | Depression | Overall | 286 | Weighted median | 1.00 (0.99 to 1.00) | 0.32 | | |
| Lower HbA1c | Depression | Overall | 286 | MR Egger | 1.00 (0.99 to 1.01) | 0.67 | 0.55 | |
| Lower HbA1c | Depression | Overall | 286 | MR-RAPS | 1.00 (1.00 to 1.00) | 0.92 | | |
| Lower HbA1c | Depression | Women | 193 | IVW | 1.00 (0.99 to 1.01) | 0.92 | | 0.01 |
| Lower HbA1c | Depression | Women | 193 | Weighted median | 1.00 (0.99 to 1.02) | 0.59 | | |
| Lower HbA1c | Depression | Women | 193 | MR Egger | 1.01 (0.99 to 1.02) | 0.49 | 0.44 | |

| | | | | | | | | |
|---|---|---|---|---|---|---|---|---|
| Lower HbA1c | Depression | Women | 193 | MR-RAPS | 1.00 (0.99 to 1.01) | 0.85 | | |
| Lower HbA1c | Depression | Men | 168 | IVW | 1.00 (0.99 to 1.01) | 0.90 | | 0.60 |
| Lower HbA1c | Depression | Men | 168 | Weighted median | 1.00 (0.98 to 1.02) | 0.85 | | |
| Lower HbA1c | Depression | Men | 168 | MR Egger | 0.99 (0.97 to 1.01) | 0.45 | 0.41 | |
| Lower HbA1c | Depression | Men | 168 | MR-RAPS | 1.00 (0.99 to 1.01) | 0.62 | | |
| Lower HbA1c | Post-partum depression | Women | 192 | IVW | 1.00 (0.99 to 1.01) | 0.41 | | 0.002 |
| Lower HbA1c | Post-partum depression | Women | 192 | Weighted median | 0.99 (0.98 to 1.01) | 0.49 | | |
| Lower HbA1c | Post-partum depression | Women | 192 | MR Egger | 1.00 (0.98 to 1.02) | 0.82 | 0.74 | |
| Lower HbA1c | Post-partum depression | Women | 192 | MR-RAPS | 1.00 (0.99 to 1.01) | 0.42 | | |
| Lower HbA1c | Bipolar disorder | Overall | 286 | IVW | 1.01 (0.99 to 1.02) | 0.34 | | <0.001 |
| Lower HbA1c | Bipolar disorder | Overall | 286 | Weighted median | 1.00 (0.99 to 1.01) | 0.54 | | |
| Lower HbA1c | Bipolar disorder | Overall | 286 | MR Egger | 0.99 (0.97 to 1.01) | 0.17 | 0.02 | |
| Lower HbA1c | Bipolar disorder | Overall | 286 | MR-RAPS | 1.00 (1.00 to 1.01) | 0.60 | | |
| Lower HbA1c | Bipolar disorder I | Overall | 284 | IVW | 1.01 (1.00 to 1.02) | 0.25 | | <0.001 |
| Lower HbA1c | Bipolar disorder I | Overall | 284 | Weighted median | 1.00 (0.99 to 1.02) | 0.66 | | |
| Lower HbA1c | Bipolar disorder I | Overall | 284 | MR Egger | 0.99 (0.96 to 1.01) | 0.18 | 0.02 | |
| Lower HbA1c | Bipolar disorder I | Overall | 284 | MR-RAPS | 1.00 (1.00 to 1.01) | 0.28 | | |
| Lower HbA1c | Bipolar disorder II | Overall | 283 | IVW | 1.00 (0.99 to 1.02) | 0.86 | | 0.002 |
| Lower HbA1c | Bipolar disorder II | Overall | 283 | Weighted median | 1.00 (0.97 to 1.02) | 0.80 | | |
| Lower HbA1c | Bipolar disorder II | Overall | 283 | MR Egger | 0.97 (0.95 to 1.00) | 0.06 | 0.02 | |
| Lower HbA1c | Bipolar disorder II | Overall | 283 | MR-RAPS | 1.00 (0.99 to 1.02) | 0.50 | | |
| Lower HbA1c | PTSD | Overall | 300 | IVW | 1.00 (0.99 to 1.01) | 0.64 | | 0.43 |
| Lower HbA1c | PTSD | Overall | 300 | Weighted median | 1.02 (1.00 to 1.03) | 0.07 | | |
| Lower HbA1c | PTSD | Overall | 300 | MR Egger | 1.02 (1.01 to 1.04) | 0.01 | 0.01 | |
| Lower HbA1c | PTSD | Overall | 300 | MR-RAPS | 1.00 (0.99 to 1.02) | 0.36 | | |
| Lower HbA1c | Schizophrenia | Overall | 287 | IVW | 1.00 (0.99 to 1.01) | 0.45 | | <0.001 |
| Lower HbA1c | Schizophrenia | Overall | 287 | Weighted median | 1.00 (0.98 to 1.01) | 0.37 | | |
| Lower HbA1c | Schizophrenia | Overall | 287 | MR Egger | 0.99 (0.97 to 1.01) | 0.17 | 0.03 | |

| | | | | | | | | |
|---|---|---|---|---|---|---|---|---|
| Lower HbA1c | Schizophrenia | Overall | 287 | MR-RAPS | 1.00 (0.99 to 1.01) | 0.95 | | |
| Lower HbA1c | Schizophrenia | Overall | 286 | IVW (Steiger filtering) | 1.00 (0.99 to 1.01) | 0.53 | | <0.001 |
| Lower HbA1c | Schizophrenia | Overall | 286 | Weighted median (Steiger filtering) | 1.00 (0.98 to 1.01) | 0.36 | | |
| Lower HbA1c | Schizophrenia | Overall | 286 | MR Egger (Steiger filtering) | 0.99 (0.97 to 1.01) | 0.23 | 0.06 | |
| Lower HbA1c | Schizophrenia | Overall | 286 | MR-RAPS (Steiger filtering) | 1.00 (0.99 to 1.01) | >0.999 | | |
| Lower HbA1c | Schizophrenia | Women | 184 | IVW | 0.99 (0.98 to 1.01) | 0.30 | | <0.001 |
| Lower HbA1c | Schizophrenia | Women | 184 | Weighted median | 0.98 (0.97 to 1.00) | 0.05 | | |
| Lower HbA1c | Schizophrenia | Women | 184 | MR Egger | 0.99 (0.97 to 1.01) | 0.33 | 0.64 | |
| Lower HbA1c | Schizophrenia | Women | 184 | MR-RAPS | 1.00 (0.99 to 1.00) | 0.32 | | |
| Lower HbA1c | Schizophrenia | Men | 157 | IVW | 1.00 (0.99 to 1.01) | 0.86 | | <0.001 |
| Lower HbA1c | Schizophrenia | Men | 157 | Weighted median | 1.00 (0.98 to 1.01) | 0.46 | | |
| Lower HbA1c | Schizophrenia | Men | 157 | MR Egger | 0.99 (0.97 to 1.02) | 0.59 | 0.45 | |
| Lower HbA1c | Schizophrenia | Men | 157 | MR-RAPS | 1.00 (0.99 to 1.01) | 0.80 | | |
| Lower HbA1c | Schizophrenia | Men | 156 | IVW (Steiger filtering) | 1.00 (0.99 to 1.01) | 0.93 | | <0.001 |
| Lower HbA1c | Schizophrenia | Men | 156 | Weighted median (Steiger filtering) | 1.00 (0.98 to 1.01) | 0.46 | | |
| Lower HbA1c | Schizophrenia | Men | 156 | MR Egger (Steiger filtering) | 1.00 (0.98 to 1.02) | 0.72 | 0.71 | |
| Lower HbA1c | Schizophrenia | Men | 156 | MR-RAPS (Steiger filtering) | 1.00 (0.99 to 1.01) | 0.87 | | |
| Lower HbA1c | Anorexia nervosa | Overall | 246 | IVW | 1.02 (1.00 to 1.03) | 0.02 | | <0.001 |
| Lower HbA1c | Anorexia nervosa | Overall | 246 | Weighted median | 1.01 (0.99 to 1.02) | 0.53 | | |
| Lower HbA1c | Anorexia nervosa | Overall | 246 | MR Egger | 1.01 (0.99 to 1.03) | 0.42 | 0.60 | |

| | | | | | | | | |
|---|---|---|---|---|---|---|---|---|
| Lower HbA1c | Anorexia nervosa | Overall | 246 | MR-RAPS | 1.02 (1.00 to 1.03) | 0.01 | | |
| Lower HbA1c | ADHD | Overall | 270 | IVW | 0.99 (0.98 to 1.00) | 0.02 | | <0.001 |
| Lower HbA1c | ADHD | Overall | 270 | Weighted median | 0.99 (0.98 to 1.01) | 0.28 | | |
| Lower HbA1c | ADHD | Overall | 270 | MR Egger | 1.01 (0.99 to 1.02) | 0.46 | 0.01 | |
| Lower HbA1c | ADHD | Overall | 270 | MR-RAPS | 0.99 (0.98 to 1.00) | 0.004 | | |
| Lower HbA1c | ADHD | Women | 150 | IVW | 1.00 (0.98 to 1.02) | 0.92 | | 0.71 |
| Lower HbA1c | ADHD | Women | 150 | Weighted median | 1.01 (0.97 to 1.04) | 0.75 | | |
| Lower HbA1c | ADHD | Women | 150 | MR Egger | 1.01 (0.98 to 1.04) | 0.60 | 0.56 | |
| Lower HbA1c | ADHD | Women | 150 | MR-RAPS | 1.00 (0.98 to 1.02) | 0.80 | | |
| Lower HbA1c | ADHD | Men | 133 | IVW | 0.99 (0.97 to 1.00) | 0.12 | | <0.001 |
| Lower HbA1c | ADHD | Men | 133 | Weighted median | 0.99 (0.96 to 1.01) | 0.21 | | |
| Lower HbA1c | ADHD | Men | 133 | MR Egger | 0.99 (0.97 to 1.02) | 0.63 | 0.66 | |
| Lower HbA1c | ADHD | Men | 133 | MR-RAPS | 0.99 (0.98 to 1.00) | 0.05 | | |
| Lower HbA1c | Autism spectrum disorder | Overall | 250 | IVW | 1.00 (0.99 to 1.02) | 0.43 | | 0.02 |
| Lower HbA1c | Autism spectrum disorder | Overall | 250 | Weighted median | 1.01 (1.00 to 1.03) | 0.11 | | |
| Lower HbA1c | Autism spectrum disorder | Overall | 250 | MR Egger | 1.01 (0.99 to 1.03) | 0.59 | 0.90 | |
| Lower HbA1c | Autism spectrum disorder | Overall | 250 | MR-RAPS | 1.00 (0.99 to 1.02) | 0.42 | | |
| Lower HbA1c | Autism spectrum disorder | Overall | 249 | IVW (Steiger filtering) | 1.00 (0.99 to 1.01) | 0.57 | | 0.19 |
| Lower HbA1c | Autism spectrum disorder | Overall | 249 | Weighted median (Steiger filtering) | 1.01 (1.00 to 1.03) | 0.10 | | |
| Lower HbA1c | Autism spectrum disorder | Overall | 249 | MR Egger (Steiger filtering) | 1.01 (0.99 to 1.03) | 0.52 | 0.69 | |
| Lower HbA1c | Autism spectrum disorder | Overall | 249 | MR-RAPS (Steiger filtering) | 1.00 (0.99 to 1.01) | 0.47 | | |
| Lower HbA1c | Tourette syndrome | Overall | 255 | IVW | 0.99 (0.97 to 1.01) | 0.18 | | 0.46 |
| Lower HbA1c | Tourette syndrome | Overall | 255 | Weighted median | 0.99 (0.96 to 1.02) | 0.37 | | |
| Lower HbA1c | Tourette syndrome | Overall | 255 | MR Egger | 1.00 (0.97 to 1.04) | 0.96 | 0.35 | |

| | | | | | | |
|---|---|---|---|---|---|---|
| | Lower HbA1c | Tourette syndrome | Overall | 255 | MR-RAPS | 0.99 (0.97 to 1.01) | 0.31 |

ADHD, attention deficit hyperactivity disorder; BMI, body mass index; GLP-1R, glucagon-like peptide-1 receptor; HbA1c, glycated hemoglobin; IVW, inverse variance weighting; MR, Mendelian randomization; PTSD, post-traumatic stress disorder; RAPS, robust adjusted profile score. Estimates are presented as odds ratio per 1-kg/$m^2$ decrease in BMI or per 1-mmol/mol decrease in HbA1c.

**eTable 9. Sensitivity MR analyses for the associations of GLP-1R activation, lower BMI, and lower HbA1c with the risk of substance use disorders.**

| Exposure | Outcome | SNPs | Method | Beta (95% CI) | *P* value | *P* value (intercept) | *P* value (Q) |
|---|---|---|---|---|---|---|---|
| Lower BMI via GLP-1R activation | Substance use disorders | 2 | IVW | -0.07 (-0.14 to 0.001) | 0.05 | | 0.05 |
| Lower BMI via GLP-1R activation | Cannabis use disorder | 3 | IVW | -0.34 (-0.77 to 0.09) | 0.12 | | 0.83 |
| Lower BMI via GLP-1R activation | Cannabis use disorder | 3 | Weighted median | -0.30 (-0.80 to 0.19) | 0.23 | | |
| Lower BMI via GLP-1R activation | Cannabis use disorder | 3 | MR Egger | 0.28 (-2.14 to 2.69) | 0.82 | 0.61 | |
| Lower BMI via GLP-1R activation | Cannabis use disorder | 3 | MR-RAPS | -0.34 (-0.81 to 0.13) | 0.15 | | |
| Lower BMI via GLP-1R activation | Alcohol dependence | 3 | IVW | -0.05 (-0.59 to 0.48) | 0.85 | | 0.21 |
| Lower BMI via GLP-1R activation | Alcohol dependence | 3 | Weighted median | -0.02 (-0.69 to 0.65) | 0.96 | | |
| Lower BMI via GLP-1R activation | Alcohol dependence | 3 | MR Egger | -2.67 (-5.63 to 0.30) | 0.08 | 0.08 | |
| Lower BMI via GLP-1R activation | Alcohol dependence | 3 | MR-RAPS | -0.05 (-0.62 to 0.51) | 0.86 | | |
| Lower BMI via GLP-1R activation | AUDIT (total score) | 3 | IVW | -0.03 (-0.06 to 0.001) | 0.06 | | 0.91 |
| Lower BMI via GLP-1R activation | AUDIT (total score) | 3 | Weighted median | -0.03 (-0.06 to 0.003) | 0.08 | | |
| Lower BMI via GLP-1R activation | AUDIT (total score) | 3 | MR Egger | -0.06 (-0.22 to 0.10) | 0.48 | 0.72 | |
| Lower BMI via GLP-1R activation | AUDIT (total score) | 3 | MR-RAPS | -0.03 (-0.06 to 0.004) | 0.08 | | |
| Lower BMI via GLP-1R activation | AUDIT (alcohol consumption) | 3 | IVW | -0.02 (-0.05 to 0.01) | 0.16 | | 0.91 |
| Lower BMI via GLP-1R activation | AUDIT (alcohol consumption) | 3 | Weighted median | -0.02 (-0.05 to 0.01) | 0.23 | | |
| Lower BMI via GLP-1R activation | AUDIT (alcohol consumption) | 3 | MR Egger | -0.05 (-0.20 to 0.10) | 0.50 | 0.67 | |
| Lower BMI via GLP-1R activation | AUDIT (alcohol consumption) | 3 | MR-RAPS | -0.02 (-0.05 to 0.01) | 0.20 | | |
| Lower BMI via GLP-1R activation | AUDIT (alcohol problems) | 3 | IVW | -0.03 (-0.05 to -0.001) | 0.05 | | 0.66 |
| Lower BMI via GLP-1R activation | AUDIT (alcohol problems) | 3 | Weighted median | -0.03 (-0.06 to 0.01) | 0.10 | | |
| Lower BMI via GLP-1R activation | AUDIT (alcohol problems) | 3 | MR Egger | -0.02 (-0.16 to 0.12) | 0.74 | 0.97 | |
| Lower BMI via GLP-1R activation | AUDIT (alcohol problems) | 3 | MR-RAPS | -0.03 (-0.05 to 0.002) | 0.06 | | |
| Lower BMI | Substance use disorders | 359 | IVW | -0.01 (-0.02 to -0.01) | <0.001 | | <0.001 |
| Lower BMI | Substance use disorders | 359 | Weighted median | -0.01 (-0.02 to -0.01) | <0.001 | | |
| Lower BMI | Substance use disorders | 359 | MR Egger | 0.003 (-0.01 to 0.02) | 0.70 | 0.02 | |

| | | | | | | | |
|---|---|---|---|---|---|---|---|
| Lower BMI | Substance use disorders | 359 | MR-RAPS | -0.01 (-0.02 to -0.01) | <0.001 | | |
| Lower BMI | Cannabis use disorder | 453 | IVW | -0.06 (-0.08 to -0.03) | <0.001 | | <0.001 |
| Lower BMI | Cannabis use disorder | 453 | Weighted median | -0.06 (-0.10 to -0.02) | 0.01 | | |
| Lower BMI | Cannabis use disorder | 453 | MR Egger | -0.01 (-0.08 to 0.06) | 0.72 | 0.19 | |
| Lower BMI | Cannabis use disorder | 453 | MR-RAPS | -0.06 (-0.08 to -0.03) | <0.001 | | |
| Lower BMI | Alcohol dependence | 452 | IVW | -0.01 (-0.05 to 0.02) | 0.35 | | 0.14 |
| Lower BMI | Alcohol dependence | 452 | Weighted median | 0.002 (-0.05 to 0.06) | 0.94 | | |
| Lower BMI | Alcohol dependence | 452 | MR Egger | 0.04 (-0.04 to 0.12) | 0.34 | 0.15 | |
| Lower BMI | Alcohol dependence | 452 | MR-RAPS | -0.01 (-0.05 to 0.02) | 0.39 | | |
| Lower BMI | Alcohol dependence | 451 | IVW (Steiger filtering) | -0.01 (-0.04 to 0.02) | 0.44 | | 0.26 |
| Lower BMI | Alcohol dependence | 451 | Weighted median (Steiger filtering) | 0.003 (-0.05 to 0.06) | 0.93 | | |
| Lower BMI | Alcohol dependence | 451 | MR Egger (Steiger filtering) | 0.04 (-0.04 to 0.12) | 0.31 | 0.15 | |
| Lower BMI | Alcohol dependence | 451 | MR-RAPS (Steiger filtering) | -0.01 (-0.04 to 0.02) | 0.43 | | |
| Lower BMI | AUDIT (total score) | 452 | IVW | 0.003 (0.001 to 0.01) | 0.01 | | <0.001 |
| Lower BMI | AUDIT (total score) | 452 | Weighted median | 0.002 (-0.001 to 0.005) | 0.27 | | |
| Lower BMI | AUDIT (total score) | 452 | MR Egger | 0.002 (-0.004 to 0.01) | 0.53 | 0.71 | |
| Lower BMI | AUDIT (total score) | 452 | MR-RAPS | 0.002 (0.001 to 0.004) | 0.01 | | |
| Lower BMI | AUDIT (total score) | 451 | IVW (Steiger filtering) | 0.003 (0.001 to 0.01) | 0.007 | | <0.001 |
| Lower BMI | AUDIT (total score) | 451 | Weighted median (Steiger filtering) | 0.002 (-0.001 to 0.005) | 0.27 | | |
| Lower BMI | AUDIT (total score) | 451 | MR Egger (Steiger filtering) | 0.001 (-0.004 to 0.01) | 0.63 | 0.54 | |

| | | | | | | | |
|---|---|---|---|---|---|---|---|
| Lower BMI | AUDIT (total score) | 451 | MR-RAPS (Steiger filtering) | 0.002 (0.001 to 0.004) | 0.005 | | |
| Lower BMI | AUDIT (alcohol consumption) | 452 | IVW | 0.003 (0.001 to 0.01) | 0.001 | | <0.001 |
| Lower BMI | AUDIT (alcohol consumption) | 452 | Weighted median | 0.002 (-0.001 to 0.01) | 0.15 | | |
| Lower BMI | AUDIT (alcohol consumption) | 452 | MR Egger | 0.001 (-0.004 to 0.01) | 0.60 | 0.43 | |
| Lower BMI | AUDIT (alcohol consumption) | 452 | MR-RAPS | 0.003 (0.001 to 0.004) | <0.001 | | |
| Lower BMI | AUDIT (alcohol consumption) | 451 | IVW (Steiger filtering) | 0.004 (0.002 to 0.01) | <0.001 | | <0.001 |
| Lower BMI | AUDIT (alcohol consumption) | 451 | Weighted median (Steiger filtering) | 0.002 (-0.001 to 0.01) | 0.14 | | |
| Lower BMI | AUDIT (alcohol consumption) | 451 | MR Egger (Steiger filtering) | 0.001 (-0.004 to 0.01) | 0.70 | 0.30 | |
| Lower BMI | AUDIT (alcohol consumption) | 451 | MR-RAPS (Steiger filtering) | 0.003 (0.001 to 0.004) | <0.001 | | |
| Lower BMI | AUDIT (alcohol problems) | 452 | IVW | -0.001 (-0.003 to 0.001) | 0.16 | | <0.001 |
| Lower BMI | AUDIT (alcohol problems) | 452 | Weighted median | 0.000 (-0.002 to 0.002) | 0.99 | | |
| Lower BMI | AUDIT (alcohol problems) | 452 | MR Egger | 0.000 (-0.005 to 0.005) | 0.98 | 0.54 | |
| Lower BMI | AUDIT (alcohol problems) | 452 | MR-RAPS | -0.001 (-0.003 to 0.000) | 0.11 | | |
| Lower BMI | AUDIT (alcohol problems) | 451 | IVW (Steiger filtering) | -0.001 (-0.003 to 0.001) | 0.20 | | <0.001 |
| Lower BMI | AUDIT (alcohol problems) | 451 | Weighted median (Steiger filtering) | 0.000 (-0.002 to 0.002) | 0.99 | | |
| Lower BMI | AUDIT (alcohol problems) | 451 | MR Egger (Steiger filtering) | -0.000 (-0.005 to 0.004) | 0.91 | 0.67 | |
| Lower BMI | AUDIT (alcohol problems) | 451 | MR-RAPS (Steiger filtering) | -0.001 (-0.003 to 0.000) | 0.13 | | |
| Lower HbA1c via GLP-1R activation | Substance use disorders | 2 | IVW | 0.004 (-0.04 to 0.04) | 0.86 | | 0.63 |
| Lower HbA1c via GLP-1R activation | Cannabis use disorder | 2 | IVW | 0.09 (-0.21 to 0.39) | 0.55 | | 0.22 |

| | | | | | | | |
|---|---|---|---|---|---|---|---|
| Lower HbA1c via GLP-1R activation | Alcohol dependence | 2 | IVW | -0.20 (-0.60 to 0.20) | 0.33 | | 0.68 |
| Lower HbA1c via GLP-1R activation | AUDIT (total score) | 2 | IVW | 0.01 (-0.01 to 0.03) | 0.33 | | 0.47 |
| Lower HbA1c via GLP-1R activation | AUDIT (alcohol consumption) | 2 | IVW | 0.01 (-0.01 to 0.03) | 0.28 | | 0.28 |
| Lower HbA1c via GLP-1R activation | AUDIT (alcohol problems) | 2 | IVW | 0.01 (-0.01 to 0.03) | 0.33 | | 0.79 |
| Lower HbA1c | Substance use disorders | 192 | IVW | -0.001 (-0.004 to 0.002) | 0.49 | | <0.001 |
| Lower HbA1c | Substance use disorders | 192 | Weighted median | -0.000 (-0.003 to 0.003) | 0.83 | | |
| Lower HbA1c | Substance use disorders | 192 | MR Egger | -0.000 (-0.01 to 0.005) | 0.87 | 0.80 | |
| Lower HbA1c | Substance use disorders | 192 | MR-RAPS | -0.001 (-0.003 to 0.001) | 0.22 | | |
| Lower HbA1c | Cannabis use disorder | 253 | IVW | -0.01 (-0.02 to 0.004) | 0.17 | | 0.02 |
| Lower HbA1c | Cannabis use disorder | 253 | Weighted median | -0.01 (-0.03 to 0.01) | 0.27 | | |
| Lower HbA1c | Cannabis use disorder | 253 | MR Egger | -0.001 (-0.03 to 0.02) | 0.92 | 0.45 | |
| Lower HbA1c | Cannabis use disorder | 253 | MR-RAPS | -0.01 (-0.02 to 0.001) | 0.08 | | |
| Lower HbA1c | Alcohol dependence | 252 | IVW | 0.002 (-0.02 to 0.02) | 0.86 | | 0.16 |
| Lower HbA1c | Alcohol dependence | 252 | Weighted median | 0.01 (-0.02 to 0.03) | 0.69 | | |
| Lower HbA1c | Alcohol dependence | 252 | MR Egger | 0.02 (-0.02 to 0.05) | 0.36 | 0.33 | |
| Lower HbA1c | Alcohol dependence | 252 | MR-RAPS | 0.001 (-0.02 to 0.02) | 0.90 | | |
| Lower HbA1c | AUDIT (total score) | 262 | IVW | 0.001 (0.000 to 0.002) | 0.02 | | <0.001 |
| Lower HbA1c | AUDIT (total score) | 262 | Weighted median | 0.000 (-0.001 to 0.002) | 0.62 | | |
| Lower HbA1c | AUDIT (total score) | 262 | MR Egger | -0.001 (-0.003 to 0.001) | 0.44 | 0.01 | |
| Lower HbA1c | AUDIT (total score) | 262 | MR-RAPS | 0.001 (0.000 to 0.002) | 0.01 | | |
| Lower HbA1c | AUDIT (total score) | 261 | IVW (Steiger filtering) | 0.001 (0.000 to 0.002) | 0.02 | | <0.001 |
| Lower HbA1c | AUDIT (total score) | 261 | Weighted median (Steiger filtering) | 0.000 (-0.001 to 0.002) | 0.62 | | |
| Lower HbA1c | AUDIT (total score) | 261 | MR Egger (Steiger filtering) | -0.001 (-0.002 to 0.001) | 0.59 | 0.03 | |
| Lower HbA1c | AUDIT (total score) | 261 | MR-RAPS (Steiger filtering) | 0.001 (0.000 to 0.002) | 0.009 | | |

| | | | | | | | |
|---|---|---|---|---|---|---|---|
| Lower HbA1c | AUDIT (alcohol consumption) | 262 | IVW | 0.001 (0.000 to 0.002) | 0.01 | | <0.001 |
| Lower HbA1c | AUDIT (alcohol consumption) | 262 | Weighted median | 0.000 (-0.001 to 0.002) | 0.71 | | |
| Lower HbA1c | AUDIT (alcohol consumption) | 262 | MR Egger | -0.001 (-0.002 to 0.001) | 0.50 | 0.01 | |
| Lower HbA1c | AUDIT (alcohol consumption) | 262 | MR-RAPS | 0.001 (0.001 to 0.002) | 0.001 | | |
| Lower HbA1c | AUDIT (alcohol consumption) | 261 | IVW (Steiger filtering) | 0.001 (0.000 to 0.002) | 0.009 | | <0.001 |
| Lower HbA1c | AUDIT (alcohol consumption) | 261 | Weighted median (Steiger filtering) | 0.000 (-0.001 to 0.002) | 0.71 | | |
| Lower HbA1c | AUDIT (alcohol consumption) | 261 | MR Egger (Steiger filtering) | -0.000 (-0.002 to 0.001) | 0.65 | 0.02 | |
| Lower HbA1c | AUDIT (alcohol consumption) | 261 | MR-RAPS (Steiger filtering) | 0.001 (0.000 to 0.002) | 0.002 | | |
| Lower HbA1c | AUDIT (alcohol problems) | 262 | IVW | 0.000 (-0.001 to 0.001) | 0.78 | | 0.002 |
| Lower HbA1c | AUDIT (alcohol problems) | 262 | Weighted median | -0.000 (-0.001 to 0.001) | 0.81 | | |
| Lower HbA1c | AUDIT (alcohol problems) | 262 | MR Egger | -0.001 (-0.002 to 0.001) | 0.34 | 0.18 | |
| Lower HbA1c | AUDIT (alcohol problems) | 262 | MR-RAPS | 0.000 (-0.001 to 0.001) | 0.65 | | |

AUDIT, Alcohol Use Disorders Identification Test; BMI, body mass index; GLP-1R, glucagon-like peptide-1 receptor; HbA1c, glycated hemoglobin; IVW, inverse variance weighting; MR, Mendelian randomization; RAPS, robust adjusted profile score. Positive associations with AUDIT scores indicate higher risk of alcohol use disorders. Estimates are presented in standard deviation unit for substance use disorder risk, as logodds for cannabis use disorder and alcohol dependence, and as log10 transformed score for AUDIT scores per 1-kg/$m^2$ decrease in BMI or per 1-mmol/mol decrease in HbA1c. Only two variants (rs4714290 and rs9394581) were used to predict lower BMI via GLP-1R activation because rs17757975 and its proxy were not available in the substance use disorders data.

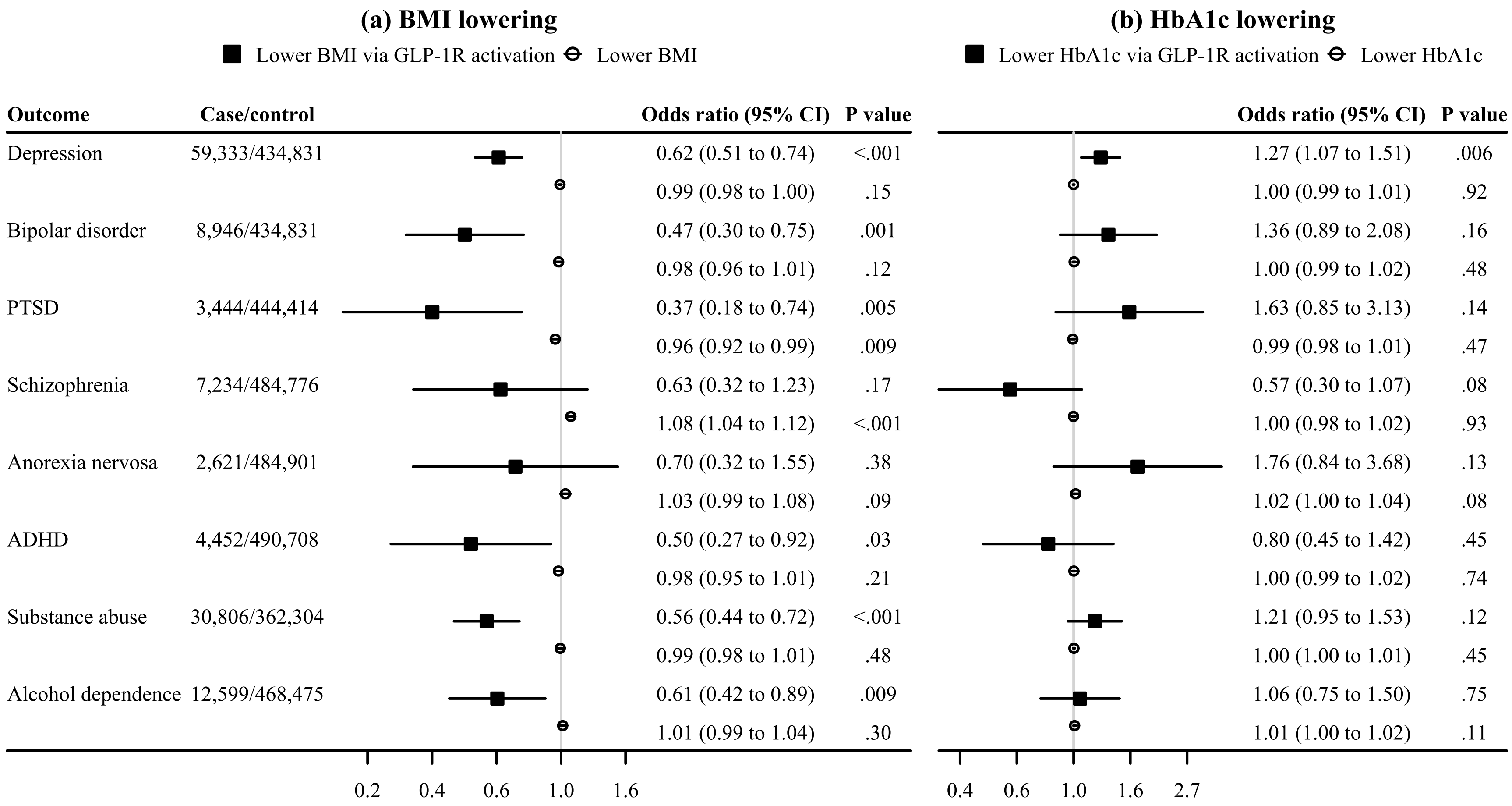

**eFigure 4. IVW MR estimates for the associations of GLP-1R activation with the risk of mental health outcomes in comparison with those for lower BMI and lower HbA1c in FinnGen.**

ADHD, attention deficit hyperactivity disorder; BMI, body mass index; GLP-1R, glucagon-like peptide-1 receptor; HbA1c, glycated hemoglobin; IVW, inverse variance weighted; MR, Mendelian randomization; PTSD, post-traumatic stress disorder. Black squares denote genetically predicted (a) lower BMI or (b) lower HbA1c via GLP-1R activation based on *GLP1R* variants. White circles denote genetically predicted (a) lower BMI or (b) lower HbA1c based on genome-wide variants. Estimates are presented as odds ratio per 1-kg/m$^2$ decrease in BMI or per 1-mmol/mol decrease in HbA1c.

**eTable 10. Sensitivity MR analyses for the associations of GLP-1R activation, lower BMI, and lower HbA1c with mental health outcomes in FinnGen.**

| Exposure | Outcome | SNPs | Method | Odds ratio (95% CI) | *P* value | *P* value (intercept) | *P* value (Q) |
|---|---|---|---|---|---|---|---|
| Lower BMI via GLP-1R activation | Depression | 3 | IVW | 0.66 (0.57 to 0.78) | <0.001 | | 0.21 |
| Lower BMI via GLP-1R activation | Depression | 3 | Weighted median | 0.65 (0.52 to 0.81) | <0.001 | | |
| Lower BMI via GLP-1R activation | Depression | 3 | MR Egger | 0.53 (0.11 to 2.45) | 0.41 | 0.76 | |
| Lower BMI via GLP-1R activation | Depression | 3 | MR-RAPS | 0.66 (0.54 to 0.79) | <0.001 | | |
| Lower BMI via GLP-1R activation | Bipolar disorder | 3 | IVW | 0.45 (0.30 to 0.66) | <0.001 | | 0.85 |
| Lower BMI via GLP-1R activation | Bipolar disorder | 3 | Weighted median | 0.45 (0.28 to 0.75) | 0.002 | | |
| Lower BMI via GLP-1R activation | Bipolar disorder | 3 | MR Egger | 0.82 (0.08 to 8.17) | 0.87 | 0.60 | |
| Lower BMI via GLP-1R activation | Bipolar disorder | 3 | MR-RAPS | 0.45 (0.28 to 0.70) | <0.001 | | |
| Lower BMI via GLP-1R activation | PTSD | 3 | IVW | 0.29 (0.16 to 0.53) | <0.001 | | 0.39 |
| Lower BMI via GLP-1R activation | PTSD | 3 | Weighted median | 0.32 (0.15 to 0.71) | 0.01 | | |
| Lower BMI via GLP-1R activation | PTSD | 3 | MR Egger | 0.68 (0.01 to 57.76) | 0.87 | 0.70 | |
| Lower BMI via GLP-1R activation | PTSD | 3 | MR-RAPS | 0.29 (0.14 to 0.57) | <0.001 | | |
| Lower BMI via GLP-1R activation | Schizophrenia | 3 | IVW | 0.66 (0.37 to 1.18) | 0.16 | | 0.84 |
| Lower BMI via GLP-1R activation | Schizophrenia | 3 | Weighted median | 0.71 (0.36 to 1.40) | 0.32 | | |
| Lower BMI via GLP-1R activation | Schizophrenia | 3 | MR Egger | 0.24 (0.01 to 7.25) | 0.41 | 0.56 | |
| Lower BMI via GLP-1R activation | Schizophrenia | 3 | MR-RAPS | 0.66 (0.35 to 1.24) | 0.19 | | |
| Lower BMI via GLP-1R activation | Anorexia nervosa | 3 | IVW | 0.94 (0.48 to 1.87) | 0.86 | | 0.16 |
| Lower BMI via GLP-1R activation | Anorexia nervosa | 3 | Weighted median | 1.00 (0.41 to 2.45) | 0.999 | | |
| Lower BMI via GLP-1R activation | Anorexia nervosa | 3 | MR Egger | 0.02 (0.000 to 1.14) | 0.06 | 0.06 | |
| Lower BMI via GLP-1R activation | Anorexia nervosa | 3 | MR-RAPS | 0.96 (0.47 to 1.98) | 0.91 | | |
| Lower BMI via GLP-1R activation | ADHD | 3 | IVW | 0.37 (0.21 to 0.62) | <0.001 | | 0.09 |
| Lower BMI via GLP-1R activation | ADHD | 3 | Weighted median | 0.39 (0.18 to 0.83) | 0.02 | | |
| Lower BMI via GLP-1R activation | ADHD | 3 | MR Egger | 9.79 (0.44 to 218.60) | 0.15 | 0.04 | |
| Lower BMI via GLP-1R activation | ADHD | 3 | MR-RAPS | 0.35 (0.19 to 0.64) | <0.001 | | |

| | | | | | | | |
|---|---|---|---|---|---|---|---|
| Lower BMI via GLP-1R activation | Substance abuse | 3 | IVW | 0.68 (0.55 to 0.85) | <0.001 | | 0.002 |
| Lower BMI via GLP-1R activation | Substance abuse | 3 | Weighted median | 0.66 (0.48 to 0.92) | 0.01 | | |
| Lower BMI via GLP-1R activation | Substance abuse | 3 | MR Egger | 0.43 (0.01 to 33.31) | 0.70 | 0.83 | |
| Lower BMI via GLP-1R activation | Substance abuse | 3 | MR-RAPS | 0.62 (0.48 to 0.79) | <0.001 | | |
| Lower BMI via GLP-1R activation | Alcohol dependence | 3 | IVW | 0.77 (0.56 to 1.06) | 0.11 | | 0.02 |
| Lower BMI via GLP-1R activation | Alcohol dependence | 3 | Weighted median | 0.75 (0.48 to 1.18) | 0.21 | | |
| Lower BMI via GLP-1R activation | Alcohol dependence | 3 | MR Egger | 0.47 (0.002 to 90.91) | 0.78 | 0.85 | |
| Lower BMI via GLP-1R activation | Alcohol dependence | 3 | MR-RAPS | 0.73 (0.52 to 1.02) | 0.07 | | |
| Lower BMI | Depression | 500 | IVW | 0.99 (0.98 to 1.00) | 0.15 | | <0.001 |
| Lower BMI | Depression | 500 | Weighted median | 1.00 (0.99 to 1.02) | 0.85 | | |
| Lower BMI | Depression | 500 | MR Egger | 1.01 (0.97 to 1.04) | 0.71 | 0.32 | |
| Lower BMI | Depression | 500 | MR-RAPS | 1.00 (0.99 to 1.00) | 0.38 | | |
| Lower BMI | Bipolar disorder | 500 | IVW | 0.98 (0.96 to 1.01) | 0.12 | | <0.001 |
| Lower BMI | Bipolar disorder | 500 | Weighted median | 0.97 (0.94 to 1.01) | 0.17 | | |
| Lower BMI | Bipolar disorder | 500 | MR Egger | 0.99 (0.93 to 1.06) | 0.76 | 0.77 | |
| Lower BMI | Bipolar disorder | 500 | MR-RAPS | 0.98 (0.96 to 1.01) | 0.15 | | |
| Lower BMI | PTSD | 500 | IVW | 0.96 (0.92 to 0.99) | 0.01 | | 0.01 |
| Lower BMI | PTSD | 500 | Weighted median | 1.00 (0.95 to 1.06) | 0.97 | | |
| Lower BMI | PTSD | 500 | MR Egger | 1.07 (0.98 to 1.17) | 0.12 | 0.01 | |
| Lower BMI | PTSD | 500 | MR-RAPS | 0.97 (0.93 to 1.00) | 0.03 | | |
| Lower BMI | Schizophrenia | 500 | IVW | 1.08 (1.04 to 1.12) | <0.001 | | <0.001 |
| Lower BMI | Schizophrenia | 500 | Weighted median | 1.10 (1.04 to 1.17) | <0.001 | | |
| Lower BMI | Schizophrenia | 500 | MR Egger | 1.02 (0.93 to 1.11) | 0.74 | 0.17 | |
| Lower BMI | Schizophrenia | 500 | MR-RAPS | 1.09 (1.05 to 1.12) | <0.001 | | |
| Lower BMI | Anorexia nervosa | 500 | IVW | 1.03 (0.99 to 1.08) | 0.09 | | 0.001 |
| Lower BMI | Anorexia nervosa | 500 | Weighted median | 1.05 (0.99 to 1.11) | 0.12 | | |
| Lower BMI | Anorexia nervosa | 500 | MR Egger | 1.08 (0.97 to 1.20) | 0.16 | 0.40 | |
| Lower BMI | Anorexia nervosa | 500 | MR-RAPS | 1.03 (1.00 to 1.07) | 0.08 | | |

| | | | | | | | |
|---|---|---|---|---|---|---|---|
| Lower BMI | ADHD | 500 | IVW | 0.98 (0.95 to 1.01) | 0.21 | | <0.001 |
| Lower BMI | ADHD | 500 | Weighted median | 0.99 (0.94 to 1.05) | 0.78 | | |
| Lower BMI | ADHD | 500 | MR Egger | 0.98 (0.90 to 1.07) | 0.62 | 0.97 | |
| Lower BMI | ADHD | 500 | MR-RAPS | 0.98 (0.96 to 1.01) | 0.29 | | |
| Lower BMI | Substance abuse | 500 | IVW | 0.99 (0.98 to 1.01) | 0.48 | | <0.001 |
| Lower BMI | Substance abuse | 500 | Weighted median | 1.01 (0.99 to 1.04) | 0.23 | | |
| Lower BMI | Substance abuse | 500 | MR Egger | 1.08 (1.03 to 1.13) | 0.002 | <0.001 | |
| Lower BMI | Substance abuse | 500 | MR-RAPS | 1.00 (0.98 to 1.01) | 0.44 | | |
| Lower BMI | Alcohol dependence | 500 | IVW | 1.01 (0.99 to 1.04) | 0.30 | | <0.001 |
| Lower BMI | Alcohol dependence | 500 | Weighted median | 1.04 (1.01 to 1.08) | 0.01 | | |
| Lower BMI | Alcohol dependence | 500 | MR Egger | 1.09 (1.02 to 1.16) | 0.01 | 0.01 | |
| Lower BMI | Alcohol dependence | 500 | MR-RAPS | 1.01 (1.00 to 1.03) | 0.10 | | |
| Lower HbA1c via GLP-1R activation | Depression | 2 | IVW | 1.05 (0.94 to 1.18) | 0.36 | | 0.01 |
| Lower HbA1c via GLP-1R activation | Bipolar disorder | 2 | IVW | 1.25 (0.94 to 1.65) | 0.12 | | 0.60 |
| Lower HbA1c via GLP-1R activation | PTSD | 2 | IVW | 1.29 (0.84 to 1.98) | 0.24 | | 0.34 |
| Lower HbA1c via GLP-1R activation | Schizophrenia | 2 | IVW | 0.77 (0.51 to 1.16) | 0.21 | | 0.22 |
| Lower HbA1c via GLP-1R activation | Anorexia nervosa | 2 | IVW | 1.33 (0.82 to 2.16) | 0.25 | | 0.32 |
| Lower HbA1c via GLP-1R activation | ADHD | 2 | IVW | 0.92 (0.63 to 1.34) | 0.65 | | 0.53 |
| Lower HbA1c via GLP-1R activation | Substance abuse | 2 | IVW | 1.12 (0.96 to 1.31) | 0.15 | | 0.42 |
| Lower HbA1c via GLP-1R activation | Alcohol dependence | 2 | IVW | 0.97 (0.77 to 1.21) | 0.77 | | 0.50 |
| Lower HbA1c | Depression | 291 | IVW | 1.00 (0.99 to 1.01) | 0.92 | | <0.001 |
| Lower HbA1c | Depression | 291 | Weighted median | 1.00 (0.99 to 1.01) | 0.52 | | |
| Lower HbA1c | Depression | 291 | MR Egger | 1.00 (0.99 to 1.01) | 0.76 | 0.77 | |
| Lower HbA1c | Depression | 291 | MR-RAPS | 1.00 (1.00 to 1.01) | 0.66 | | |
| Lower HbA1c | Bipolar disorder | 291 | IVW | 1.00 (0.99 to 1.02) | 0.48 | | 0.01 |
| Lower HbA1c | Bipolar disorder | 291 | Weighted median | 1.01 (0.99 to 1.03) | 0.17 | | |
| Lower HbA1c | Bipolar disorder | 291 | MR Egger | 1.01 (0.99 to 1.03) | 0.35 | 0.52 | |
| Lower HbA1c | Bipolar disorder | 291 | MR-RAPS | 1.00 (0.99 to 1.02) | 0.42 | | |

| | | | | | | | |
|---|---|---|---|---|---|---|---|
| Lower HbA1c | PTSD | 291 | IVW | 0.99 (0.98 to 1.01) | 0.47 | | 0.03 |
| Lower HbA1c | PTSD | 291 | Weighted median | 0.99 (0.97 to 1.02) | 0.63 | | |
| Lower HbA1c | PTSD | 291 | MR Egger | 1.01 (0.97 to 1.04) | 0.66 | 0.32 | |
| Lower HbA1c | PTSD | 291 | MR-RAPS | 0.99 (0.98 to 1.01) | 0.50 | | |
| Lower HbA1c | Schizophrenia | 291 | IVW | 1.00 (0.98 to 1.02) | 0.93 | | 0.01 |
| Lower HbA1c | Schizophrenia | 291 | Weighted median | 0.99 (0.97 to 1.02) | 0.68 | | |
| Lower HbA1c | Schizophrenia | 291 | MR Egger | 0.99 (0.96 to 1.02) | 0.50 | 0.45 | |
| Lower HbA1c | Schizophrenia | 291 | MR-RAPS | 1.00 (0.98 to 1.02) | 0.88 | | |
| Lower HbA1c | Anorexia nervosa | 291 | IVW | 1.02 (1.00 to 1.04) | 0.08 | | 0.04 |
| Lower HbA1c | Anorexia nervosa | 291 | Weighted median | 1.02 (0.99 to 1.06) | 0.17 | | |
| Lower HbA1c | Anorexia nervosa | 291 | MR Egger | 1.01 (0.97 to 1.05) | 0.53 | 0.71 | |
| Lower HbA1c | Anorexia nervosa | 291 | MR-RAPS | 1.02 (1.00 to 1.04) | 0.09 | | |
| Lower HbA1c | ADHD | 291 | IVW | 1.00 (0.99 to 1.02) | 0.74 | | 0.16 |
| Lower HbA1c | ADHD | 291 | Weighted median | 1.00 (0.98 to 1.03) | 0.93 | | |
| Lower HbA1c | ADHD | 291 | MR Egger | 1.01 (0.98 to 1.04) | 0.61 | 0.69 | |
| Lower HbA1c | ADHD | 291 | MR-RAPS | 1.00 (0.99 to 1.02) | 0.56 | | |
| Lower HbA1c | Substance abuse | 291 | IVW | 1.00 (1.00 to 1.01) | 0.45 | | <0.001 |
| Lower HbA1c | Substance abuse | 291 | Weighted median | 1.00 (0.99 to 1.01) | 0.75 | | |
| Lower HbA1c | Substance abuse | 291 | MR Egger | 1.00 (0.99 to 1.02) | 0.72 | 0.95 | |
| Lower HbA1c | Substance abuse | 291 | MR-RAPS | 1.00 (1.00 to 1.01) | 0.43 | | |
| Lower HbA1c | Alcohol dependence | 291 | IVW | 1.01 (1.00 to 1.02) | 0.11 | | <0.001 |
| Lower HbA1c | Alcohol dependence | 291 | Weighted median | 1.00 (0.99 to 1.02) | 0.78 | | |
| Lower HbA1c | Alcohol dependence | 291 | MR Egger | 1.00 (0.98 to 1.02) | 0.74 | 0.15 | |
| Lower HbA1c | Alcohol dependence | 291 | MR-RAPS | 1.01 (1.00 to 1.02) | 0.15 | | |

ADHD, attention deficit hyperactivity disorder; BMI, body mass index; GLP-1R, glucagon-like peptide-1 receptor; HbA1c, glycated hemoglobin; IVW, inverse variance weighting; MR, Mendelian randomization; PTSD, post-traumatic stress disorder; RAPS, robust adjusted profile score. Estimates are presented as odds ratio per 1-kg/$m^2$ decrease in BMI or per 1-mmol/mol decrease in HbA1c.

**eTable 11. The posterior probabilities of different hypotheses in pairwise colocalization analyses for BMI and each significant outcome at the *GLP1R* gene.**

| Outcome | SNPs | $H_0$ | $H_1$ | $H_2$ | $H_3$ | $H_4$ | Conditional $H_4$ |
|---|---|---|---|---|---|---|---|
| Well-being spectrum | 2133 | <0.1% | 29.4% | <0.1% | 11.3% | 59.3% | 84.0% |
| Life satisfaction | 2133 | <0.1% | 13.4% | <0.1% | 12.9% | 73.8% | 85.2% |
| Positive affect | 2133 | <0.1% | 17.9% | <0.1% | 14.0% | 68.1% | 82.9% |
| Neuroticism | 2133 | <0.1% | 24.5% | <0.1% | 14.4% | 61.1% | 80.9% |
| Depressive symptoms | 2133 | <0.1% | 24.2% | <0.1% | 13.0% | 62.7% | 82.8% |
| Depression | 2394 | <0.1% | 88.0% | <0.1% | 8.2% | 3.8% | 31.3% |
| Bipolar disorder | 2375 | <0.1% | 50.0% | <0.1% | 4.1% | 45.9% | 91.8% |
| Bipolar disorder I | 2336 | <0.1% | 60.7% | <0.1% | 4.5% | 34.7% | 88.5% |

$H_0$, no association with either trait; $H_1$, association with BMI only; $H_2$, association with outcome only; $H_3$, associations of two independent variants and one for each trait; $H_4$, associations of one shared variant with both traits; conditional $H_4$, associations of one shared variant with both traits conditional on the presence of a variant associated with outcome ($H_4/(H_2+H_3+H_4)$).
